# A genome-wide association study in Chinese children identifies and functionally validates 5q31.1 as a neuroblastoma susceptibility locus

**DOI:** 10.1101/2025.08.11.25333455

**Authors:** Linzehao Li, Yingchao Song, Qiwen Zheng, Junjie Ge, Yue Jiang, Bichen Peng, Xuedi Yu, Jingfu Wang, Sharon Diskin, John Maris, Yiju Wei, Hakon Hakonarson, Xiao Chang

**Affiliations:** College of Medical Information and Artificial Intelligence, Shandong First Medical University, Jinan, China; China National Center for Bioinformation, Beijing, China; Beijing Institute of Genomics, Chinese Academy of Sciences, Beijing, China; Department of Pediatric Oncology, Shandong Cancer Hospital and Institute, Shandong First Medical University, Jinan, China; Division of Oncology and Center for Childhood Cancer Research, Children’s Hospital of Philadelphia, USA; Medical Science and Technology Innovation Center, Shandong First Medical University, Jinan, China; The Center for Applied Genomics, Children’s Hospital of Philadelphia, Philadelphia, USA

## Abstract

**Background:** Neuroblastoma, the most common extracranial solid tumor in children, exhibits considerable clinical heterogeneity influenced by genetic predisposition. While genome-wide association studies (GWAS) in European populations have identified eight susceptibility loci, the genetic basis of neuroblastoma in East Asian populations remains poorly understood.

**Methods:** We conducted the first GWAS in a Chinese cohort comprising 235 neuroblastoma patients and 3,100 controls, followed by multi-omics analyses of gene expression. The novel risk loci were further validated in an independent East Asian cohort (76 cases/269 controls). Functional characterization of a novel locus was carried out in neuroblastoma cell lines using CRISPR/Cas9-mediated deletion and overexpression assays to evaluate its regulatory effects on candidate genes.

**Findings:** We replicated six of eight known loci including genome-wide significant associations at *CASC15* (6p22.3; P = 1.55 × 10⁻⁹) and *BARD1* (2q35; P = 3.44 × 10⁻⁷), and identified 11 novel risk loci. These novel associations implicate genes involved in DNA repair (*MUTYH* at 1p34.1), neurodevelopment (*BASP1* at 4q13.2 and S*LC22A4/SLC22A5* at 5q31.1), and immune regulation (HLA at 6p21 and *IDO1/IDO2* at 5q31.1). Multi-omics integration revealed that lead variants modulate gene expression (cis-eQTLs) and DNA methylation (mQTLs) in neural crest-derived tissues and immune cells. Two loci (rs2631372 at 5q31.1: P= 0.045; rs2956095 at 11p13: P= 0.027) showed consistent associations in the replication cohort. Functional studies demonstrated that deletion of the 5q31.1 risk interval reduced expression of *SLC22A4*, *SLC22A5*, and *LOC553103*, while their overexpression promoted neuroblastoma cell proliferation.

**Interpretation:** These findings highlight both shared and population-specific genetic contributions to neuroblastoma susceptibility, underscoring the importance of diversifying GWAS efforts to advance ancestry-informed risk assessment and therapeutic strategies.

## Introduction

Neuroblastoma, an embryonal malignancy derived from sympathetic nervous system precursors, represents the most common extracranial solid tumor in children, accounting for approximately 8–10% of all pediatric cancers and about 15% of pediatric cancer mortality ^1,2^. Its clinical course is highly heterogeneous, ranging from spontaneous regression or maturation into benign ganglioneuromas in some infants, to aggressive metastatic disease despite intensive multimodal treatment in others. Prognostic outcomes are influenced significantly by age at diagnosis, tumor stage, and genetic factors, notably *MYCN* amplification.

Genetic susceptibility plays a crucial role in neuroblastoma etiology. Genome-wide association studies (GWAS) over the past decade, predominantly conducted in populations of European ancestry, have successfully identified eight risk loci ^3–9^. These discoveries have illuminated key pathways in neural crest differentiation and tumorigenesis. Notably, five of these loci, including 2q35 (*BARD1*), 3q25.32 (*MLF1*), 6p22.3 (*CASC15*), 6q16.3 (*HACE1*/*LIN28B*), and 11p15.4 (*LMO1*), have also been validated in Chinese cohorts ^10–14^. Furthermore, *BARD1* and *LMO1* have been replicated in a Korean population^15^, suggesting shared genetic susceptibility between individuals of European and East Asian ancestries, particularly among Chinese populations. However, GWAS data from East Asian populations remain markedly underrepresented, with only a single GWAS reported in a Korean cohort. To date, no GWAS specifically addressing neuroblastoma susceptibility in Chinese populations has been conducted, constituting a critical knowledge gap given potential population-specific genetic architecture.

To address this gap, we undertook the first genome-wide association study of neuroblastoma risk in a Chinese cohort. Our study aims to validate previously identified loci from European-ancestry studies and identify potential genetic variants that uniquely contribute to neuroblastoma susceptibility in the Chinese population. These findings are expected to enhance our understanding of neuroblastoma genetics and support the development of ancestry-informed risk assessment tools and targeted prevention strategies.

## Results

### Genome-wide association study in Han Chinese individuals

To explore potential genetic loci associated with neuroblastoma in Chinese populations, we conducted a GWAS study comprising 235 neuroblastoma patients and 3,100 cancer-free controls. The demographic and clinical information of the cases was shown in Table S1, the patient population exhibited balanced sex distribution (male:55.6%), with INSS stage 4 disease predominating (stage 4: 90.6%) and the majority classified as high-risk (67.2%). As an initial step, we examined whether eight genome-wide significant loci previously identified in individuals of European ancestry were also associated with disease risk in our Chinese cohort. Six of the eight loci showed at least nominal evidence of association (*P* < 0.05; Supplementary Table S2), including a genome-wide significant signal at 6p22 (rs6939340, *CASC15*; *P* = 1.55 × 10⁻⁹, OR = 0.48) and a near-genome-wide significant association at 2q35 (rs6435862, *BARD1*; *P* = 3.44 × 10⁻⁷, OR = 1.86). These results indicate the robustness of our analytical framework and support the cross-population generalizability of key neuroblastoma risk loci between European and East Asian populations.

Beyond the validation of known risk loci, our GWAS revealed 11 additional loci that surpassed the genome-wide significance threshold (Table 1, Figure 1 and Figure S1). Several of the newly identified loci harbor genes with compelling functional relevance. At 1p34.1, the lead variant rs3219489 is a missense variant in *MUTYH*, a gene involved in base excision repair. The signal at 4q24 maps near *TACR3*, encoding the neurokinin-3 receptor implicated in neuronal signaling. *BASP1* (4q13.2), *SLC22A4*/*SLC22A5* (5q31.1), and *PNPLA3*/*SAMM50* (11p13) are also located near lead variants, with roles in neurodevelopment, mitochondrial function, and metabolic regulation, respectively. Notably, the association at 6p21 falls within the extended HLA region, suggesting a possible immunogenetic component in neuroblastoma susceptibility. Additional loci include *IDO1*/*IDO2* (5q31.1), enzymes involved in tryptophan metabolism and immune modulation, and *APIP*/*PDHX* (8p11.21), genes implicated in apoptosis and cellular energy metabolism. Finally, the signal at 22q13.31 maps to a region containing *PNPLA3*, previously associated with lipid processing and inflammation.

**Figure 1.**
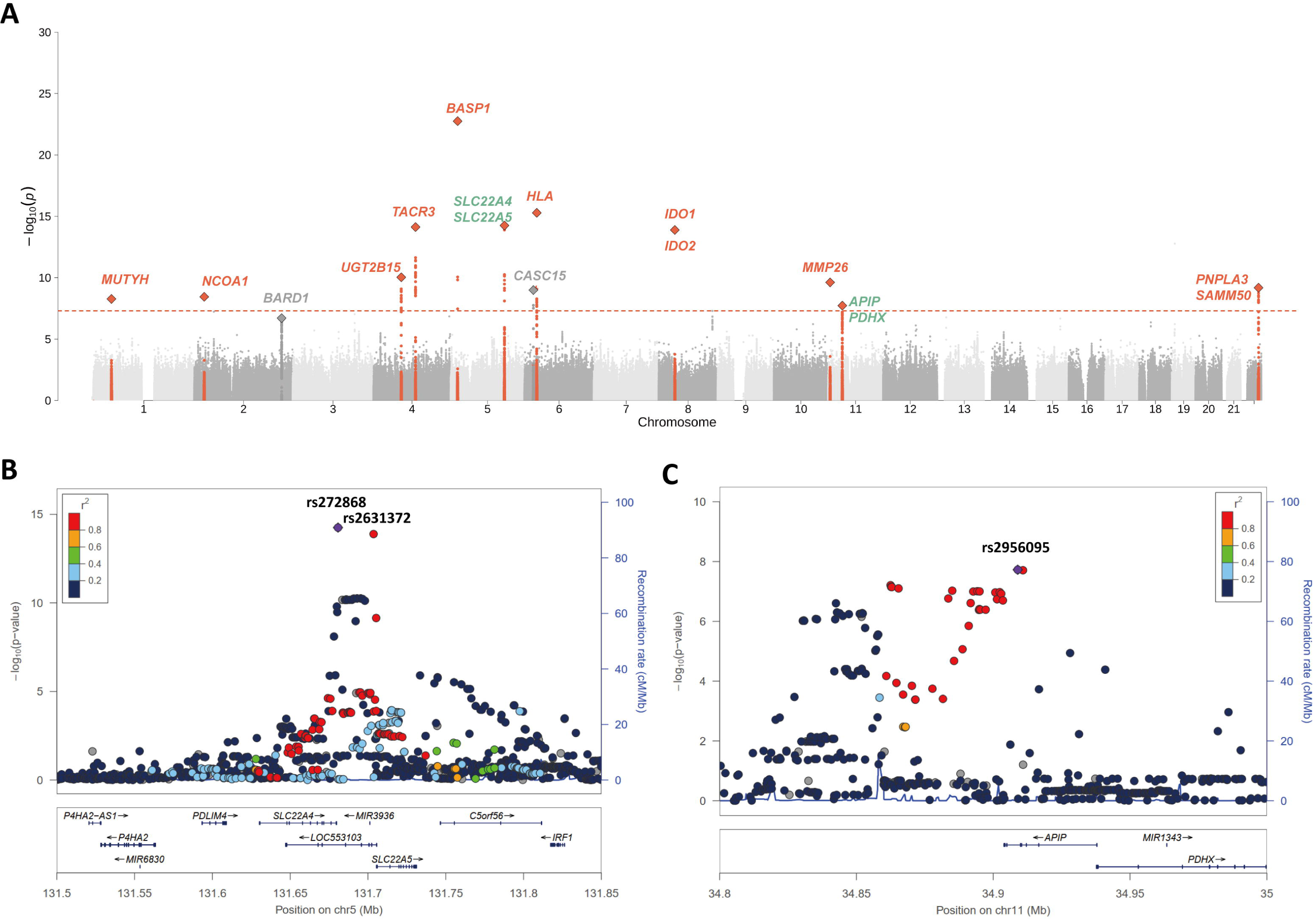
Genome-wide association study (GWAS) of neuroblastoma in Chinese populations. (A) Manhattan plot displaying GWAS results. Novel neuroblastoma-associated loci (red) and previously reported loci (gray) are highlighted. (B) Regional association plot for the 5q31.1 locus. SNPs are colored by linkage disequilibrium (LD) with the lead variant rs272868 (purple diamond). (C) Regional association plot for the 11p13 locus, with SNPs colored by LD relative to rs2956095 (purple diamond).

**Table 1.** Genome-wide association study (GWAS) results of neuroblastoma risk loci in the Chinese discovery cohort.

| SNP | A1 | A2 | CHR | BP | OR | SE | STAT | P | Gene | Locus | Annotation |
| --- | --- | --- | --- | --- | --- | --- | --- | --- | --- | --- | --- |
| rs3219489 | G | C | 1 | 45331833 | 1.936 | 0.1132 | 5.836 | 5.36E-09 | *MUTYH* | 1p34.1 | new |
| rs2165738 | C | G | 2 | 24469940 | 2.208 | 0.1342 | 5.902 | 3.60E-09 | *NCOA1* | 2p23.3 | new |
| rs10024137 | A | G | 4 | 68667043 | 0.2737 | 0.1999 | -6.479 | 9.21E-11 | *UGT2B15* | 4q13.2 | new |
| rs3733631 | C | G | 4 | 103719946 | 2.502 | 0.1179 | 7.776 | 7.46E-15 | *TACR3* | 4q24 | new |
| rs2962370 | G | C | 5 | 17215335 | 3.093 | 0.1131 | 9.985 | 1.77E-23 | *BASP1* | 5p15.1 | new |
| rs272868 | C | G | 5 | 132345058 | 2.479 | 0.1162 | 7.813 | 5.59E-15 | *SLC22A4/SLC22A5* | 5q31.1 | new |
| rs9358491 | A | G | 6 | 22139885 | 0.4812 | 0.1198 | -6.107 | 1.02E-09 | *CASC15* | 6p22.3 | known |
| rs2534815 | C | G | 6 | 30531350 | 2.703 | 0.1227 | 8.106 | 5.22E-16 | *HLA* | 6p21.32 | new |
| rs2160860 | A | T | 8 | 39965626 | 2.337 | 0.1101 | 7.707 | 1.29E-14 | *IDO1/IDO2* | 8p11.21 | new |
| rs2196122 | C | G | 11 | 4864318 | 2.173 | 0.1226 | 6.333 | 2.40E-10 | *MMP26* | 11p15.4 | new |
| rs2956095 | C | T | 11 | 34887382 | 2.374 | 0.1537 | 5.623 | 1.87E-08 | *APIP/PDHX* | 11p13 | new |
| rs28550680 | T | C | 22 | 43950046 | 0.3999 | 0.1484 | -6.176 | 6.56E-10 | *PNPLA3/SAMM50* | 22q13.31 | new |

To further characterize the regulatory potential of these novel risk loci, we examined expression and methylation QTLs using multiple complementary datasets. Single-tissue eQTLs from the Genotype-Tissue Expression (GTEx) project (v8), single-cell eQTLs (sc-eQTLs) from scQTLbase^16^, and methylation QTLs (mQTLs) from recent large-scale studies ^17–20^ are summarized in Tables S3–S6. Notably, several lead variants, including rs3219489 (1p34.1), rs2165738 (2p23.3), rs3733631 (4q24), rs272868 (5q31.1), rs2160860 (8p11.21), rs2956095 (11p13), and rs28550680 (22q13.31), exhibited consistent regulatory effects across multiple omics layers. For instance, rs272868 (5q31.1) was identified as a cis-eQTL for *P4HA2*, *PDLIM4*, *SLC22A4*, and *SLC22A5* in several tissues, including thyroid, tibial nerve, spleen, whole blood, and multiple brain regions (Table S3). This variant also displayed cell-type-specific regulatory effects in CD4⁺ and CD8⁺ T cells, naïve B cells, and monocytes (Table S4), and was associated with extensive DNA methylation changes, with the strongest mQTL signal detected at probe cg24060327 near *SLC22A5* (*P* = 1.55 × 10⁻²¹⁸; Table S5). Similarly, rs2956095 (11p13) was an sc-eQTL for *APIP* in both immune cells (CD4⁺ T cells, CD8⁺ T cells, monocytes) and neuronal populations (inhibitory and excitatory neurons), and was linked to differential methylation at two nearby CpG sites (cg06937548: *P* = 3.84 × 10⁻⁵⁵; cg15745106: *P* = 3.11 × 10⁻¹⁴). These convergent regulatory signals support a functional role for these risk variants in modulating gene expression and epigenetic state in disease-relevant tissues and cell types.

### Replication of novel associations in an independent East Asian cohort

To support the reliability of these associations, we compared the minor allele frequencies (MAFs) of lead variants in our cases and controls, and examined consistency with allele frequencies reported in East Asian populations from gnomAD ^21^ and GenomeAsia100K ^22^. We found that MAFs in controls were highly concordant with reference frequencies from both databases (Table S7), indicating high-quality genotyping and confirming that the observed associations are unlikely to be driven by population stratification or technical artifacts.

To further validate the novel loci identified in the discovery phase, we performed a replication GWAS in an independent East Asian cohort from the Children’s Hospital of Philadelphia, comprising 76 neuroblastoma cases and 269 cancer-free controls. Despite the limited statistical power due to modest sample size, two loci showed nominal evidence of association with neuroblastoma risk: rs2631372 at 5q31.1 (in high LD with rs272868, r² = 0.99; *P* = 0.045, OR = 1.49) and rs2956095 at 11p13 (*P* = 0.027, OR = 0.51) (Figure 1B, C and Table S8). Moreover, the direction of effect for both variants was consistent with that observed in the discovery cohort, lending further support to their potential involvement in neuroblastoma susceptibility.

### Assessing cis-regulatory potential at the 5q31.1 GWAS locus

We next prioritized the 5q31.1 locus for functional validation based on its convergence of genetic, transcriptomic, and epigenomic evidence. Regional association analysis identified rs272868 and rs2631372 as the lead variants (Figure 1B), both residing within an intergenic region flanked by *SLC22A5* and *SLC22A4*. This region also overlaps the lncRNA gene *LOC553103*, raising the possibility that it harbors cis-regulatory elements controlling gene expression. Moreover, both bulk and single-cell eQTL analyses demonstrated that rs272868 is significantly associated with the expression of *SLC22A5*, *SLC22A4*, and *LOC553103*, as well as more distal genes such as *P4HA2* and *PDLIM4* (Tables S3-S4). Notably, all of these genes have been implicated in cancer-related pathways, further supporting the functional relevance of this locus. Furthermore, Epigenomic annotation from ENCODE revealed that the region surrounding these SNPs is enriched for canonical features of regulatory activity, including candidate cis-regulatory elements (cCREs), DNase I hypersensitive sites, and histone modifications such as H3K27ac and H3K4me1 (Figure S2). Several transcription factor binding clusters also overlap this region, suggesting an open and transcriptionally active chromatin state in relevant cell types. Collectively, these lines of evidence nominate the 5q31.1 intergenic region as a putative regulatory hub, warranting direct experimental interrogation.

To investigate whether the neuroblastoma-associated region at 5q31.1 exerts cis-regulatory effects, we employed CRISPR/Cas9-mediated deletion targeting the genomic intervals flanking rs2631372 and rs272868. For each variant, two independent sgRNAs (designated A1 and A2) were designed using the MIT CRISPR design tool and cloned into a dual-sgRNA lentiviral vector (pLV[2gRNA]-lentiCRISPR-v2-Puro; Table S9). Sanger sequencing confirmed successful integration of the guide sequences, with no mismatches relative to the designed targets (Figure 2A). Following lentiviral transduction and antibiotic selection in neuroblastoma cells, we assessed gene expression changes via qRT-PCR. Deletion of the regions surrounding either rs272868 or rs2631372 led to significant downregulation of *SLC22A5*, *SLC22A4*, and *LOC553103*, whereas expression of the more distal genes *P4HA2* and *PDLIM4* remained unaffected (Figure 2B). These findings suggest that the intergenic region specifically regulates nearby genes in neuroblastoma cells rather than exerting broad transcriptional effects across the locus.

**Figure 2.**
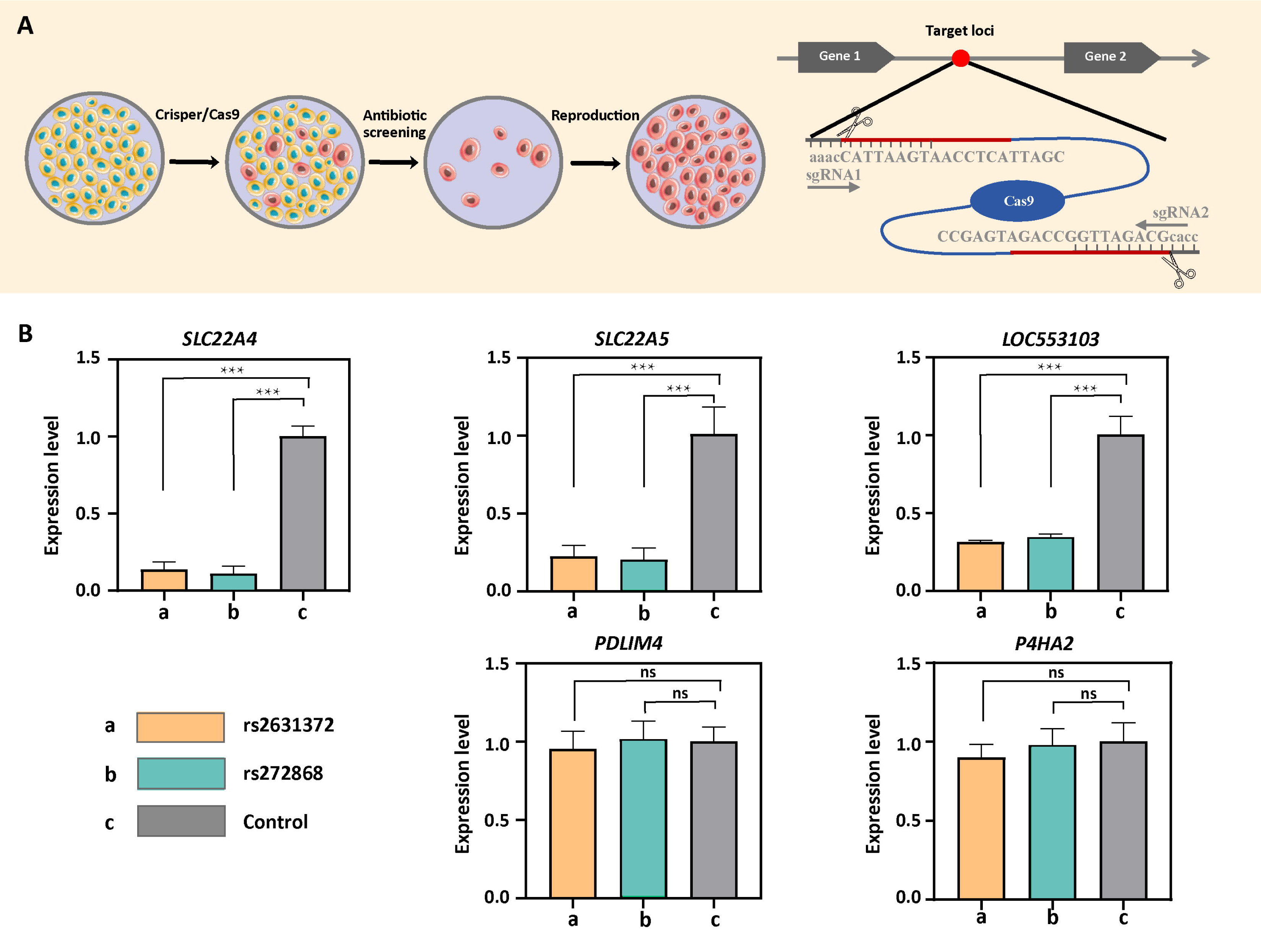
CRISPR-Cas9-mediated gene knockout and validation in SH-SY5Y cells. (A) Lentiviral delivery of sgRNA targeting SH-SY5Y cells. Lentiviral vectors encoding custom sgRNAs were transduced into cells, inducing frameshift mutations at target loci. Transduced cells were selected with puromycin (1 μg/mL, 72 h) and neomycin (6μg/mL, 72 h), then expanded for analysis. (B) qRT-PCR analysis of gene expression in CRISPR-edited cells. Knockout of the target locus significantly altered expression of *SLC22A4* (*P* < 0.001), *SLC22A5* (*P* < 0.001), and *LOC553103* (*P* < 0.001) compared to controls. No significant changes were observed for *PDLIM4* or *P4HA2* (*P* > 0.05). Data represent mean ± SEM; n = 3 biological replicates. Statistical significance: ns (not significant), *P* > 0.05); *, *P* < 0.05; **, *P* < 0.01; ***, *P* < 0.001.

### Overexpression of 5q31.1 target genes promotes neuroblastoma cell proliferation

To investigate whether altered expression of 5q31.1 target genes contributes to neuroblastoma cell growth, we conducted CCK-8 assays following lentiviral overexpression of *SLC22A4*, *SLC22A5*, and the lncRNA *LOC553103* in SH-SY5Y cells (Table S10). Each of these genes significantly promoted cell proliferation compared to control, with effects emerging on day 2 and persisting through day 4 (Figure 3A). Similarly, we also added *PDMLIM4* and *P4HA2*, which although have a certain impact on the proliferation of SH-SY5Y, their effect is not as satisfactory as the above three genes. These proliferative effects were reproducible across three replicates. Electron microscopy at day 3 further confirmed these findings at the morphological level, showing visibly denser cell populations in the *SLC22A4*, *SLC22A5*, and *LOC553103* groups. The overexpression of *PDLIM4* also showed a slightly denser trend in cell morphology, while *P4HA2* exhibited a lower cell morphology (Figure 3B). These observations suggest that regulatory elements at the 5q31.1 locus modulate neuroblastoma proliferation through direct control of nearby gene expression.

**Figure 3.**
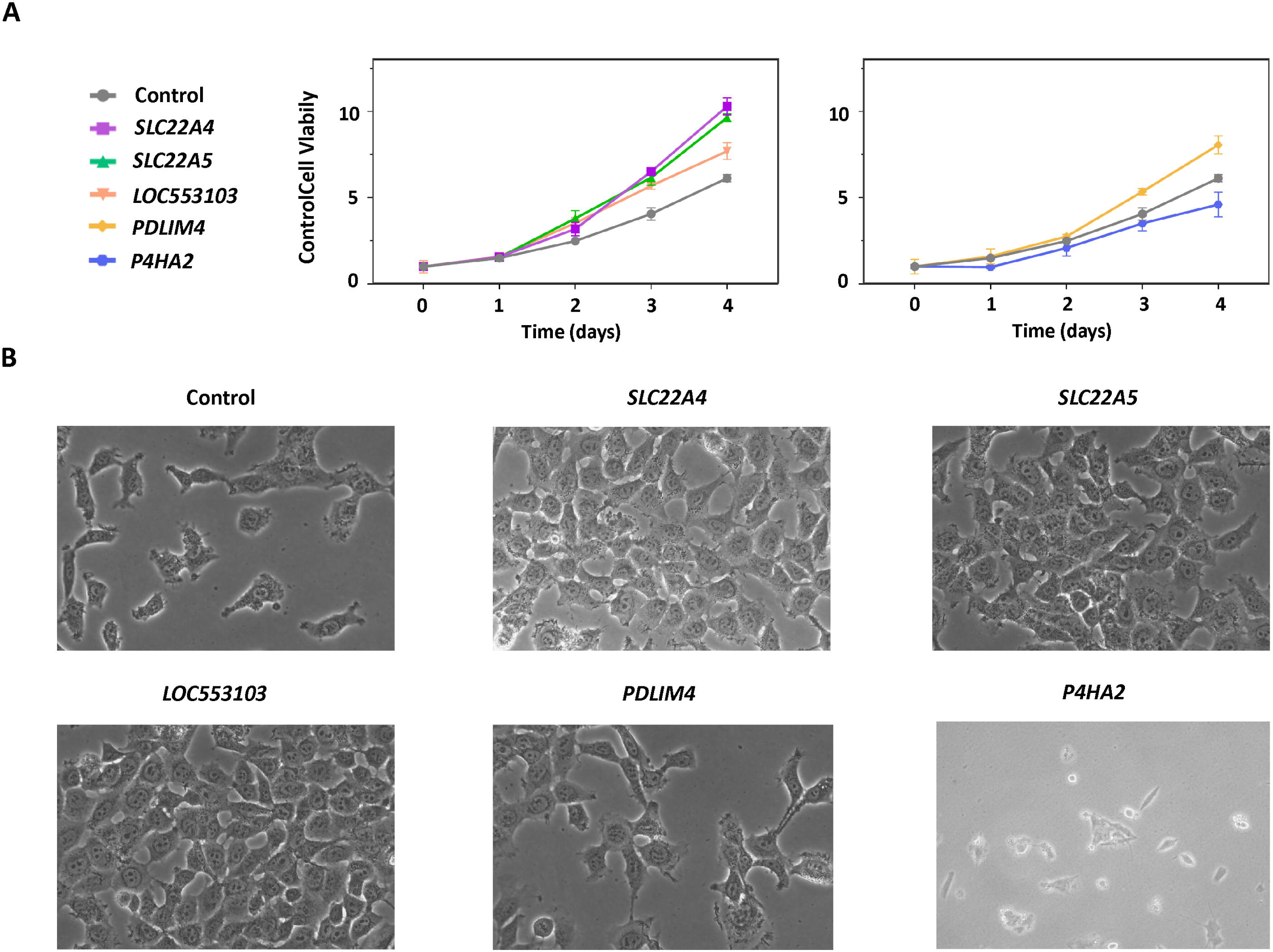
Functional effects of gene overexpression on SH-SY5Y cell proliferation and morphology. (A) Cell proliferation assessed by CCK-8 assay. Overexpression of *SLC22A4*, *SLC22A5*, or *LOC553103* enhanced proliferation (*P* < 0.001 from Day 2–4). PDLIM4-overexpressing cells showed increased proliferation at Day 3 (*P* < 0.001), whereas *P4HA2*-overexpressing cells exhibited reduced proliferation (*P* < 0.05 from Day 2 onward). (B) Phase-contrast microscopy (Day 3). Compared to the control group, cells overexpressing *SLC22A4*, *SLC22A5*, or *LOC553103* exhibited markedly increased cell density and compact morphology. *PDLIM4* overexpression also enhanced cell growth but to a lesser extent, while *P4HA2*-overexpressing cells displayed sparse distribution, reduced size, and membrane abnormalities (e.g., blebbing). Scale bars: 100 μm.

## Discussion

This study represents the first GWAS of neuroblastoma conducted in a Chinese population, addressing a substantial and previously unmet need to characterize the genetic underpinnings of this pediatric malignancy in East Asians. By leveraging a large, well-curated cohort with rigorous clinical and genotyping quality control, we not only replicated multiple susceptibility loci initially reported in European populations but also identified novel risk variants that appear specific to Chinese individuals. These findings underscore the critical importance of expanding GWAS efforts beyond populations of European ancestry to capture the full landscape of genetic diversity influencing neuroblastoma susceptibility.

Despite the relatively smaller sample size compared to previously published European GWASs^9^, our study successfully replicated six of the eight known susceptibility loci, including robust associations at *CASC15*, which reached genome-wide significance. This high replication rate may be partly attributable to the clinical composition of our cohort, which predominantly comprised high-risk neuroblastoma cases. Indeed, loci such as *CASC15* and *BARD1*, have been shown to confer stronger effects in high-risk subgroups, suggesting that enrichment for aggressive disease in our cohort may have enhanced statistical power for detecting these associations. In contrast, the lack of replication for *HSD17B12* and *TP53* in our study likely reflects differences in cohort composition and allele frequency. *HSD17B12* was previously identified in low-risk patients^7^, who are underrepresented in our dataset, whereas the top SNP at *TP53* (rs35850753) is nearly exclusive to European populations and virtually absent in East Asians^8^.

Our study identified 11 novel genome-wide significant loci, several of which implicate genes with established roles in DNA repair, neural development, and immune regulation. For example, the association at 1p34.1 involves a missense variant in *MUTYH*, a gene critical for oxidative DNA damage repair ^23^, supporting the role of genomic instability in neuroblastoma pathogenesis. Other loci point to genes involved in neurodevelopmental signaling (*TACR3*, *BASP1*) ^24,25^ and metabolic homeostasis (*PNPLA3*, *SAMM50*) ^26,27^. Notably, one signal resides in the extended HLA region at 6p21, raising the possibility of immunogenetic contributions to disease susceptibility, a hypothesis that warrants further exploration given emerging links between neuroblastoma and host immune response. Additionally, the presence of lead variants near *IDO1* and *IDO2*, which are key regulators of tryptophan metabolism and immune modulation ^28^, further supports a role for immune-metabolic crosstalk in neuroblastoma etiology. These newly identified associations offer promising avenues for future mechanistic investigation and highlight the multifaceted biological processes that may contribute to neuroblastoma risk.

To support the novel associations identified in our discovery analysis, we examined an independent East Asian cohort and observed loci 5q31.1 and 11p13 showing evidence of replication. The 5q31.1 locus emerged as particularly compelling, supported by convergent genetic, transcriptomic, and epigenomic evidence. eQTL analyses implicated rs272868 in the regulation of multiple cancer-related genes, including *SLC22A4* ^29,30^, *SLC22A5* ^31^, *LOC553103* ^32,33^, *P4HA2* ^34,35^ and *PDLIM4* ^36,37^. Functional interrogation using CRISPR/Cas9-mediated deletion of the regions flanking rs272868 and rs2631372 in neuroblastoma cells led to downregulation of *SLC22A4*, *SLC22A5* and *LOC553103*, while overexpression assays further demonstrated their capacity to enhance cell proliferation. These findings nominate 5q31.1 as a putative regulatory hub that modulates neuroblastoma growth. However, it remains unclear whether the regulatory elements at this locus act directly on *SLC22A4* and *SLC22A5*, or instead exert their effects indirectly via modulation of *LOC553103*, which in turn may influence the expression of the adjacent solute carrier genes. Interestingly, both *SLC22A4* and *SLC22A5* have been previously implicated in cancer biology, supporting their potential functional relevance in tumorigenesis ^38–40^. Moreover, *LOC553103* has been reported as a non-specific diagnostic and prognostic biomarker across multiple cancer types, further suggesting a potential role in cancer-related transcriptional regulation ^41^. Further mechanistic studies are needed to delineate the precise regulatory architecture at this locus.

In summary, our study provides new insights into the genetic architecture of neuroblastoma in Chinese children, revealing both shared and population-specific risk loci. By integrating GWAS with eQTL, epigenomic annotation, and functional validation, we demonstrate the utility of a multi-layered approach to prioritize and characterize candidate susceptibility regions. These findings not only expand the catalog of genetic variants implicated in neuroblastoma but also underscore the importance of including diverse populations in genetic studies of pediatric cancer. Continued efforts to increase sample sizes, refine functional annotations, and explore ancestry-specific regulatory mechanisms will be essential to translating GWAS discoveries into biological understanding and, ultimately, novel therapeutic opportunities.

## Materials and methods

### Participants

Neuroblastoma case subjects in the discovery phase were recruited from the Shandong Cancer Hospital and Institute. All participants were ethnic Han Chinese children diagnosed with neuroblastoma and treated or undergoing surgery at the hospital. Peripheral blood samples were collected at the time of diagnosis, and clinical data including age, tumor site, and INSS disease stage were recorded by treating physicians. Control subjects for the discovery cohort were selected from the CAS cohort, a prospective multi-omics study comprising 3,197 adult participants from various institutes or offices of the Chinese Academy of Sciences in Beijing, China ^20,42–44^. Individuals with self-reported histories of cancer were excluded based on questionnaire information. Written informed consent was obtained from all participants or their legal guardians. This study was approved by the Institutional Review Boards of the Shandong Cancer Hospital and Institute, the Beijing Institute of Genomics (Chinese Academy of Sciences) and the Beijing Zhongguancun Hospital.

Replication analysis was performed using neuroblastoma case samples from the Children’s Oncology Group (COG) Neuroblastoma biorepository, collected at diagnosis and annotated with relevant clinical features. Self-reported individuals of East Asian ancestry were selected for inclusion. Control subjects for the replication cohort were recruited through the Center for Applied Genomics (CAG) at the Children’s Hospital of Philadelphia (CHOP), using the same inclusion criteria as previously described: no serious underlying medical conditions and no history of cancer. The study was approved by the Research Ethics Board of CHOP, and informed consent was obtained by trained clinical staff.

### Genotyping, quality control (QC) and imputation

In the discovery cohort, genotyping was conducted using the Infinium Asian Screening Array (ASA), while samples in the replication cohort were genotyped using the Illumina HumanHap550 or HumanHap610 SNP arrays. To assess and control for population stratification, principal component analysis (PCA) was performed using the EIGENSTRAT method. Individuals of East Asian ancestry were retained based on clustering patterns in PCA relative to HapMap3 reference populations (Figure S1).

Samples with a genotype call rate below 95% were excluded. In addition, single nucleotide polymorphisms (SNPs) were filtered out if they had a minor allele frequency (MAF) below 1%, a call rate less than 98%, or deviated from Hardy–Weinberg equilibrium (P < 1×10⁻⁶). To remove related or duplicate individuals, pairwise identity-by-descent (IBD) analysis was carried out using PLINK.

Genotype imputation was performed via the TOPMed Imputation Server using the minimac4 algorithm and the TOPMed reference panel, which consists of whole-genome sequencing data from over 100,000 individuals. For downstream association analysis, we retained imputed variants with MAF > 1% and imputation quality score (Rsq) > 0.3, or MAF between 0.5% and 1% with Rsq > 0.5.

### GWAS Analysis

Association testing was performed using logistic regression under an additive genetic model implemented in PLINK, based on the imputed dosage of the effect allele. The analysis was adjusted for sex and the top five principal components to control for residual population structure. To evaluate potential inflation due to population stratification, we calculated the genomic inflation factor (λ). No notable inflation was detected (λ ≈ 1.0), suggesting minimal confounding and well-controlled population structure in the final dataset.

### Cell Culture

SH-SY5Y human neuroblastoma cells and HEK293T cells were cultured in Dulbecco’s Modified Eagle’s Medium (DMEM) supplemented with 10% fetal bovine serum (FBS), 100 U/mL penicillin, and 100 μg/mL streptomycin at 37°C in a humidified atmosphere containing 5% CO₂. Cells were passaged every 2–3 days upon reaching 70–80% confluence.

### CRISPR/Cas9-mediated gene knockout/editing

Dual-guide RNA (gRNA) sequences targeting the 5q31.1 region were designed using the MIT CRISPR design tool (http://crispr.mit.edu). Synthesized oligonucleotides included 5’ overhangs (CACC and AAAC) for directional cloning into the pLV[2gRNA]-lentiCRISPR-v2-Puro/pLV[2gRNA]-lenticCRISPR-v2-Neo vector via BsmBI enzyme digestion. Lentiviral particles were produced in HEK293T cells maintained in DMEM supplemented with 10% fetal bovine serum (FBS), 100 U/mL penicillin, and 100 μg/mL streptomycin. Cells were transfected at 70–80% confluence using calcium phosphate-DNA precipitation with plasmid constructs and packaging plasmids (pCMV-VSVG, pMDLG/pRRE, and pRSV-Rev). Viral supernatants were harvested at 48 and 72 hours post-transfection, concentrated by ultracentrifugation (50,000 × g, 2 hours, 4°C), and used to transduce SH-SY5Y cells at an MOI of 10 with 8 μg/mL Polybrene. After 48 hours, cells underwent puromycin(1μg/mL) or neomycin selection (6 μg/mL) for 5 days^45^. Monoclonal lines were established by limiting dilution in 96-well plates, and knockout efficiency was confirmed via Sanger sequencing After cultivation, gene expression was measured by quantitative real-time PCR (qPCR).

### Lentiviral-mediated Gene Overexpression

Full-length cDNA sequences of candidate genes were synthesized and cloned into the pCDH-CMV-MCS-EF1a-GFP+Puro lentiviral vector using NEBuilder HiFi DNA Assembly. Lentiviral particles were similarly produced in HEK293T cells, concentrated, and used to transduce SH-SY5Y cells. Stable overexpression cell lines were selected with 2 μg/mL puromycin for 7 days, and overexpression efficiency was validated by qPCR.

### Quantitative Real-time PCR Analysis

Total RNA was extracted from SH-SY5Y cells using commercial RNA extraction kits (Thermo Scientific AM9775, USA) according to the manufacturer’s instructions. RNA purity and concentration were measured using a NanoDrop (Thermo Scientific One/Oneᶜ , USA) spectrophotometer, ensuring A260/A280 ratios between 1.8 and 2.0. 500μg of total RNA was reverse-transcribed using the PrimeScript RT Reagent Kit (TaKaRa Perfect Real Time RR037B, Japan) by 37°C for 2 min, 55°C for 15 min, 85°C for 5 min, 4°C hold. qPCR analysis was performed using BrightCycle Universal SYBR Green qPCR Mix with UDG on a Bio-Rad CFX96 Real-Time PCR System (BIOER Quant Gene 500, China) with a protocol including 37°C for 2 min (UDG incubation), 95°C for 3 min (pre-denaturation), 40 cycles of 95°C for 5 s and 60°C for 32 s (amplification), and a melting curve analysis (instrument default). Primers are detailed in Supplementary Table 8. Relative gene expression was normalized against β-Actin and calculated using the 2^−ΔΔCt method ^46^. Genes exhibiting fold changes ≥2.0 and P < 0.05 (Student’s t-test) were considered significantly regulated.

### Cell Proliferation Assays

Cell proliferation was assessed using the Cell Counting Kit-8 (CCK-8, Beyotime, China) according to the manufacturer’s protocol. SH-SY5Y cells were seeded at 5×10³ cells per well into 96-well plates in RPMI 1640 medium with 10% FBS. Cell viability was measured daily from days 0 to 4 by adding 10 μL of CCK-8 reagent per well and incubating for 2 hours at 37°C. Absorbance was measured at 450 nm using a Bio-Rad microplate reader. Background controls consisted of medium-only wells. Each experiment included three biological replicates with three technical replicates ^47^.

### Transmission Electron Microscopy

SH-SY5Y cells were collected on day 3 post-treatment and fixed with 2.5% glutaraldehyde. Following post-fixation in osmium tetroxide, cells were dehydrated in graded ethanol, embedded in resin, and sectioned. Cell morphology and proliferation density were assessed using a JEOL JEM-1400 electron microscope.

### Statistical Analysis

Data are presented as mean ± standard deviation (SD) from at least three independent experiments. Two-tailed Student’s t-tests were used for comparisons between two groups, and one-way or two-way ANOVA followed by Tukey’s post-hoc tests were applied for multiple comparisons. Statistical analyses were performed using GraphPad Prism software (version 8.0), with significance set at *P* < 0.05.

## Description of Supplemental Information

Supplemental Information includes three figures and ten tables.

## Data Availability

All data produced in the present study are available upon reasonable request to the authors

## Acknowledgements

The study was supported by grants from the National Natural Science Foundation of China (32270661), the Special Funds of Taishan Scholar Project, China (tsqn202211224), and Excellent Youth Science Fund Project (Overseas) of Shandong China (2023HWYQ-082).

