## Supplemental Tables and Figures for "A genome-wide association study in Chinese children identifies and functionally validates 5q31.1 as a neuroblastoma susceptibility locus"

**Li *et al.***

**Supplementary Online Contents**

**Figure S1.** Regional plots of novel genome-wide significant loci.
**Figure S2.** Genomic annotation of the 5q31.1 neuroblastoma risk locus (chr5:132,340,000-132,370,000, hg38) from UCSC Genome Browser.
**Figure S3.** Principal component analysis (PCA) for ancestry quality control in the discovery and replication cohorts.

**Table S1.** Demographics of cases included in the discovery and replication stages of genome-wide association studies.
**Table S2.** Replication of previously reported six high-risk neuroblastoma susceptibility loci in the Chinese population.
**Table S3.** Single-Tissue eQTLs for novel loci identified in the Chinese population.
**Table S4.** Single-Cell eQTLs for novel loci identified in the Chinese population.
**Table S5.** mQTLs for novel loci identified in the Chinese population.
**Table S6.** mQTLs for novel loci identified in the Chinese population using Chinese-derived methylation data.
**Table S7.** Allele frequency comparison of novel risk-associated SNPs across Chinese and East Asian populations.
**Table S8.** Associations of novel risk loci in the replication cohort.
**Table S9.** gRNA Primers for rs2631372 and rs272868.
**Table S10.** Primer design for qPCR gene determination.

**Figure S1.** Regional plots of novel genome-wide significant loci. Purple diamond indicates the lead SNP, and circles represent the other SNPs in the region, with coloring from the linkage disequilibrium ($r^2$, based on the 1000 Genomes Project Europeans/East Asians) between each SNP and the lead SNP.

a. 1p34.1

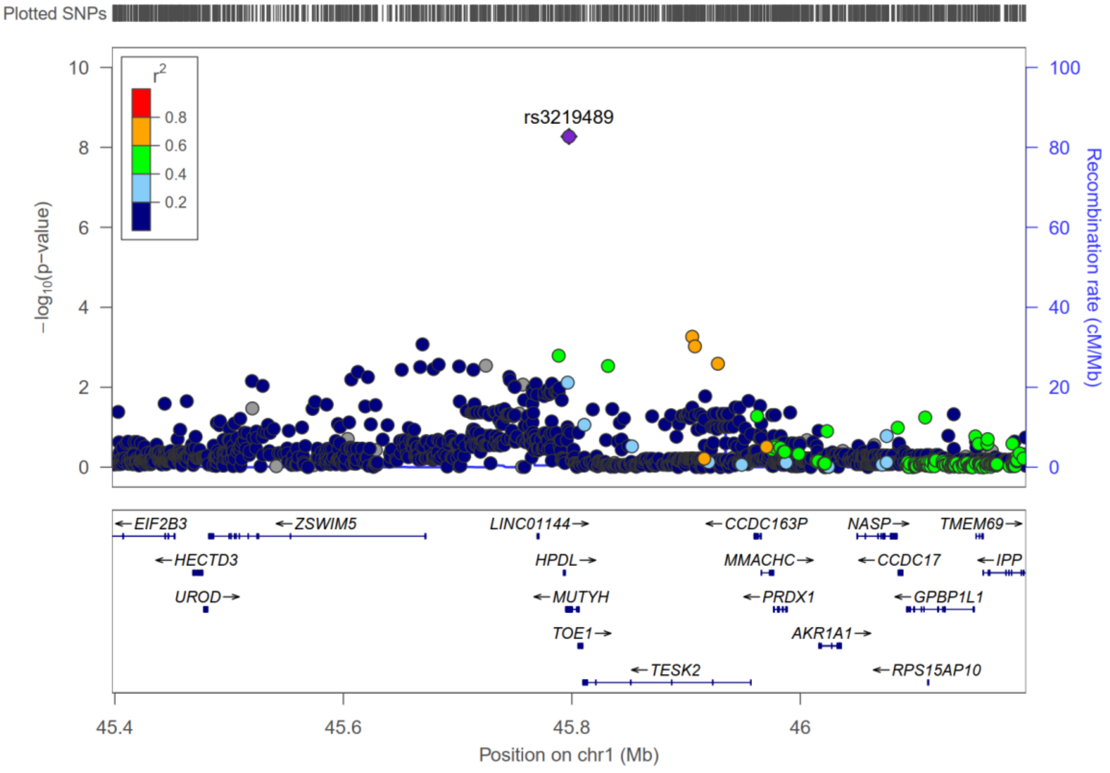

b. 2p23.3

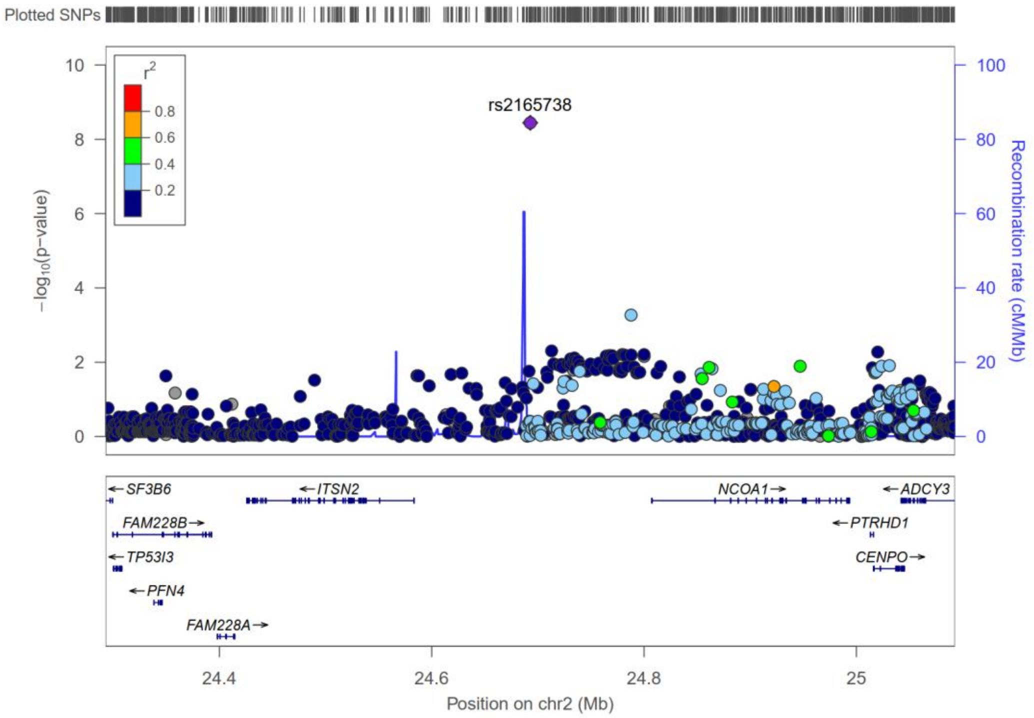


c. 4q13.2

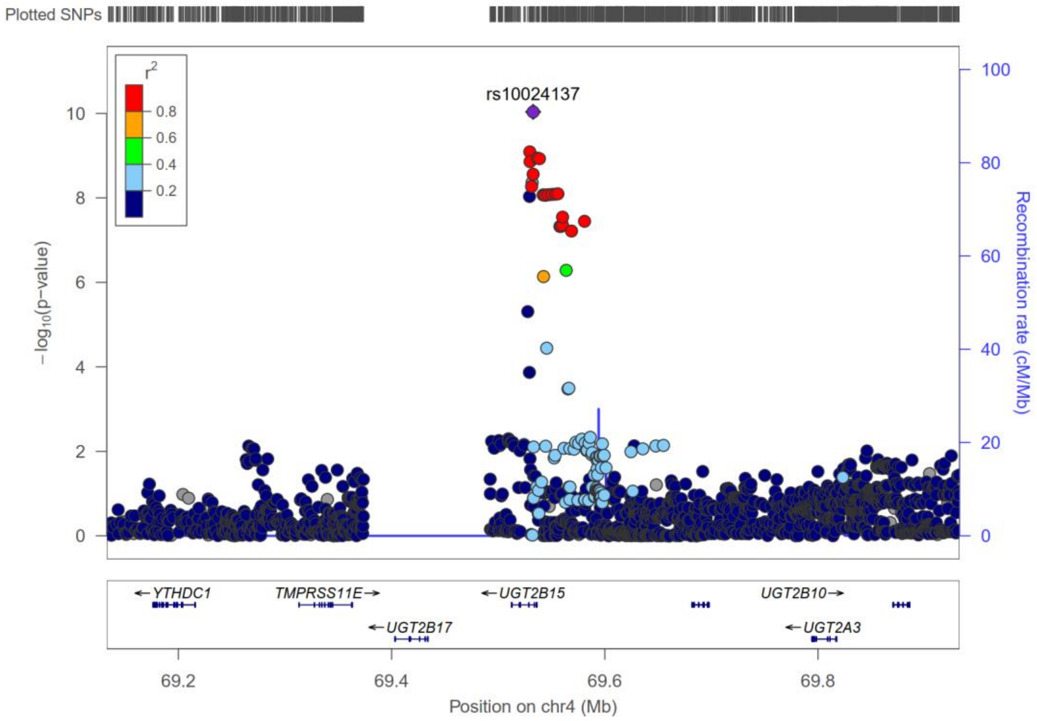

d. 4q24

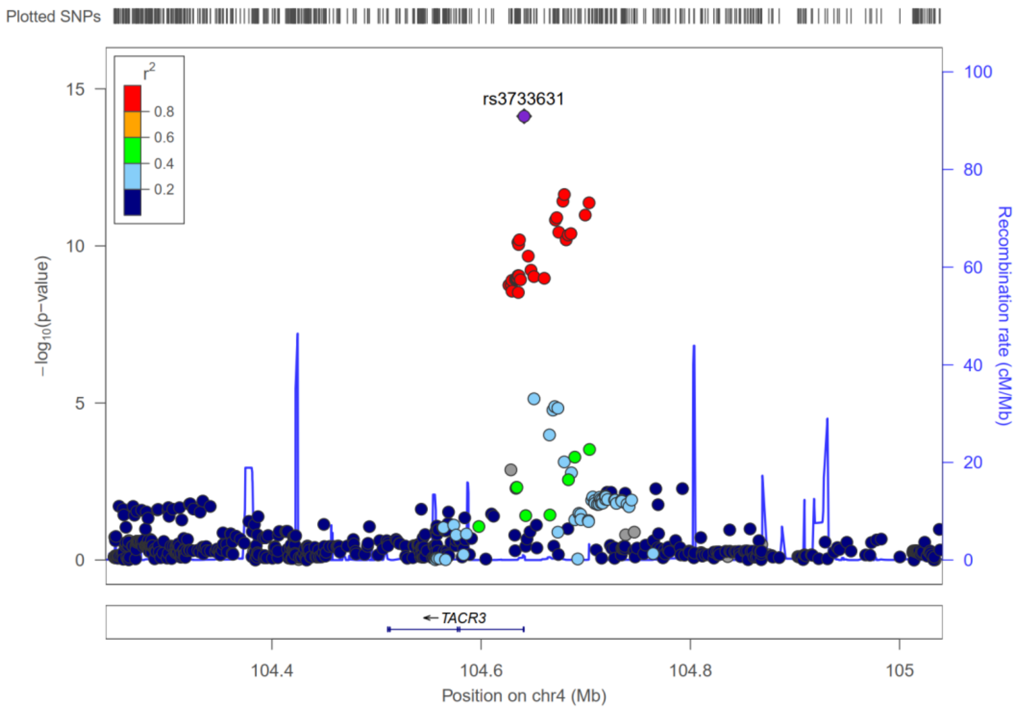


e. 5p15.1

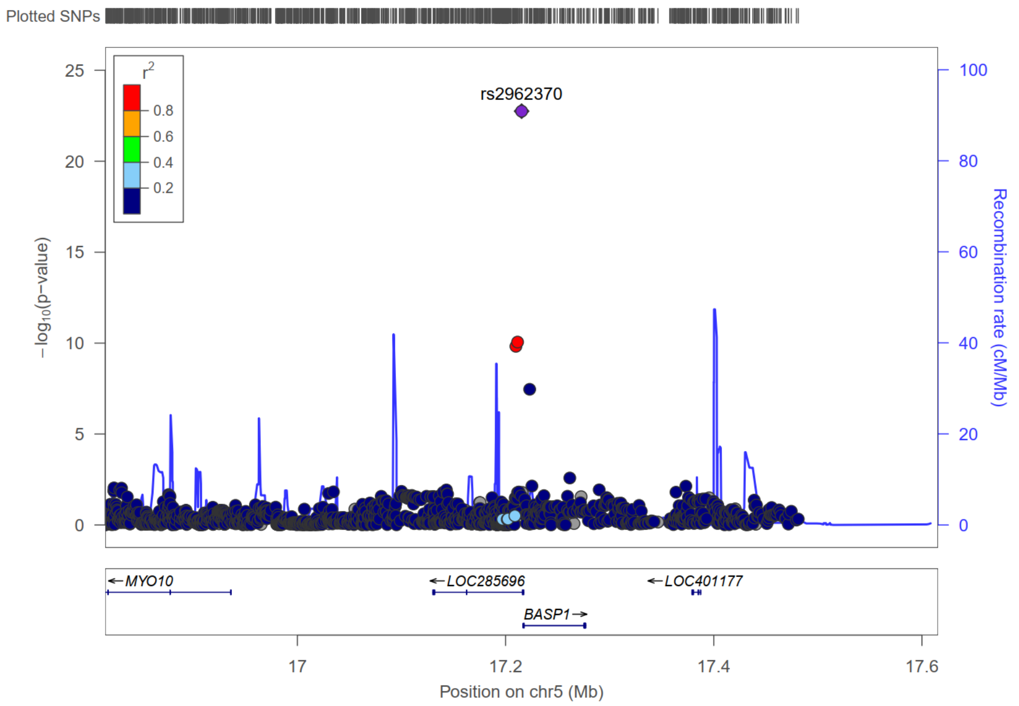

f. 5q31.1

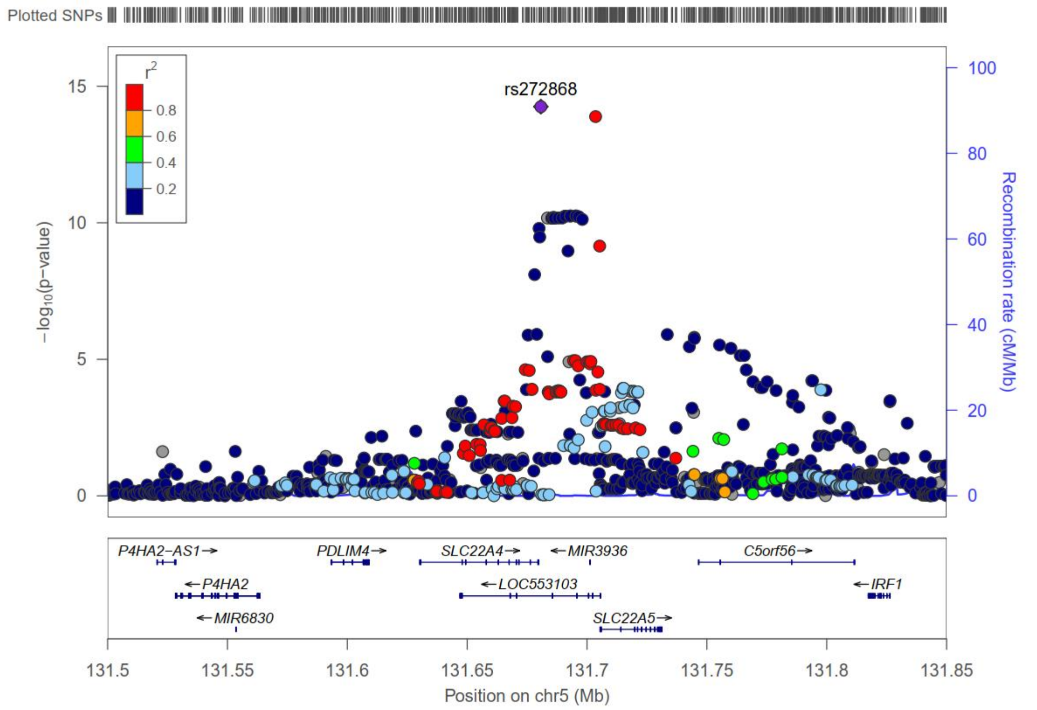


g. 6p21.32

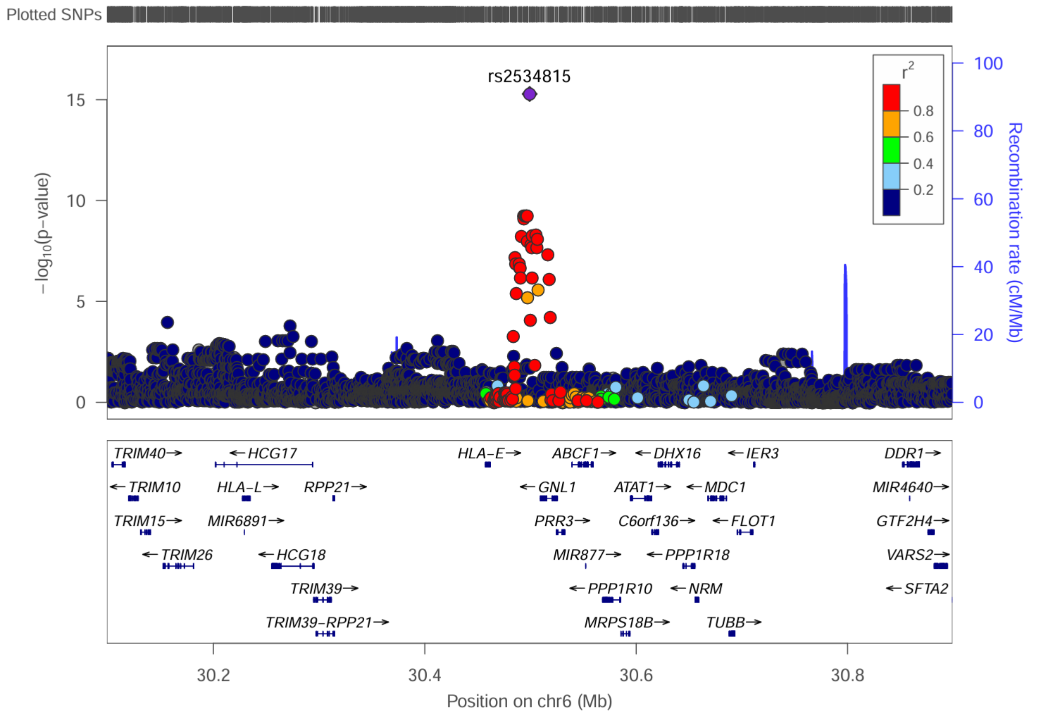

h. 8p11.21

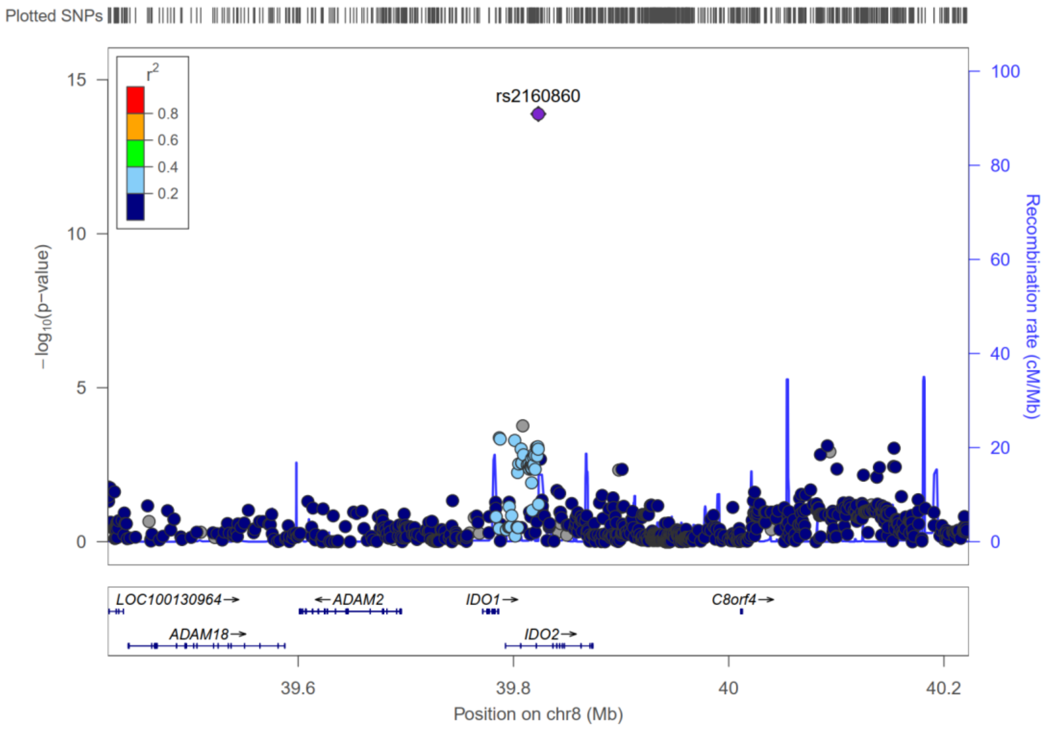


i. 11p15.4

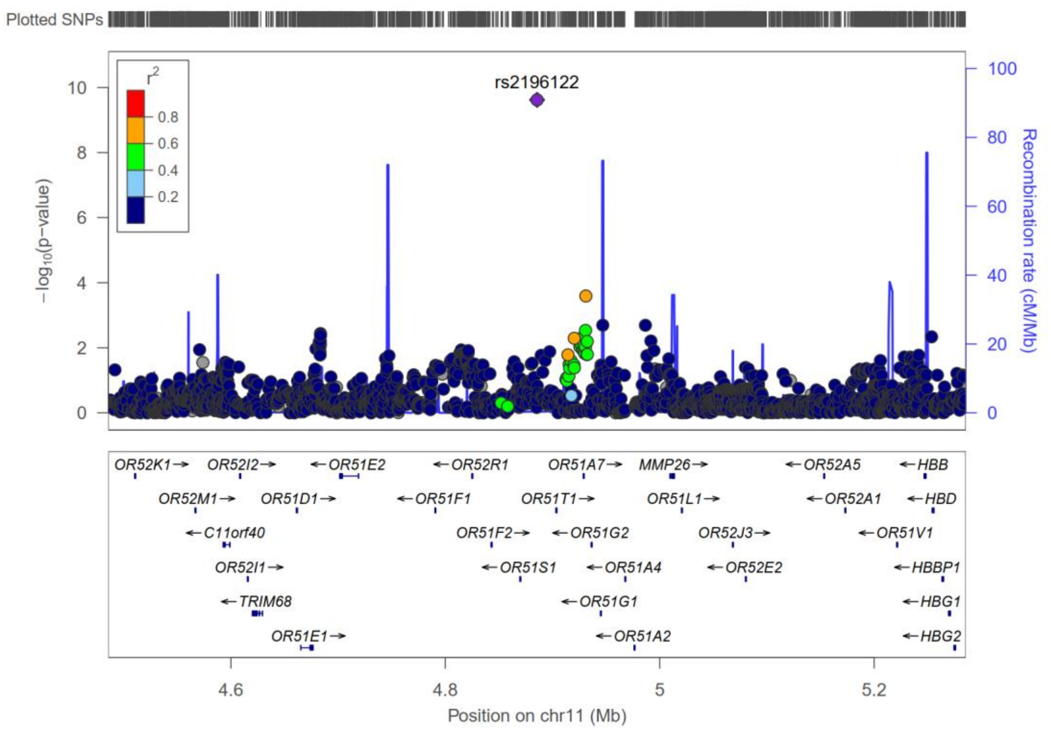

j. 11p13

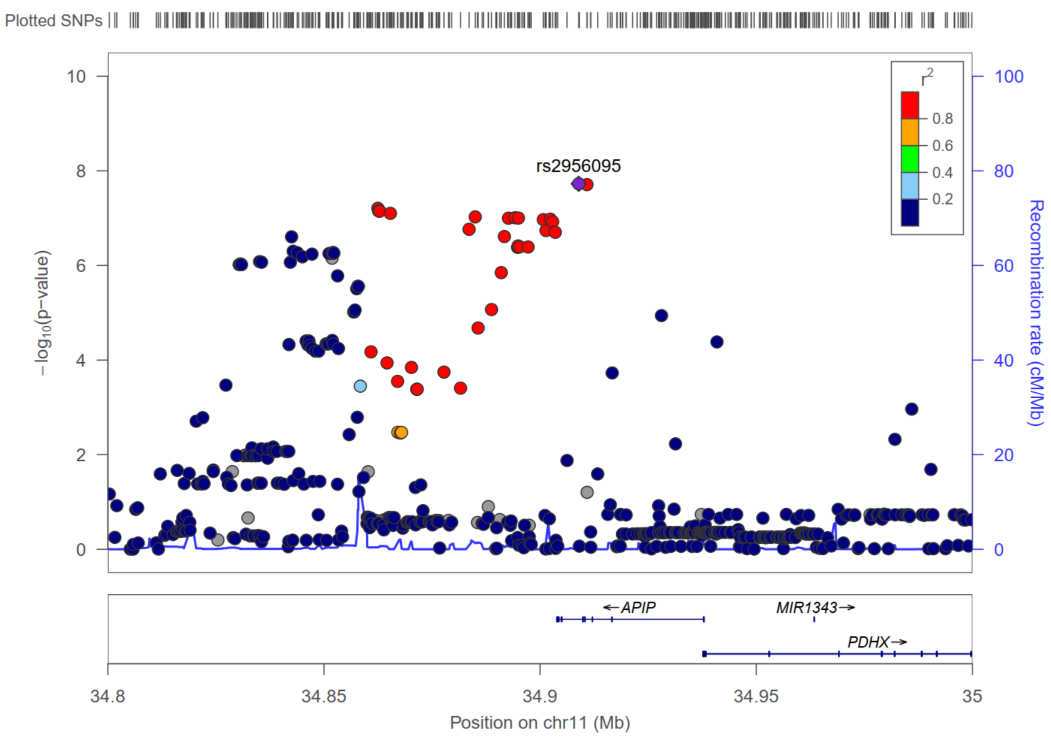


k. 22q13.31

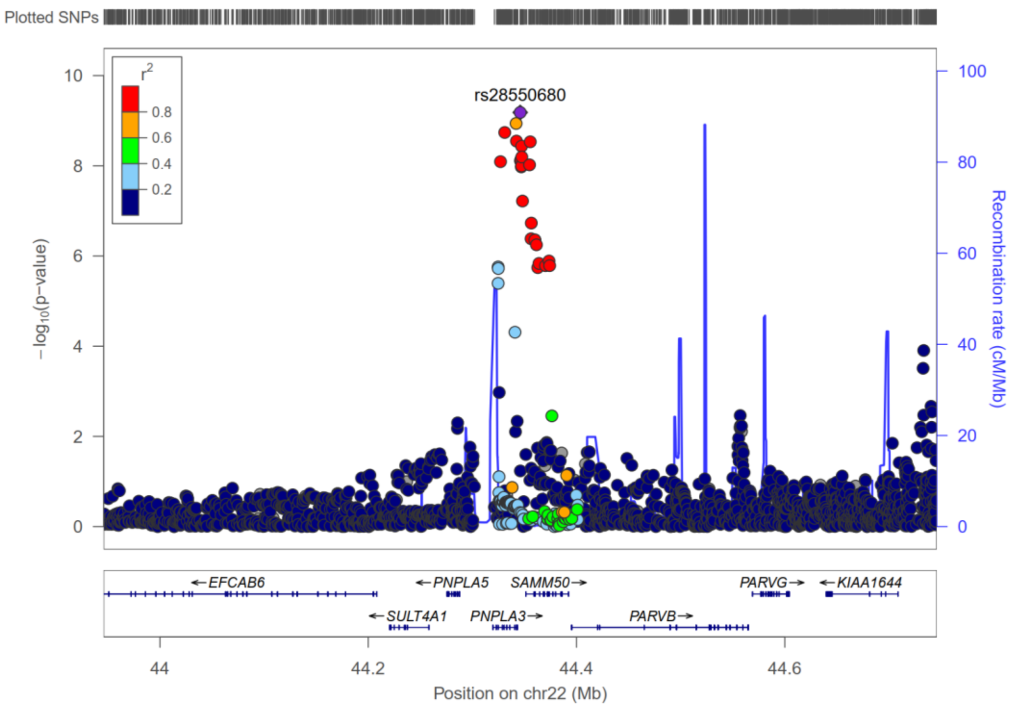

**Figure S2.** Genomic annotation of the 5q31.1 neuroblastoma risk locus (chr5:132,340,000-132,370,000, hg38) from UCSC Genome Browser. The figure displays a 30-kb genomic region containing GWAS-identified neuroblastoma risk variants (highlighting lead SNP rs2728681 and functional SNP rs2631372), overlapping with CRISPR-edited regulatory regions. Key protein-coding genes (*SLC22A4*, *SLC22A5*, *LOC553103* (*MIR3936HG*)) are annotated with their MANE Select Plus clinical transcripts. The regulatory landscape includes ENCODE-derived annotations: candidate cis-regulatory elements (cCREs), transcription factor binding clusters (912 factors across 1152 biosamples), histone modifications (H3K4me1, H3K4me3, H3K27ac in 7 cell lines), and DNase I hypersensitivity signals. RNA-seq data from 9 cell lines are shown at the bottom. Scale bar indicates 10 kb intervals.

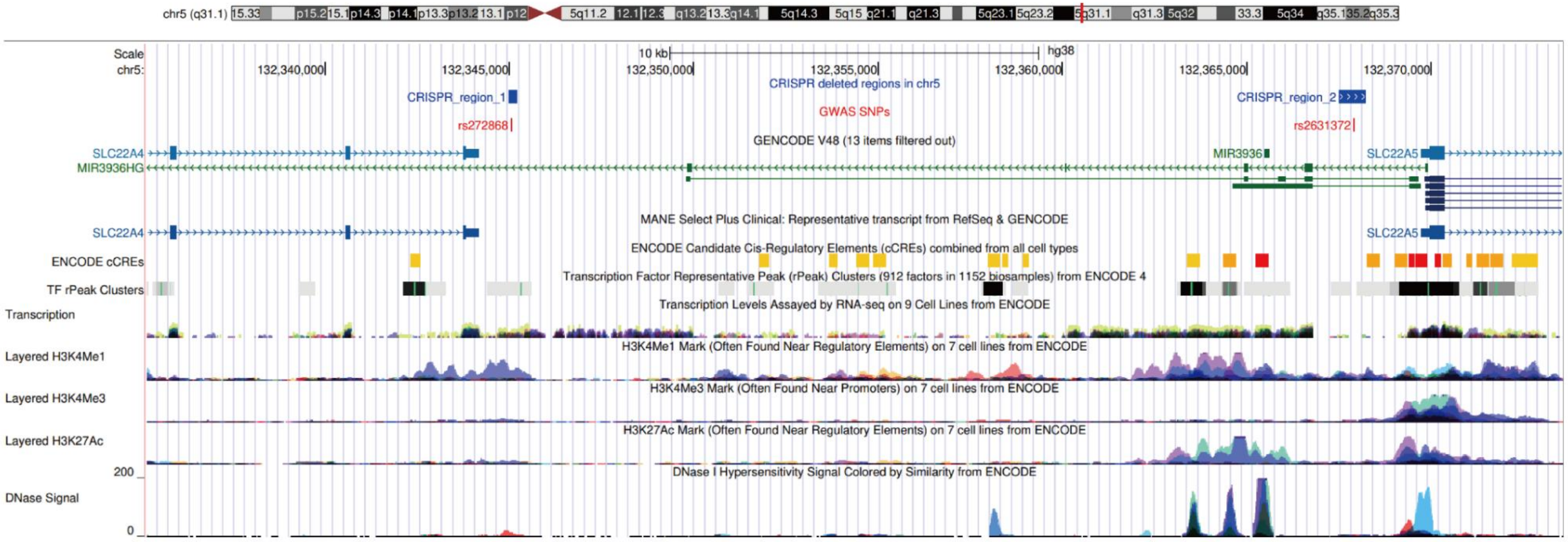

**Figure S3.** Principal component analysis (PCA) for ancestry quality control in the discovery and replication cohorts. The plot displays genetic clustering patterns of cases (red) and controls (black) against HapMap3 reference populations: African (AFR, purple), Admixed American (AMR, orange), East Asian (EAS, green), European (EUR, blue), and South Asian (SAS, brown). PCA was performed using EIGENSTRAT, with PC1 and PC2 representing the first two principal components. All study samples (cases and controls) cluster within the East Asian (EAS) reference group (green), confirming the exclusively East Asian ancestry of the analyzed cohorts. (a) Discovery cohort; (b) Replication cohort.

a. discovery cohort

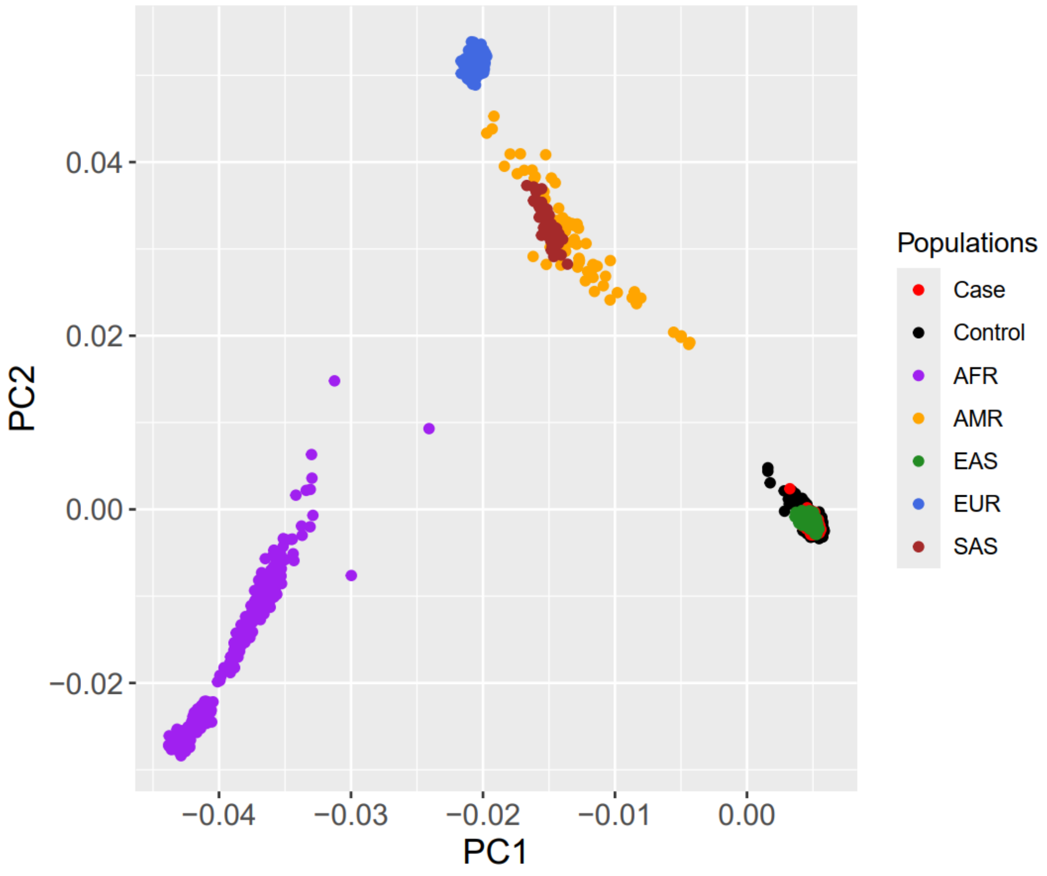

b. replication cohort

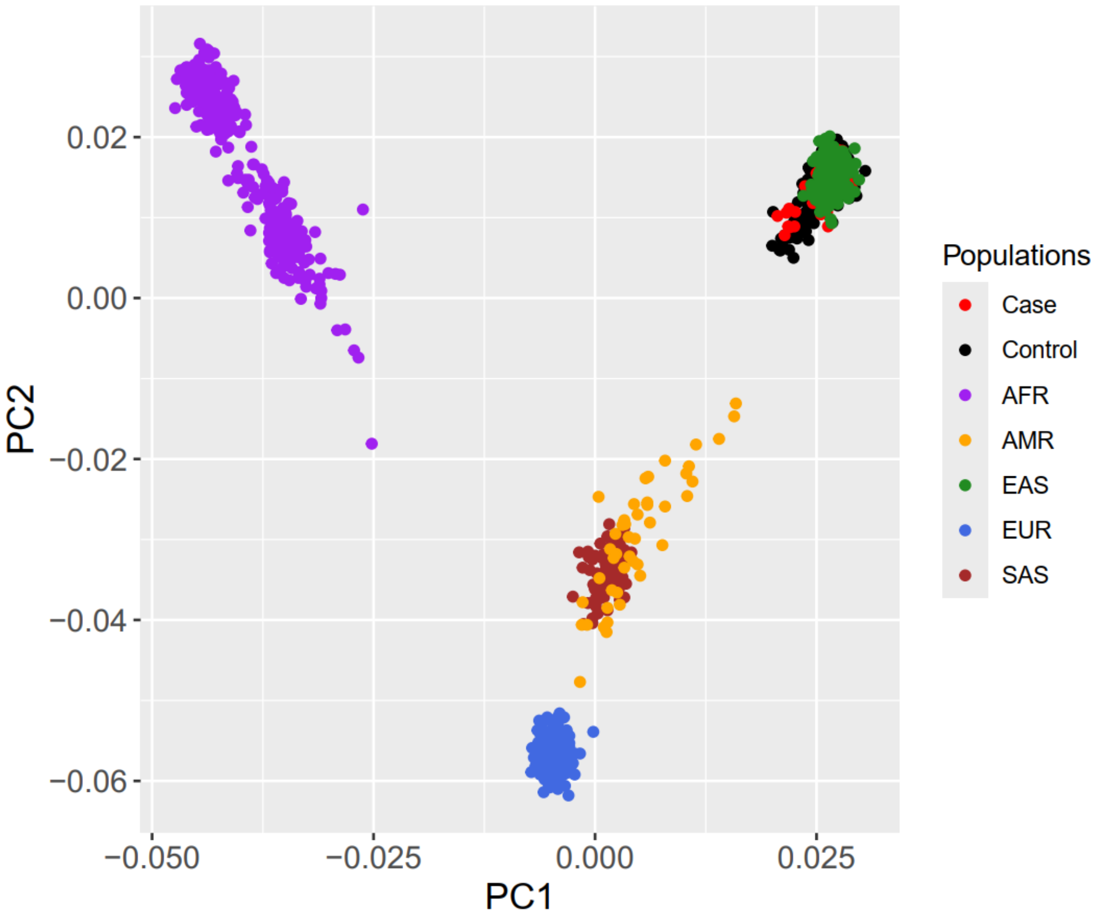

**Table S1.** Demographics of cases included in the discovery and replication stages of genome-wide association studies.

| **Dataset** | Discovery Cohort, Number(%) | Replication Cohort, Number(%) |
|---|---|---|
| **Sex** | | |
| female | 95(44.4) | 38(52.1) |
| male | 119(55.6) | 35(47.9) |
| **INSS Stage** | | |
| stage 1 | 0(0) | 12(16.4) |
| stage 2 | 0(0) | 10(13.7) |
| stage 3 | 5(2.3) | 11(15.1) |
| stage 4 | 196(90.6) | 34(46.6) |
| stage 4s | 2(0.4) | 6(8.2) |
| **Risk Group** | | |
| low | 0(0) | 19(26.0) |
| intermediate | 2(0.9) | 19(26.0) |
| high | 144(67.2) | 33(45.2) |
| **MYCN Status** | | |
| Amplified | 72(33.2) | 15(20.5) |
| non amplified | 140(64.5) | 51(60.9) |

**Table S2.** Replication of previously reported six high-risk neuroblastoma susceptibility loci in the Chinese population.

| SNP | CHR | BP | A1 | A2 | OR | SE | STAT | P | Gene | Locus | Data source |
|---|---|---|---|---|---|---|---|---|---|---|---|
| rs6435862 | 2 | 214807822 | T | G | 1.863 | 0.122 | 5.097 | 3.444E-07 | *BARD1* | 2q35 | Nat Genet_2009 |
| rs6441201 | 3 | 158460535 | A | G | 1.327 | 0.1118 | 2.533 | 0.0113 | *RSRC1/MLF1* | 3q25 | PLoS Genet_2017 |
| rs3796727 | 4 | 8611299 | A | G | 1.314 | 0.1173 | 2.326 | 0.02001 | *CPZ* | 4p16 | PLoS Genet_2017 |
| rs6939340 | 6 | 22139775 | G | A | 0.4886 | 0.1186 | -6.039 | 1.554E-09 | *CASC15* | 6p22 | N Engl J Med_2008 |
| rs7741733 | 6 | 104734964 | T | C | 0.7955 | 0.0997 | -2.296 | 0.02168 | *HACE1/LIN28B* | 6q16-q21 | PLoS Genet_2017 (rs17065417) |
| rs110419 | 11 | 8231306 | G | A | 0.6822 | 0.1031 | -3.71 | 0.000207 | *LMO1* | 11p15 | Nature_2010 |

**Table S3.** Single-Tissue eQTLs for novel loci identified in the Chinese population.

| SNP | Cytogenetic band | CHR | BP | A1 | A2 | Gene Symbol | P-Value | NES | Tissue |
|---|---|---|---|---|---|---|---|---|---|
| rs3219489 | 1p34.1 | 1 | 45331833 | G | C | *AKR1A1* | 5.40E-07 | 0.16 | Brain - Cerebellar Hemisphere |
| | | | | | | | 7.00E-06 | 0.26 | Brain - Cerebellum |
| | | | | | | | 3.60E-05 | -0.083 | Thyroid |
| | | | | | | | 0.00041 | 0.081 | Esophagus - Mucosa |
| | | | | | | *CCDC163* | 1.40E-13 | -0.44 | Skin - Not Sun Exposed (Suprapubic) |
| | | | | | | | 1.80E-13 | -0.35 | Whole Blood |
| | | | | | | | 9.90E-13 | -0.38 | Adipose - Visceral (Omentum) |
| | | | | | | | 3.50E-11 | -0.38 | Skin - Sun Exposed (Lower leg) |
| | | | | | | | 4.60E-11 | -0.36 | Thyroid |
| | | | | | | | 7.10E-11 | -0.37 | Esophagus - Mucosa |
| | | | | | | | 5.20E-10 | -0.37 | Artery - Tibial |
| | | | | | | | 1.10E-09 | -0.36 | Nerve - Tibial |
| | | | | | | | 7.40E-09 | -0.32 | Adipose - Subcutaneous |
| | | | | | | | 1.70E-08 | -0.42 | Cells - EBV-transformed lymphocytes |
| | | | | | | | 1.80E-08 | -0.44 | Esophagus - Gastroesophageal Junction |
| | | | | | | | 3.50E-08 | -0.29 | Muscle - Skeletal |
| | | | | | | | 6.00E-08 | -0.33 | Lung |
| | | | | | | | 6.60E-08 | -0.39 | Colon - Sigmoid |
| | | | | | | | 6.90E-08 | -0.53 | Small Intestine - Terminal Ileum |
| | | | | | | | 7.50E-08 | -0.34 | Cells - Cultured fibroblasts |
| | | | | | | | 1.80E-07 | -0.39 | Liver |
| | | | | | | | 2.20E-07 | -0.43 | Brain - Nucleus accumbens (basal ganglia) |
| | | | | | | | 3.20E-07 | -0.33 | Breast - Mammary Tissue |
| | | | | | | | 4.80E-07 | -0.47 | Pituitary |
| | | | | | | | 6.50E-07 | -0.31 | Colon - Transverse |

| | | | | | | | | | |
|---|---|---|---|---|---|---|---|---|---|
| | | | | | | | 7.10E-07 | -0.35 | Heart - Left Ventricle |
| | | | | | | | 7.90E-07 | 0.36 | Brain - Substantia nigra |
| | | | | | | | 4.00E-06 | -0.34 | Artery - Aorta |
| | | | | | | | 7.60E-06 | -0.38 | Prostate |
| | | | | | | | 1.40E-05 | -0.29 | Pancreas |
| | | | | | | | 1.50E-05 | -0.29 | Testis |
| | | | | | | | 3.10E-05 | -0.27 | Esophagus - Muscularis |
| | | | | | | | 3.40E-05 | -0.41 | Artery - Coronary |
| | | | | | | | 6.70E-05 | -0.44 | Brain - Cerebellum |
| | | | | | | | 0.00017 | -0.29 | Brain - Frontal Cortex (BA9) |
| | | | | | | *CCDC17* | 2.60E-06 | -0.18 | Skin - Not Sun Exposed (Suprapubic) |
| | | | | | | *EIF2B3* | 0.00012 | 0.086 | Skin - Sun Exposed (Lower leg) |
| | | | | | | *ENSG00000230896* | 6.60E-05 | -0.14 | Muscle - Skeletal |
| | | | | | | *ENSG00000234329* | 0.00031 | 0.16 | Artery - Tibial |
| | | | | | | *ENSG00000289407* | 4.80E-11 | -0.57 | Cells - EBV-transformed lymphocytes |
| | | | | | | | 2.60E-05 | -0.22 | Cells - Cultured fibroblasts |
| | | | | | | | 0.0001 | -0.26 | Colon - Transverse |
| | | | | | | | 0.00015 | -0.22 | Nerve - Tibial |
| | | | | | | *HPDL* | 0.0002 | 0.13 | Brain - Caudate (basal ganglia) |
| | | | | | | *IPP* | 1.10E-06 | -0.12 | Muscle - Skeletal |
| | | | | | | *MAST2* | 0.00045 | -0.087 | Whole Blood |
| | | | | | | *MMACHC* | 9.70E-18 | 0.25 | Artery - Tibial |
| | | | | | | | 4.30E-12 | 0.17 | Heart - Left Ventricle |
| | | | | | | | 6.80E-12 | 0.18 | Cells - Cultured fibroblasts |
| | | | | | | | 1.00E-10 | 0.18 | Adipose - Subcutaneous |
| | | | | | | | 2.50E-09 | 0.14 | Heart - Atrial Appendage |
| | | | | | | | 5.60E-09 | 0.099 | Muscle - Skeletal |
| | | | | | | | 1.80E-08 | 0.16 | Skin - Sun Exposed (Lower leg) |

| | | | | | | | | | |
|---|---|---|---|---|---|---|---|---|---|
| | | | | | | | 2.40E-08 | 0.16 | Esophagus - Muscularis |
| | | | | | | | 6.60E-08 | 0.19 | Esophagus - Mucosa |
| | | | | | | | 1.20E-05 | 0.17 | Esophagus - Gastroesophageal Junction |
| | | | | | | | 1.80E-05 | 0.19 | Artery - Aorta |
| | | | | | | | 7.90E-05 | 0.13 | Breast - Mammary Tissue |
| | | | | | | | 9.10E-05 | 0.24 | Brain - Cerebellar Hemisphere |
| | | | | | | | 0.0001 | 0.13 | Nerve - Tibial |
| | | | | | | | 0.00012 | 0.34 | Brain - Spinal cord (cervical c-1) |
| | | | | | | | 0.0002 | 0.13 | Skin - Not Sun Exposed (Suprapubic) |
| | | | | | | *MUTYH* | 8.00E-08 | -0.11 | Cells - Cultured fibroblasts |
| | | | | | | | 3.00E-07 | -0.17 | Stomach |
| | | | | | | | 6.10E-06 | -0.1 | Colon - Transverse |
| | | | | | | | 0.00011 | -0.16 | Cells - EBV-transformed lymphocytes |
| | | | | | | | 0.00011 | -0.12 | Colon - Sigmoid |
| | | | | | | | 0.00012 | -0.11 | Esophagus - Mucosa |
| | | | | | | | 0.00019 | -0.12 | Artery - Aorta |
| | | | | | | *NASP* | 5.50E-06 | -0.098 | Nerve - Tibial |
| | | | | | | *PRDX1* | 5.10E-43 | -0.27 | Thyroid |
| | | | | | | | 9.70E-18 | -0.11 | Cells - Cultured fibroblasts |
| | | | | | | | 1.30E-14 | -0.17 | Artery - Tibial |
| | | | | | | | 1.60E-11 | -0.17 | Adipose - Subcutaneous |
| | | | | | | | 3.10E-10 | -0.17 | Artery - Aorta |
| | | | | | | | 1.20E-09 | -0.14 | Lung |
| | | | | | | | 2.20E-09 | 0.11 | Testis |
| | | | | | | | 5.10E-07 | 0.17 | Brain - Caudate (basal ganglia) |
| | | | | | | | 7.20E-07 | 0.2 | Brain - Putamen (basal ganglia) |
| | | | | | | | 1.50E-05 | -0.091 | Adipose - Visceral (Omentum) |

| | | | | | | | | | |
|---|---|---|---|---|---|---|---|---|---|
| | | | | | | | 3.50E-05 | -0.11 | Liver |
| | | | | | | | 4.30E-05 | -0.081 | Nerve - Tibial |
| | | | | | | | 6.30E-05 | -0.089 | Heart - Left Ventricle |
| | | | | | | | 6.80E-05 | -0.13 | Artery - Coronary |
| | | | | | | | 8.50E-05 | 0.14 | Brain - Frontal Cortex (BA9) |
| | | | | | | | 0.00021 | -0.12 | Pancreas |
| | | | | | | *TESK2* | 7.70E-18 | 0.4 | Brain - Caudate (basal ganglia) |
| | | | | | | | 2.20E-13 | 0.37 | Brain - Putamen (basal ganglia) |
| | | | | | | | 3.40E-11 | 0.18 | Whole Blood |
| | | | | | | | 1.00E-10 | 0.19 | Testis |
| | | | | | | | 1.60E-09 | 0.23 | Brain - Frontal Cortex (BA9) |
| | | | | | | | 2.30E-09 | 0.28 | Brain - Cortex |
| | | | | | | | 7.20E-08 | 0.28 | Spleen |
| | | | | | | | 4.10E-07 | 0.15 | Cells - Cultured fibroblasts |
| | | | | | | | 1.60E-06 | 0.11 | Esophagus - Mucosa |
| | | | | | | | 1.60E-06 | -0.2 | Brain - Spinal cord (cervical c-1) |
| | | | | | | | 2.00E-06 | 0.16 | Brain - Hypothalamus |
| | | | | | | | 2.10E-05 | 0.087 | Skin - Not Sun Exposed (Suprapubic) |
| | | | | | | | 5.90E-05 | 0.21 | Brain - Substantia nigra |
| | | | | | | *TMEM69* | 0.00027 | 0.083 | Esophagus - Mucosa |
| | | | | | | *TOE1* | 0.00044 | 0.075 | Thyroid |
| | | | | | | *UROD* | 1.30E-05 | -0.11 | Nerve - Tibial |
| | | | | | | *ZSWIM5* | 1.90E-08 | 0.13 | Thyroid |
| | | | | | | | 2.90E-07 | 0.18 | Adipose - Subcutaneous |
| | | | | | | | 9.80E-07 | 0.16 | Nerve - Tibial |
| | | | | | | | 3.90E-06 | 0.18 | Heart - Atrial Appendage |
| | | | | | | | 2.10E-05 | 0.13 | Esophagus - Muscularis |

| | | | | | | | | | |
|---|---|---|---|---|---|---|---|---|---|
| | | | | | | | 8.70E-05 | 0.13 | Artery - Tibial |
| | | | | | | | 9.20E-05 | -0.11 | Testis |
| | | | | | | | 0.00023 | 0.13 | Artery - Aorta |
| rs2165738 | 2p23.3 | 2 | 24469940 | C | G | *ADCY3* | 7.20E-19 | -0.4 | Whole Blood |
| | | | | | | | 1.20E-06 | -0.15 | Artery - Tibial |
| | | | | | | | 3.90E-06 | 0.11 | Nerve - Tibial |
| | | | | | | | 1.10E-05 | 0.093 | Skin - Not Sun Exposed (Suprapubic) |
| | | | | | | *CENPO* | 6.80E-13 | -0.29 | Whole Blood |
| | | | | | | | 2.30E-09 | -0.19 | Artery - Tibial |
| | | | | | | | 3.30E-08 | -0.22 | Lung |
| | | | | | | *DNAJC27* | 1.20E-11 | -0.24 | Whole Blood |
| | | | | | | *DNAJC27-AS1* | 3.30E-07 | 0.14 | Nerve - Tibial |
| | | | | | | *ENSG00000271936* | 7.10E-06 | -0.24 | Lung |
| | | | | | | *NCOA1* | 1.30E-06 | -0.17 | Heart - Atrial Appendage |
| | | | | | | | 1.60E-06 | -0.13 | Lung |
| | | | | | | | 2.60E-05 | -0.094 | Artery - Tibial |
| | | | | | | | 8.70E-05 | -0.13 | Nerve - Tibial |
| rs10024137 | 4q13.2 | 4 | 68667043 | A | G | *UGT2B4* | 0.00014 | 0.21 | Lung |
| rs3733631 | 4q24 | 4 | 103719946 | C | G | *BDH2* | 2.10E-05 | 0.13 | Adipose - Subcutaneous |
| | | | | | | *TACR3* | 2.40E-15 | -0.39 | Lung |
| | | | | | | | 5.00E-06 | -0.47 | Kidney - Cortex |
| | | | | | | | 7.00E-06 | 0.34 | Brain - Frontal Cortex (BA9) |
| rs272868 | 5q31.1 | 5 | 132345058 | C | G | *ENSG00000224431* | 1.10E-09 | 0.22 | Thyroid |
| | | | | | | *IRF1* | 1.50E-07 | -0.18 | Muscle - Skeletal |
| | | | | | | *IRF1-AS1* | 0.00028 | -0.12 | Muscle - Skeletal |
| | | | | | | *KIF3A* | 7.40E-05 | 0.11 | Nerve - Tibial |
| | | | | | | *LOC553103 (MIR3936HG)* | 1.70E-68 | 0.49 | Thyroid |
| | | | | | | | 4.90E-57 | 0.42 | Muscle - Skeletal |

| | | | | | | | | | |
|---|---|---|---|---|---|---|---|---|---|
| | | | | | | | 5.70E-37 | 0.49 | Heart - Left Ventricle |
| | | | | | | | 1.10E-26 | 0.4 | Stomach |
| | | | | | | | 5.10E-24 | 0.44 | Heart - Atrial Appendage |
| | | | | | | | 4.00E-23 | 0.66 | Brain - Nucleus accumbens (basal ganglia) |
| | | | | | | | 3.00E-20 | 0.62 | Brain - Caudate (basal ganglia) |
| | | | | | | | 7.50E-19 | 0.57 | Spleen |
| | | | | | | | 2.70E-18 | 0.37 | Pituitary |
| | | | | | | | 5.90E-18 | 0.42 | Adrenal Gland |
| | | | | | | | 2.00E-17 | 0.71 | Brain - Putamen (basal ganglia) |
| | | | | | | | 4.80E-15 | 0.24 | Esophagus - Mucosa |
| | | | | | | | 5.70E-15 | 0.57 | Brain - Frontal Cortex (BA9) |
| | | | | | | | 9.70E-15 | 0.28 | Prostate |
| | | | | | | | 1.70E-14 | 0.21 | Adipose - Visceral (Omentum) |
| | | | | | | | 9.30E-14 | 0.27 | Lung |
| | | | | | | | 9.90E-13 | 0.57 | Brain - Cortex |
| | | | | | | | 3.70E-12 | 0.2 | Skin - Sun Exposed (Lower leg) |
| | | | | | | | 6.70E-12 | 0.27 | Whole Blood |
| | | | | | | | 9.20E-12 | 0.5 | Brain - Substantia nigra |
| | | | | | | | 1.50E-11 | 0.57 | Brain - Anterior qcingulate cortex (BA24) |
| | | | | | | | 1.80E-11 | 0.42 | Brain - Hypothalamus |
| | | | | | | | 4.40E-11 | 0.46 | Cells - EBV-transformed lymphocytes |
| | | | | | | | 6.20E-11 | 0.3 | Brain - Cerebellar Hemisphere |
| | | | | | | | 3.80E-10 | 0.3 | Small Intestine - Terminal Ileum |
| | | | | | | | 4.20E-10 | 0.16 | Colon - Transverse |
| | | | | | | | 2.20E-09 | 0.15 | Artery - Aorta |
| | | | | | | | 3.00E-09 | 0.2 | Adipose - Subcutaneous |
| | | | | | | | 1.20E-08 | 0.41 | Brain - Hippocampus |
| | | | | | | | 3.50E-08 | 0.21 | Liver |
| | | | | | | | 3.90E-08 | 0.17 | Skin - Not Sun Exposed (Suprapubic) |

| | | | | | | | | | |
|---|---|---|---|---|---|---|---|---|---|
| | | | | | | | 4.40E-07 | 0.27 | Testis |
| | | | | | | | 1.10E-05 | 0.35 | Brain - Amygdala |
| | | | | | | | 1.40E-05 | 0.16 | Cells - Cultured fibroblasts |
| | | | | | | | 3.30E-05 | 0.1 | Breast - Mammary Tissue |
| | | | | | | | 0.00011 | 0.15 | Esophagus - Muscularis |
| | | | | | | *P4HA2* | 5.10E-26 | 0.35 | Thyroid |
| | | | | | | | 1.10E-19 | 0.23 | Nerve - Tibial |
| | | | | | | | 3.60E-15 | 0.3 | Esophagus - Mucosa |
| | | | | | | | 7.00E-12 | 0.25 | Artery - Aorta |
| | | | | | | | 2.40E-10 | 0.15 | Artery - Tibial |
| | | | | | | | 4.50E-08 | 0.085 | Cells - Cultured fibroblasts |
| | | | | | | | 2.20E-07 | 0.2 | Testis |
| | | | | | | | 3.50E-07 | 0.24 | Brain - Cerebellar Hemisphere |
| | | | | | | | 4.80E-06 | 0.21 | Cells - EBV-transformed lymphocytes |
| | | | | | | | 5.30E-05 | 0.13 | Esophagus - Muscularis |
| | | | | | | | 5.80E-05 | 0.15 | Esophagus - Gastroesophageal Junction |
| | | | | | | | 6.70E-05 | -0.13 | Brain - Hypothalamus |
| | | | | | | | 0.0002 | 0.19 | Brain - Cerebellum |
| | | | | | | *PDLIM4* | 5.90E-22 | -0.25 | Skin - Sun Exposed (Lower leg) |
| | | | | | | | 1.50E-17 | -0.13 | Cells - Cultured fibroblasts |
| | | | | | | | 3.40E-17 | -0.23 | Skin - Not Sun Exposed (Suprapubic) |
| | | | | | | | 6.70E-17 | -0.32 | Brain - Frontal Cortex (BA9) |
| | | | | | | | 7.90E-16 | -0.16 | Adipose - Subcutaneous |
| | | | | | | | 2.80E-12 | -0.26 | Heart - Left Ventricle |
| | | | | | | | 1.30E-11 | -0.28 | Brain - Anterior qcingulate cortex (BA24) |
| | | | | | | | 8.80E-11 | -0.12 | Breast - Mammary Tissue |
| | | | | | | | 1.40E-10 | -0.37 | Cells - EBV-transformed lymphocytes |
| | | | | | | | 9.50E-10 | -0.25 | Brain - Cortex |
| | | | | | | | 1.10E-07 | -0.13 | Thyroid |

| | | | | | | | | | |
|---|---|---|---|---|---|---|---|---|---|
| | | | | | | | 2.30E-07 | -0.34 | Brain - Amygdala |
| | | | | | | | 4.50E-07 | -0.11 | Adipose - Visceral (Omentum) |
| | | | | | | | 5.90E-07 | -0.11 | Muscle - Skeletal |
| | | | | | | | 7.90E-07 | -0.23 | Brain - Hypothalamus |
| | | | | | | | 8.20E-07 | -0.24 | Pancreas |
| | | | | | | | 1.20E-05 | -0.18 | Brain - Hippocampus |
| | | | | | | | 0.00011 | -0.12 | Pituitary |
| | | | | | | *RAD50* | 7.30E-05 | 0.1 | Skin - Not Sun Exposed (Suprapubic) |
| | | | | | | | 0.00032 | 0.094 | Nerve - Tibial |
| | | | | | | *RAPGEF6* | 1.80E-05 | -0.19 | Testis |
| | | | | | | *SLC22A4* | 1.20E-17 | -0.22 | Muscle - Skeletal |
| | | | | | | | 4.90E-11 | -0.19 | Skin - Sun Exposed (Lower leg) |
| | | | | | | | 2.60E-08 | 0.2 | Artery - Aorta |
| | | | | | | | 8.20E-08 | 0.42 | Brain - Cerebellum |
| | | | | | | | 9.30E-06 | -0.067 | Whole Blood |
| | | | | | | | 9.70E-06 | 0.19 | Thyroid |
| | | | | | | | 7.90E-05 | -0.13 | Spleen |
| | | | | | | *SLC22A5* | 3.60E-45 | -0.42 | Cells - Cultured fibroblasts |
| | | | | | | | 1.00E-25 | -0.31 | Skin - Sun Exposed (Lower leg) |
| | | | | | | | 1.10E-17 | -0.27 | Skin - Not Sun Exposed (Suprapubic) |
| | | | | | | | 6.90E-15 | 0.31 | Brain - Cerebellum |
| | | | | | | | 2.20E-14 | 0.28 | Brain - Cerebellar Hemisphere |
| | | | | | | | 3.00E-13 | -0.31 | Adipose - Subcutaneous |
| | | | | | | | 9.00E-13 | -0.28 | Lung |
| | | | | | | | 5.60E-12 | -0.22 | Esophagus - Mucosa |
| | | | | | | | 2.00E-09 | -0.17 | Breast - Mammary Tissue |
| | | | | | | | 4.40E-09 | -0.15 | Colon - Transverse |
| | | | | | | | 2.50E-07 | -0.15 | Stomach |

| | | | | | | | | | |
|---|---|---|---|---|---|---|---|---|---|
| | | | | | | | 2.60E-07 | -0.1 | Muscle - Skeletal |
| | | | | | | | 8.20E-06 | -0.33 | Brain - Spinal cord (cervical c-1) |
| | | | | | | | 1.80E-05 | -0.15 | Whole Blood |
| | | | | | | | 3.90E-05 | -0.32 | Spleen |
| | | | | | | *TH2LCRR* | 0.0002 | 0.19 | Skin - Sun Exposed (Lower leg) |
| rs2160860 | 8p11.21 | 8 | 39965626 | A | T | *IDO1* | 1.40E-12 | -0.28 | Adipose - Visceral (Omentum) |
| | | | | | | | 6.50E-09 | -0.23 | Colon - Transverse |
| | | | | | | | 9.40E-09 | -0.26 | Breast - Mammary Tissue |
| | | | | | | | 7.50E-07 | -0.2 | Lung |
| | | | | | | | 1.10E-06 | -0.18 | Adipose - Subcutaneous |
| | | | | | | | 4.70E-06 | -0.22 | Stomach |
| | | | | | | | 5.20E-06 | -0.17 | Skin - Sun Exposed (Lower leg) |
| | | | | | | | 1.50E-05 | -0.24 | Colon - Sigmoid |
| | | | | | | *IDO2* | 1.90E-07 | -0.17 | Thyroid |
| | | | | | | | 2.60E-06 | -0.21 | Lung |
| | | | | | | | 8.20E-06 | -0.24 | Adipose - Visceral (Omentum) |
| rs2956095 | 11p13 | 11 | 34887382 | C | T | *APIP* | 2.20E-17 | -0.51 | Cells - EBV-transformed lymphocytes |
| | | | | | | | 1.60E-09 | -0.17 | Cells - Cultured fibroblasts |
| | | | | | | | 5.40E-07 | -0.18 | Muscle - Skeletal |
| | | | | | | | 3.60E-06 | -0.15 | Whole Blood |
| | | | | | | | 8.60E-06 | 0.41 | Brain - Spinal cord (cervical c-1) |
| | | | | | | | 1.20E-05 | -0.26 | Esophagus - Mucosa |
| | | | | | | | 5.80E-05 | -0.28 | Testis |
| | | | | | | *PDHX* | 7.70E-18 | -0.32 | Esophagus - Mucosa |
| | | | | | | | 1.10E-17 | -0.28 | Skin - Not Sun Exposed (Suprapubic) |
| | | | | | | | 1.20E-17 | -0.31 | Skin - Sun Exposed (Lower leg) |
| | | | | | | | 1.10E-09 | -0.4 | Cells - EBV-transformed lymphocytes |
| | | | | | | | 2.80E-07 | 0.098 | Muscle - Skeletal |
| | | | | | | | 1.10E-05 | -0.1 | Heart - Atrial Appendage |

| | | | | | | | | | |
|---|---|---|---|---|---|---|---|---|---|
| | | | | | | | 2.30E-05 | 0.14 | Artery - Tibial |
| | | | | | | | 0.00012 | -0.15 | Nerve - Tibial |
| rs28550680 | 22q13.31 | 22 | 43950046 | T | C | *PNPLA3* | 5.30E-17 | 0.26 | Skin - Not Sun Exposed (Suprapubic) |
| | | | | | | | 7.70E-14 | 0.2 | Skin - Sun Exposed (Lower leg) |
| | | | | | | | 1.80E-07 | 0.2 | Cells - Cultured fibroblasts |
| | | | | | | *SAMM50* | 7.40E-07 | 0.11 | Whole Blood |
| | | | | | | | 2.50E-05 | 0.21 | Brain - Hypothalamus |
| | | | | | | | 3.60E-05 | 0.31 | Brain - Hippocampus |

**Table S4.** Single-Cell eQTLs for novel loci identified in the Chinese population.

| SNP | Cytogenetic band | CHR | BP | A1 | A2 | geneName | QTLtype | cellTypeName | pValue | beta | se | study |
|---|---|---|---|---|---|---|---|---|---|---|---|---|
| rs3219489 | 1p34.1 | 1 | 45331833 | G | C | *AKR1A1* | Cell-type-specific eQTL | CD4+ T Cell | 4.07E-05 | -0.62 | 0.15 | Schmiedel2022SI |
| | | | | | | *CCDC163* | Cell-type-specific eQTL | Naive T Cell | 2.57E-05 | -0.71 | 0.16 | Soskic2022NG |
| | | | | | | | Cell-type-specific eQTL | Naive T Cell | 2.27E-04 | -0.67 | 0.17 | Soskic2022NG |
| | | | | | | | Cell-type-specific eQTL | Naive T Cell | 1.11E-04 | -0.68 | 0.17 | Soskic2022NG |
| | | | | | | | Cell-type-specific eQTL | CD4+ Central Memory T | 4.02E-04 | -0.63 | 0.17 | Soskic2022NG |
| | | | | | | | Cell-type-specific eQTL | CD4+ Naive T | 1.20E-03 | -0.65 | 0.19 | Soskic2022NG |
| | | | | | | *IPP* | Cell-type-specific eQTL | Serotonergic-like Neurons | 5.42E-05 | -0.33 | 0.08 | Jerber2021NG |
| | | | | | | *MUTYH* | Cell-type-specific eQTL | CD4+ Naive T | 2.57E-04 | -0.61 | 0.16 | Soskic2022NG |
| | | | | | | | Cell-type-specific eQTL | CD4+ Naive T | 1.02E-03 | -0.43 | 0.13 | Soskic2022NG |
| | | | | | | | Cell-type-specific eQTL | Serotonergic-like Neurons | 4.79E-06 | -0.34 | 0.08 | Jerber2021NG |
| | | | | | | | Response eQTL | Serotonergic-like Neurons | 1.30E-04 | -0.20 | 0.05 | Jerber2021NG |
| | | | | | | | Cell-type-specific eQTL | Ependymal-like 1 | 1.96E-06 | -0.36 | 0.08 | Jerber2021NG |
| | | | | | | | Response eQTL | Ependymal-like 1 | 6.38E-05 | -0.22 | 0.06 | Jerber2021NG |

| | | | | | | | | | | | |
|---|---|---|---|---|---|---|---|---|---|---|---|
| | | | | | | | Cell-type-specific eQTL | Ependymal-like 1 | 3.17E-04 | -0.23 | 0.06 | Jerber2021NG |
| | | | | | | | Cell-type-specific eQTL | Proliferating Floor Plate Progenitors | 1.15E-08 | -0.41 | 0.07 | Jerber2021NG |
| | | | | | | | Cell-type-specific eQTL | Floor Plate Progenitors | 5.67E-06 | -0.35 | 0.08 | Jerber2021NG |
| | | | | | | *PRDX1* | Cell-type-specific eQTL | Regulatory T (Treg) | 2.91E-06 | -0.59 | 0.12 | Soskic2022NG |
| | | | | | | | Cell-type-specific eQTL | Naive T Cell | 2.47E-05 | -0.46 | 0.10 | Soskic2022NG |
| | | | | | | | Cell-type-specific eQTL | CD4+ Naive T | 6.35E-04 | -0.38 | 0.11 | Soskic2022NG |
| | | | | | | | Cell-type-specific eQTL | Dopaminergic Neurons | 2.97E-04 | -0.25 | 0.07 | Jerber2021NG |
| | | | | | | | Cell-type-specific eQTL | Dopaminergic Neurons | 3.63E-05 | -0.16 | 0.04 | Jerber2021NG |
| | | | | | | *TESK2* | Cell-type-specific eQTL | Monocyte (Mono) | 3.40E-05 | 0.64 | 0.15 | Schmiedel2022SI |
| | | | | | | | Cell-type-specific eQTL | Naive B Cell | 1.70E-16 | 1.13 | 0.14 | Schmiedel2022SI |
| rs2165738 | 2p23.3 | 2 | 24469940 | C | G | *ADCY3* | Cell-type-specific eQTL | Astrocytes | 7.92E-05 | -0.32 | 0.08 | Bryois2022NN |
| rs272868 | 5q31.1 | 5 | 1.32E+08 | C | G | *LOC553103 (AC034220.3)* | Cell-type-specific eQTL | CD8+ Naive T | 1.79E-05 | 0.60 | 0.14 | Schmiedel2022SI |
| | | | | | | | Cell-type-specific eQTL | Naive B Cell | 5.62E-10 | 0.83 | 0.13 | Schmiedel2022SI |
| | | | | | | | Cell-type-specific eQTL | CD8+ T Cell | 1.91E-07 | 0.27 | 0.05 | Aquino2023Nature |

| | | | | | | | | | | | | |
|---|---|---|---|---|---|---|---|---|---|---|---|---|
| | | | | | | | Cell-type-specific eQTL | CD8+ T Cell | 1.91E-07 | 0.27 | 0.05 | Aquino2023Nature |
| | | | | | | | Cell-type-specific eQTL | CD8+ T Cell | 5.34E-04 | 0.22 | 0.06 | Aquino2023Nature |
| | | | | | | | Cell-type-specific eQTL | CD8+ T Cell | 5.34E-04 | 0.22 | 0.06 | Aquino2023Nature |
| | | | | | | | Cell-type-specific eQTL | CD8+ T Cell | 1.90E-04 | 0.21 | 0.05 | Aquino2023Nature |
| | | | | | | | Cell-type-specific eQTL | CD8+ T Cell | 1.90E-04 | 0.21 | 0.05 | Aquino2023Nature |
| | | | | | | | Cell-type-specific eQTL | CD4+ T Cell | 9.16E-07 | 0.22 | 0.04 | Aquino2023Nature |
| | | | | | | | Cell-type-specific eQTL | CD4+ T Cell | 9.16E-07 | 0.22 | 0.04 | Aquino2023Nature |
| | | | | | | | Cell-type-specific eQTL | CD4+ T Cell | 4.34E-06 | 0.22 | 0.05 | Aquino2023Nature |
| | | | | | | | Cell-type-specific eQTL | CD4+ T Cell | 4.34E-06 | 0.22 | 0.05 | Aquino2023Nature |
| | | | | | | | Cell-type-specific eQTL | CD4+ T Cell | 7.58E-06 | 0.22 | 0.05 | Aquino2023Nature |
| | | | | | | | Cell-type-specific eQTL | CD4+ T Cell | 7.58E-06 | 0.22 | 0.05 | Aquino2023Nature |
| | | | | | | | Cell-type-specific eQTL | B Cell | 2.11E-04 | 0.34 | 0.09 | Aquino2023Nature |
| | | | | | | | Cell-type-specific eQTL | B Cell | 2.11E-04 | 0.34 | 0.09 | Aquino2023Nature |
| | | | | | | | Cell-type-specific eQTL | CD8+ Naive T | 6.10E-05 | 0.37 | 0.09 | Aquino2023Nature |

| | | | | | | | | | | | | |
|---|---|---|---|---|---|---|---|---|---|---|---|---|
| | | | | | | | Cell-type-specific eQTL | CD8+ Naive T | 6.10E-05 | 0.37 | 0.09 | Aquino2023Nature |
| | | | | | | | Cell-type-specific eQTL | CD4+ Naive T | 6.11E-07 | 0.35 | 0.07 | Aquino2023Nature |
| | | | | | | | Cell-type-specific eQTL | CD4+ Naive T | 6.11E-07 | 0.35 | 0.07 | Aquino2023Nature |
| | | | | | | | Cell-type-specific eQTL | CD4+ Naive T | 1.68E-06 | 0.30 | 0.06 | Aquino2023Nature |
| | | | | | | | Cell-type-specific eQTL | CD4+ Naive T | 1.68E-06 | 0.30 | 0.06 | Aquino2023Nature |
| | | | | | | | Cell-type-specific eQTL | CD4+ Naive T | 1.39E-05 | 0.30 | 0.07 | Aquino2023Nature |
| | | | | | | | Cell-type-specific eQTL | CD4+ Naive T | 1.39E-05 | 0.30 | 0.07 | Aquino2023Nature |
| | | | | | | *P4HA2* | Cell-type-specific eQTL | CD4+ Effector Memory T | 2.03E-04 | -0.47 | 0.12 | Soskic2022NG |
| | | | | | | | Cell-type-specific eQTL | CD4 Memory | 4.50E-05 | -0.46 | 0.11 | Soskic2022NG |
| | | | | | | *PDLIM4* | Cell-type-specific eQTL | CD4+ Effector Memory T | 4.54E-09 | 0.95 | 0.14 | Soskic2022NG |
| | | | | | | | Cell-type-specific eQTL | CD4+ Effector Memory T | 1.67E-05 | 0.65 | 0.14 | Soskic2022NG |
| | | | | | | | Response eQTL | Dendritic Cell (DC) | 3.43E-07 | 5.10 | 1.00 | Oelen2022NC |
| | | | | | | | | CD4+ T Cell | 9.05E-05 | 3.91 | 1.00 | Oelen2022NC |
| | | | | | | *SEP8* | Cell-type-specific eQTL | Serotonergic-like Neurons | 2.34E-04 | 0.21 | 0.06 | Jerber2021NG |
| | | | | | | *SLC22A4* | Cell-type-specific eQTL | CD4+ T Cell | 7.24E-07 | 0.71 | 0.14 | Schmiedel2022SI |

| | | | | | | | | | | | | |
|---|---|---|---|---|---|---|---|---|---|---|---|---|
| | | | | | | | Cell-type-specific eQTL | CD8+ Naive T | 2.99E-07 | 0.73 | 0.14 | Schmiedel2022SI |
| | | | | | | | Cell-type-specific eQTL | Naive B Cell | 3.79E-06 | 0.68 | 0.15 | Schmiedel2022SI |
| | | | | | | *SLC22A5* | Cell-type-specific eQTL | Monocyte (Mono) | 1.31E-05 | -0.61 | 0.14 | Schmiedel2022SI |
| | | | | | | | Cell-type-specific eQTL | CD8+ Naive T | 4.42E-06 | -0.60 | 0.13 | Schmiedel2022SI |
| | | | | | | | Cell-type-specific eQTL | CD4+ Naive T | 2.22E-05 | -0.55 | 0.13 | Schmiedel2022SI |
| | | | | | | | Cell-type-specific eQTL | CD8+ T Cell | 8.57E-05 | 3.93 | 1.00 | Oelen2022NC |
| | | | | | | | Cell-type-specific eQTL | CD4+ T Cell | 8.53E-05 | 3.93 | 1.00 | Oelen2022NC |
| | | | | | | | Cell-type-specific eQTL | Ependymal-like 1 | 6.99E-05 | -0.22 | 0.06 | Jerber2021NG |
| | | | | | | | Response eQTL | Ependymal-like 1 | 9.46E-07 | -0.38 | 0.08 | Jerber2021NG |
| rs60587781 | 5p15.1 | 5 | 17223168 | T | C | *BASP1* | Response eQTL | Plasmacytoid Dendritic Cell | 9.62E-05 | 0.11 | 0.03 | Perez2022Science |
| rs2956095 | 11p13 | 11 | 34887382 | C | T | *APIP* | Cell-type-specific eQTL | Naive T Cell | 4.03E-05 | 0.47 | 0.11 | Soskic2022NG |
| | | | | | | | Cell-type-specific eQTL | Naive T Cell | 4.18E-05 | 0.67 | 0.15 | Soskic2022NG |
| | | | | | | | Cell-type-specific eQTL | Naive T Cell | 1.99E-10 | 0.75 | 0.10 | Soskic2022NG |
| | | | | | | | Cell-type-specific eQTL | CD4+ Effector Memory T | 2.08E-06 | 0.38 | 0.07 | Soskic2022NG |
| | | | | | | | Cell-type-specific eQTL | CD4+ Central Memory T | 2.91E-05 | 0.66 | 0.15 | Soskic2022NG |

| | | | | | | | | | | | | |
|---|---|---|---|---|---|---|---|---|---|---|---|---|
| | | | | | | | Cell-type-specific eQTL | CD4+ Central Memory T | 7.63E-10 | 0.79 | 0.11 | Soskic2022NG |
| | | | | | | | Cell-type-specific eQTL | CD4+ Naive T | 2.07E-07 | 0.65 | 0.11 | Soskic2022NG |
| | | | | | | | Cell-type-specific eQTL | CD4+ Naive T | 5.47E-05 | 0.57 | 0.13 | Soskic2022NG |
| | | | | | | | Cell-type-specific eQTL | CD4+ Naive T | 2.06E-09 | 0.68 | 0.10 | Soskic2022NG |
| | | | | | | | Cell-type-specific eQTL | CD4 Memory | 1.92E-07 | 0.85 | 0.15 | Soskic2022NG |
| | | | | | | | Cell-type-specific eQTL | CD4 Memory | 2.32E-04 | 0.21 | 0.05 | Soskic2022NG |
| | | | | | | | Cell-type-specific eQTL | CD4 Memory | 1.62E-11 | 0.88 | 0.11 | Soskic2022NG |
| | | | | | | | Cell-type-specific eQTL | CD8+ Naive T | 8.27E-06 | -0.98 | 0.22 | Schmiedel2022SI |
| | | | | | | | Cell-type-specific eQTL | CD4+ T Cell | 8.52E-06 | 0.78 | 0.18 | Randolph2021Science |
| | | | | | | | Response eQTL | CD4+ T Cell | 1.16E-04 | 3.85 | 1.00 | Oelen2022NC |
| | | | | | | | Cell-type-specific eQTL | Ependymal-like 1 | 6.09E-06 | -0.37 | 0.08 | Jerber2021NG |
| | | | | | | | Cell-type-specific eQTL | Floor Plate Progenitors | 8.92E-04 | -0.25 | 0.08 | Jerber2021NG |
| | | | | | | | Cell-type-specific eQTL | Oligodendrocytes | 5.97E-08 | -0.69 | 0.13 | Bryois2022NN |
| | | | | | | | Cell-type-specific eQTL | Oligodendrocyte Precursor Cell | 2.87E-09 | -0.73 | 0.12 | Bryois2022NN |
| | | | | | | | Cell-type-specific eQTL | Inhibitory Neurons | 1.55E-05 | -0.48 | 0.11 | Bryois2022NN |

| | | | | | | | | | | | | |
|---|---|---|---|---|---|---|---|---|---|---|---|---|
| | | | | | | | Cell-type-specific eQTL | Excitatory Neurons | 1.20E-04 | -0.44 | 0.11 | Bryois2022NN |
| | | | | | | | Cell-type-specific eQTL | Astrocytes | 3.50E-08 | -0.65 | 0.12 | Bryois2022NN |
| | | | | | | | Cell-type-specific eQTL | CD8+ T Cell | 2.10E-04 | -0.12 | 0.03 | Aquino2023Nature |
| | | | | | | | Cell-type-specific eQTL | CD8+ T Cell | 2.10E-04 | -0.12 | 0.03 | Aquino2023Nature |
| | | | | | | | Cell-type-specific eQTL | CD4+ T Cell | 1.50E-04 | -0.10 | 0.03 | Aquino2023Nature |
| | | | | | | | Cell-type-specific eQTL | CD4+ T Cell | 1.50E-04 | -0.10 | 0.03 | Aquino2023Nature |
| | | | | | | | Cell-type-specific eQTL | Monocyte (Mono) | 7.05E-19 | -0.36 | 0.04 | Aquino2023Nature |
| | | | | | | | Cell-type-specific eQTL | Monocyte (Mono) | 7.05E-19 | -0.36 | 0.04 | Aquino2023Nature |
| | | | | | | | Cell-type-specific eQTL | Monocyte (Mono) | 4.75E-07 | -0.52 | 0.10 | Aquino2023Nature |
| | | | | | | | Cell-type-specific eQTL | Monocyte (Mono) | 4.75E-07 | -0.52 | 0.10 | Aquino2023Nature |
| | | | | | | | Cell-type-specific eQTL | Monocyte (Mono) | 6.03E-17 | -0.43 | 0.05 | Aquino2023Nature |
| | | | | | | | Cell-type-specific eQTL | Monocyte (Mono) | 6.03E-17 | -0.43 | 0.05 | Aquino2023Nature |
| | | | | | | | Cell-type-specific eQTL | CD4+ Naive T | 2.39E-04 | -0.14 | 0.04 | Aquino2023Nature |
| | | | | | | | Cell-type-specific eQTL | CD4+ Naive T | 2.39E-04 | -0.14 | 0.04 | Aquino2023Nature |

| | | | | | | | | | | | | |
|---|---|---|---|---|---|---|---|---|---|---|---|---|
| | | | | | | | Cell-type-specific eQTL | CD16+ Monocyte | 7.04E-06 | -0.60 | 0.13 | Aquino2023Nature |
| | | | | | | | Cell-type-specific eQTL | CD16+ Monocyte | 7.04E-06 | -0.60 | 0.13 | Aquino2023Nature |
| | | | | | | | Cell-type-specific eQTL | CD16+ Monocyte | 8.19E-05 | -0.53 | 0.13 | Aquino2023Nature |
| | | | | | | | Cell-type-specific eQTL | CD16+ Monocyte | 8.19E-05 | -0.53 | 0.13 | Aquino2023Nature |
| | | | | | | | Cell-type-specific eQTL | CD14+ Monocyte | 9.38E-18 | -0.38 | 0.04 | Aquino2023Nature |
| | | | | | | | Cell-type-specific eQTL | CD14+ Monocyte | 9.38E-18 | -0.38 | 0.04 | Aquino2023Nature |
| | | | | | | | Cell-type-specific eQTL | CD14+ Monocyte | 7.85E-07 | -0.61 | 0.12 | Aquino2023Nature |
| | | | | | | | Cell-type-specific eQTL | CD14+ Monocyte | 7.85E-07 | -0.61 | 0.12 | Aquino2023Nature |
| | | | | | | | Cell-type-specific eQTL | CD14+ Monocyte (CD14 Mono) | 4.21E-16 | -0.43 | 0.05 | Aquino2023Nature |
| | | | | | | | Cell-type-specific eQTL | CD14+ Monocyte (CD14 Mono) | 4.21E-16 | -0.43 | 0.05 | Aquino2023Nature |
| | | | | | | *PDHX* | Cell-type-specific eQTL | CD4+ Naive T | 3.23E-04 | -0.41 | 0.11 | Soskic2022NG |
| | | | | | | | Cell-type-specific eQTL | Oligodendrocytes | 1.02E-05 | 0.32 | 0.07 | Bryois2022NN |
| rs28550680 | 22q13.31 | 22 | 43950046 | T | C | *SAMM50* | Cell-type-specific eQTL | Naive T Cell | 8.72E-05 | 0.45 | 0.11 | Soskic2022NG |
| | | | | | | | Cell-type-specific eQTL | Naive T Cell | 6.26E-04 | 0.35 | 0.10 | Soskic2022NG |

| | | | | | | | | | | | | |
|---|---|---|---|---|---|---|---|---|---|---|---|---|
| | | | | | | | Cell-type-specific eQTL | CD4+ Effector Memory T | 4.45E-04 | 0.43 | 0.12 | Soskic2022NG |
| | | | | | | | Cell-type-specific eQTL | CD4+ Central Memory T | 4.16E-04 | 0.25 | 0.07 | Soskic2022NG |
| | | | | | | | Cell-type-specific eQTL | CD4+ Naive T | 2.02E-07 | 0.50 | 0.09 | Soskic2022NG |
| | | | | | | | Cell-type-specific eQTL | CD4 Memory | 4.00E-06 | 0.31 | 0.06 | Soskic2022NG |
| | | | | | | | Cell-type-specific eQTL | Dopaminergic Neurons | 1.04E-04 | 0.17 | 0.04 | Jerber2021NG |
| | | | | | | | Cell-type-specific eQTL | Proliferating Floor Plate Progenitors | 1.47E-04 | 0.12 | 0.03 | Jerber2021NG |

**Table S5.** mQTLs for novel loci identified in the Chinese population.

| SNP | Cytogenetic band | Chr | BP | A1 | A2 | Probe | Probe_Chr | Probe_bp | Gene | b | SE | p | Study |
|---|---|---|---|---|---|---|---|---|---|---|---|---|---|
| rs3219489 | 1p34.1 | 1 | 45331833 | G | C | cg27633763 | 1 | 45834025 | | 0.66 | 0.03 | 1.62E-86 | McRae2018SciRep;Wu2018NC |
| | | | | | | cg15605315 | | 45834250 | | -0.23 | 0.04 | 3.60E-10 | McRae2018SciRep;Wu2018NC |
| | | | | | | cg11803389 | | 45842217 | | -0.30 | 0.04 | 2.19E-15 | McRae2018SciRep;Wu2018NC |
| | | | | | | cg03123370 | | 45842540 | | -0.32 | 0.04 | 9.41E-18 | McRae2018SciRep;Wu2018NC |
| | | | | | | cg00008971 | | 45844122 | | -0.36 | 0.04 | 1.12E-23 | McRae2018SciRep;Wu2018NC |
| | | | | | | cg08483560 | | 45858623 | | 0.24 | 0.04 | 5.47E-11 | McRae2018SciRep;Wu2018NC |
| | | | | | | cg06070002 | | 45863319 | | 0.21 | 0.04 | 2.35E-08 | McRae2018SciRep;Wu2018NC |
| | | | | | | cg24332710 | | 45893374 | | 0.21 | 0.04 | 1.84E-08 | McRae2018SciRep;Wu2018NC |
| | | | | | | cg27165784 | | 45963306 | | 0.21 | 0.04 | 2.30E-08 | McRae2018SciRep;Wu2018NC |
| | | | | | | cg04296837 | | 45965037 | | -0.26 | 0.04 | 1.77E-12 | McRae2018SciRep;Wu2018NC |
| | | | | | | cg03335125 | | 45965785 | | 0.23 | 0.04 | 7.06E-10 | McRae2018SciRep;Wu2018NC |
| | | | | | | cg14220678 | | 45966268 | | 0.25 | 0.04 | 1.49E-11 | McRae2018SciRep;Wu2018NC |
| | | | | | | cg06784218 | | 45967048 | | 0.47 | 0.04 | 1.08E-38 | McRae2018SciRep;Wu2018NC |
| | | | | | | cg06170941 | | 45123809 | *BEST4* | -0.21 | 0.03 | 1.28E-11 | Hatton2024NC_East Asian ancestry |
| | | | | | | cg19804249 | | 45124946 | | -0.18 | 0.03 | 1.98E-08 | Hatton2024NC_East Asian ancestry |
| | | | | | | cg17093466 | | 45148063 | *BTBD19* | -0.22 | 0.03 | 7.03E-12 | Hatton2024NC_East Asian ancestry |
| | | | | | | cg13431108 | | 45340689 | *HECTD3* | 0.19 | 0.03 | 3.88E-09 | Hatton2024NC_East Asian ancestry |
| | | | | | | cg27633763 | | 45834025 | *TESK2* | 0.53 | 0.03 | 2.78E-70 | Hatton2024NC_East Asian ancestry |
| | | | | | | cg05343316 | | 45834040 | | 0.42 | 0.03 | 6.56E-42 | Hatton2024NC_East Asian ancestry |
| | | | | | | cg24296786 | | 45834211 | | -0.36 | 0.03 | 1.97E-32 | Hatton2024NC_East Asian ancestry |
| | | | | | | cg15605315 | | 45834250 | | -0.59 | 0.03 | 7.32E-89 | Hatton2024NC_East Asian ancestry |
| | | | | | | cg03123370 | | 45842540 | *CCDC163* | -0.26 | 0.03 | 1.94E-16 | Hatton2024NC_East Asian ancestry |
| | | | | | | cg06530443 | | 45852993 | *MMACHC* | -0.20 | 0.03 | 1.40E-10 | Hatton2024NC_East Asian ancestry |
| | | | | | | cg08483560 | | 45858623 | *PRDX1* | 0.21 | 0.03 | 2.50E-11 | Hatton2024NC_East Asian ancestry |
| | | | | | | cg06070002 | | 45863319 | | 0.22 | 0.03 | 3.21E-12 | Hatton2024NC_East Asian ancestry |

| | | | | | | | | | | | | | |
|---|---|---|---|---|---|---|---|---|---|---|---|---|---|
| | | | | | | cg24332710 | | 45893374 | *AKR1A1* | 0.34 | 0.03 | 2.74E-28 | Hatton2024NC_East Asian ancestry |
| | | | | | | cg04486013 | | 45912476 | *NASP* | 0.27 | 0.03 | 1.31E-17 | Hatton2024NC_East Asian ancestry |
| | | | | | | cg27165784 | | 45963306 | *CCDC17* | 0.31 | 0.03 | 2.14E-23 | Hatton2024NC_East Asian ancestry |
| | | | | | | cg04296837 | | 45965037 | | -0.46 | 0.03 | 1.97E-50 | Hatton2024NC_East Asian ancestry |
| | | | | | | cg03335125 | | 45965785 | | 0.21 | 0.03 | 3.34E-11 | Hatton2024NC_East Asian ancestry |
| | | | | | | cg06784218 | | 45967048 | | 0.69 | 0.03 | 1.22E-130 | Hatton2024NC_East Asian ancestry |
| | | | | | | cg16907488 | | 45967413 | | -0.26 | 0.03 | 1.02E-16 | Hatton2024NC_East Asian ancestry |
| | | | | | | cg02853497 | | 45990241 | *RPS15AP10* | 0.20 | 0.03 | 2.13E-10 | Hatton2024NC_East Asian ancestry |
| | | | | | | cg03146154 | | 46093957 | *IPP* | 0.54 | 0.03 | 1.32E-73 | Hatton2024NC_East Asian ancestry |
| | | | | | | cg22337626 | | 46297433 | *MAST2* | 0.64 | 0.03 | 6.38E-107 | Hatton2024NC_East Asian ancestry |
| | | | | | | cg08644498 | | 46379823 | *PIK3R3* | 0.18 | 0.03 | 1.43E-08 | Hatton2024NC_East Asian ancestry |
| | | | | | | cg19321695 | | 46523397 | *PIK3R3* | -0.19 | 0.03 | 3.89E-09 | Hatton2024NC_East Asian ancestry |
| | | | | | | cg04241075 | | 46684654 | *NSUN4* | -0.19 | 0.03 | 7.24E-10 | Hatton2024NC_East Asian ancestry |
| rs2165738 | 2p23.3 | 2 | 24469940 | C | G | cg11380327 | 2 | 24749997 | | 0.28 | 0.04 | 1.84E-14 | McRae2018SciRep;Wu2018NC |
| | | | | | | cg27107076 | | 25085279 | | -0.38 | 0.04 | 6.18E-27 | McRae2018SciRep;Wu2018NC |
| | | | | | | cg11023668 | | 25129486 | | -0.65 | 0.03 | 2.53E-82 | McRae2018SciRep;Wu2018NC |
| | | | | | | cg26038461 | | 25142810 | | -0.25 | 0.04 | 2.38E-12 | McRae2018SciRep;Wu2018NC |
| | | | | | | cg09505516 | | 25144744 | | -0.29 | 0.04 | 6.63E-16 | McRae2018SciRep;Wu2018NC |
| | | | | | | cg04586622 | | 25170061 | | 0.46 | 0.04 | 2.93E-38 | McRae2018SciRep;Wu2018NC |
| | | | | | | cg20518994 | | 25175985 | | 0.20 | 0.04 | 1.49E-08 | McRae2018SciRep;Wu2018NC |
| | | | | | | cg16888658 | | 25176570 | | -0.26 | 0.04 | 7.53E-13 | McRae2018SciRep;Wu2018NC |
| | | | | | | cg15423357 | | 25184430 | | 0.35 | 0.04 | 1.68E-22 | McRae2018SciRep;Wu2018NC |
| | | | | | | cg01884057 | | 25184504 | | 0.45 | 0.04 | 9.96E-37 | McRae2018SciRep;Wu2018NC |
| | | | | | | cg03879180 | | 24748067 | *NCOA1* | -0.36 | 0.05 | 3.81E-11 | Hatton2024NC_East Asian ancestry |
| | | | | | | cg11380327 | | 24749997 | | 0.60 | 0.05 | 1.31E-29 | Hatton2024NC_East Asian ancestry |
| | | | | | | cg04586622 | | 25170061 | *ADCY3* | 0.32 | 0.06 | 3.98E-09 | Hatton2024NC_East Asian ancestry |
| rs3733631 | 4q24 | 4 | 103719946 | C | G | cg24090629 | 4 | 107952049 | | -0.60 | 0.05 | 2.77E-37 | McRae2018SciRep;Wu2018NC |

| | | | | | | | | | | | | | |
|---|---|---|---|---|---|---|---|---|---|---|---|---|---|
| | | | | | | cg12866229 | | 107951171 | | 0.22 | 0.03 | 1.50E-11 | Hatton2024NC_European ancestry |
| | | | | | | cg04263186 | | 107951463 | | 0.23 | 0.03 | 4.08E-12 | Hatton2024NC_European ancestry |
| | | | | | | cg18538958 | | 107951636 | | 0.22 | 0.03 | 4.88E-11 | Hatton2024NC_European ancestry |
| | | | | | | cg14664621 | | 107951746 | | 0.22 | 0.03 | 4.87E-11 | Hatton2024NC_European ancestry |
| | | | | | | cg16461251 | | 107951807 | | 0.21 | 0.03 | 1.71E-10 | Hatton2024NC_European ancestry |
| | | | | | | cg04535008 | | 107952007 | | 0.27 | 0.03 | 9.90E-17 | Hatton2024NC_European ancestry |
| | | | | | | cg24090629 | | 107952049 | | -0.78 | 0.03 | 1.24E-131 | Hatton2024NC_European ancestry |
| | | | | | | cg00875511 | | 107952123 | | 0.28 | 0.03 | 5.29E-17 | Hatton2024NC_European ancestry |
| | | | | | | cg17160751 | | 107952227 | | 0.22 | 0.03 | 3.43E-11 | Hatton2024NC_European ancestry |
| | | | | | | cg12866229 | | 107951171 | | 0.32 | 0.03 | 4.20E-23 | Hatton2024NC_East Asian ancestry |
| | | | | | | cg04263186 | | 107951463 | | 0.25 | 0.03 | 1.75E-14 | Hatton2024NC_East Asian ancestry |
| | | | | | | cg18538958 | | 107951636 | | 0.21 | 0.03 | 8.78E-11 | Hatton2024NC_East Asian ancestry |
| | | | | | | cg14664621 | | 107951746 | | 0.22 | 0.03 | 1.88E-12 | Hatton2024NC_East Asian ancestry |
| | | | | | | cg16461251 | | 107951807 | *TACR3* | 0.20 | 0.03 | 2.63E-10 | Hatton2024NC_East Asian ancestry |
| | | | | | | cg04535008 | | 107952007 | | 0.22 | 0.03 | 2.36E-12 | Hatton2024NC_East Asian ancestry |
| | | | | | | cg24090629 | | 107952049 | | -0.87 | 0.03 | 1.20E-223 | Hatton2024NC_East Asian ancestry |
| | | | | | | cg17160751 | | 107952227 | | 0.21 | 0.03 | 2.15E-11 | Hatton2024NC_East Asian ancestry |
| rs2962370 | 5p15.1 | 5 | 17215444 | G | C | cg21979559 | 5 | 17163514 | *BASP1* | -0.25 | 0.03 | 1.47E-15 | Hatton2024NC_East Asian ancestry |
| | | | | | | cg13986272 | | 130846997 | | -0.20 | 0.04 | 2.04E-08 | McRae2018SciRep;Wu2018NC |
| | | | | | | cg26708483 | | 131941705 | | -0.37 | 0.03 | 5.56E-27 | McRae2018SciRep;Wu2018NC |
| | | | | | | cg17861653 | | 132081301 | | -0.37 | 0.04 | 1.89E-25 | McRae2018SciRep;Wu2018NC |
| | | | | | | cg24117468 | | 132082785 | | -0.53 | 0.03 | 3.61E-55 | McRae2018SciRep;Wu2018NC |
| rs272868 | 5q31.1 | 5 | 131680751 | C | G | cg18318560 | 5 | 132082952 | | -0.29 | 0.03 | 1.03E-16 | McRae2018SciRep;Wu2018NC |
| | | | | | | cg22598563 | | 132083858 | | 0.26 | 0.03 | 1.51E-13 | McRae2018SciRep;Wu2018NC |
| | | | | | | cg16205897 | | 132083987 | | 0.40 | 0.04 | 1.35E-29 | McRae2018SciRep;Wu2018NC |
| | | | | | | cg20512303 | | 132112903 | | -0.40 | 0.04 | 3.26E-29 | McRae2018SciRep;Wu2018NC |
| | | | | | | cg27615366 | | 132112918 | | -0.22 | 0.03 | 6.80E-10 | McRae2018SciRep;Wu2018NC |

| | | | | | | | | | | | | | |
|---|---|---|---|---|---|---|---|---|---|---|---|---|---|
| | | | | | | cg12564285 | | 132113048 | | -0.73 | 0.03 | 2.17E-112 | McRae2018SciRep;Wu2018NC |
| | | | | | | cg04518342 | | 132113050 | | -0.64 | 0.03 | 1.13E-80 | McRae2018SciRep;Wu2018NC |
| | | | | | | cg11843238 | | 132113135 | | -0.47 | 0.03 | 1.82E-41 | McRae2018SciRep;Wu2018NC |
| | | | | | | cg18301423 | | 132113162 | | -0.43 | 0.03 | 1.41E-34 | McRae2018SciRep;Wu2018NC |
| | | | | | | cg22638593 | | 132113202 | | -0.36 | 0.03 | 1.91E-25 | McRae2018SciRep;Wu2018NC |
| | | | | | | cg02033258 | | 132113204 | | -0.22 | 0.03 | 3.08E-10 | McRae2018SciRep;Wu2018NC |
| | | | | | | cg09877947 | | 132113230 | | -0.68 | 0.03 | 4.68E-95 | McRae2018SciRep;Wu2018NC |
| | | | | | | cg18758796 | | 132113356 | | -0.60 | 0.03 | 3.49E-69 | McRae2018SciRep;Wu2018NC |
| | | | | | | cg07262247 | | 132113673 | | -0.45 | 0.03 | 1.63E-37 | McRae2018SciRep;Wu2018NC |
| | | | | | | cg01305625 | | 132113755 | | -0.37 | 0.04 | 2.28E-25 | McRae2018SciRep;Wu2018NC |
| | | | | | | cg21743925 | | 132124851 | | 0.27 | 0.03 | 4.38E-15 | McRae2018SciRep;Wu2018NC |
| | | | | | | cg14196790 | | 132225012 | | -0.46 | 0.03 | 2.81E-40 | McRae2018SciRep;Wu2018NC |
| | | | | | | cg06968155 | | 132225089 | | -0.27 | 0.03 | 1.59E-15 | McRae2018SciRep;Wu2018NC |
| | | | | | | cg07538946 | | 132225165 | | -0.27 | 0.04 | 2.09E-14 | McRae2018SciRep;Wu2018NC |
| | | | | | | cg21911579 | | 132225202 | | -0.50 | 0.03 | 1.73E-49 | McRae2018SciRep;Wu2018NC |
| | | | | | | cg19040266 | | 132243212 | | 0.22 | 0.04 | 4.77E-10 | McRae2018SciRep;Wu2018NC |
| | | | | | | cg00594167 | | 132292012 | | 0.22 | 0.04 | 5.72E-10 | McRae2018SciRep;Wu2018NC |
| | | | | | | cg19734164 | | 132316561 | | 0.34 | 0.04 | 2.22E-22 | McRae2018SciRep;Wu2018NC |
| | | | | | | cg21138405 | | 132347677 | | 0.25 | 0.03 | 4.18E-13 | McRae2018SciRep;Wu2018NC |
| | | | | | | cg00255919 | | 132347788 | | 0.26 | 0.03 | 3.87E-14 | McRae2018SciRep;Wu2018NC |
| | | | | | | cg02551604 | | 132351615 | | -0.20 | 0.03 | 1.09E-08 | McRae2018SciRep;Wu2018NC |
| | | | | | | cg14527110 | | 132081230 | | -0.23 | 0.03 | 7.23E-14 | Hatton2024NC_East Asian ancestry |
| | | | | | | cg17861653 | | 132081301 | *P4HA2* | -0.54 | 0.03 | 2.60E-75 | Hatton2024NC_East Asian ancestry |
| | | | | | | cg07487948 | | 132086620 | | -0.20 | 0.03 | 8.53E-11 | Hatton2024NC_East Asian ancestry |
| | | | | | | cg20512303 | | 132112903 | | -0.24 | 0.03 | 8.09E-15 | Hatton2024NC_East Asian ancestry |
| | | | | | | cg12564285 | | 132113048 | *PDLIM4* | -0.38 | 0.03 | 2.98E-37 | Hatton2024NC_East Asian ancestry |
| | | | | | | cg04518342 | | 132113050 | | -0.37 | 0.03 | 3.80E-34 | Hatton2024NC_East Asian ancestry |
| | | | | | | cg11843238 | | 132113135 | | -0.32 | 0.03 | 3.44E-26 | Hatton2024NC_East Asian ancestry |

| | | | | | | | | | | | | | |
|---|---|---|---|---|---|---|---|---|---|---|---|---|---|
| | | | | | | cg18301423 | | 132113162 | | -0.22 | 0.03 | 2.15E-12 | Hatton2024NC_East Asian ancestry |
| | | | | | | cg22638593 | | 132113202 | | -0.22 | 0.03 | 5.58E-13 | Hatton2024NC_East Asian ancestry |
| | | | | | | cg09877947 | | 132113230 | | -0.40 | 0.03 | 3.99E-41 | Hatton2024NC_East Asian ancestry |
| | | | | | | cg18758796 | | 132113356 | | -0.36 | 0.03 | 2.12E-33 | Hatton2024NC_East Asian ancestry |
| | | | | | | cg07262247 | | 132113673 | | -0.36 | 0.03 | 1.04E-31 | Hatton2024NC_East Asian ancestry |
| | | | | | | cg01305625 | | 132113755 | | -0.18 | 0.03 | 1.24E-08 | Hatton2024NC_East Asian ancestry |
| | | | | | | cg21743925 | | 132124851 | | 0.20 | 0.03 | 1.19E-10 | Hatton2024NC_East Asian ancestry |
| | | | | | | cg09851765 | | 132130985 | | -0.21 | 0.03 | 1.57E-11 | Hatton2024NC_East Asian ancestry |
| | | | | | | cg13373085 | | 132149024 | *SLC22A4* | 0.41 | 0.03 | 1.38E-43 | Hatton2024NC_East Asian ancestry |
| | | | | | | cg16087826 | | 132174540 | | -0.21 | 0.03 | 1.14E-11 | Hatton2024NC_East Asian ancestry |
| | | | | | | cg14196790 | | 132225012 | *SLC22A5* | 0.33 | 0.03 | 2.78E-27 | Hatton2024NC_East Asian ancestry |
| | | | | | | cg21911579 | | 132225202 | | -0.29 | 0.03 | 5.45E-21 | Hatton2024NC_East Asian ancestry |
| | | | | | | cg24060327 | | 132225217 | | 0.83 | 0.03 | 1.55E-218 | Hatton2024NC_East Asian ancestry |
| | | | | | | cg07395648 | | 132263789 | | 0.18 | 0.03 | 2.64E-09 | Hatton2024NC_East Asian ancestry |
| | | | | | | cg00594167 | | 132292012 | *C5orf56* | 0.41 | 0.03 | 6.32E-42 | Hatton2024NC_East Asian ancestry |
| | | | | | | cg27641240 | | 132306248 | | -0.27 | 0.03 | 4.76E-19 | Hatton2024NC_East Asian ancestry |
| | | | | | | cg21171858 | | 132312687 | *IRF1* | 0.24 | 0.03 | 1.32E-14 | Hatton2024NC_East Asian ancestry |
| | | | | | | cg19734164 | | 132316561 | | 0.69 | 0.03 | 2.53E-134 | Hatton2024NC_East Asian ancestry |
| | | | | | | cg13907973 | | 132345714 | | -0.27 | 0.03 | 4.06E-18 | Hatton2024NC_East Asian ancestry |
| | | | | | | cg21138405 | | 132347677 | | -0.24 | 0.03 | 9.06E-15 | Hatton2024NC_East Asian ancestry |
| | | | | | | cg00255919 | | 132347788 | | -0.25 | 0.03 | 3.23E-16 | Hatton2024NC_East Asian ancestry |
| rs2160860 | 8p11.21 | 8 | 39823145 | T | A | cg11251498 | 8 | 40069984 | | -0.21 | 0.03 | 1.09E-09 | McRae2018SciRep;Wu2018NC |
| | | | | | | cg25981315 | | 40083916 | | 0.46 | 0.03 | 8.19E-43 | McRae2018SciRep;Wu2018NC |
| | | | | | | cg05893709 | | 40113284 | | -0.22 | 0.03 | 1.26E-10 | McRae2018SciRep;Wu2018NC |
| | | | | A | T | cg24188163 | | 40059233 | *IDO2* | 0.20 | 0.03 | 4.17E-10 | Hatton2024NC_East Asian ancestry |
| | | | | | | cg16957569 | | 40068439 | | -0.18 | 0.03 | 8.32E-09 | Hatton2024NC_East Asian ancestry |
| | | | | | | cg11251498 | | 40069984 | | 0.25 | 0.03 | 6.09E-16 | Hatton2024NC_East Asian ancestry |

| | | | | | | | | | | | | | |
|---|---|---|---|---|---|---|---|---|---|---|---|---|---|
| | | | | | | cg25981315 | | 40083916 | | -0.68 | 0.03 | 7.58E-124 | Hatton2024NC_East Asian ancestry |
| | | | | | | cg18716679 | | 40113164 | | 0.20 | 0.03 | 6.76E-10 | Hatton2024NC_East Asian ancestry |
| | | | | | | cg05621259 | | 40113227 | | 0.20 | 0.03 | 5.37E-10 | Hatton2024NC_East Asian ancestry |
| | | | | | | cg05893709 | | 40113284 | | 0.19 | 0.03 | 1.11E-09 | Hatton2024NC_East Asian ancestry |
| rs2196122 | 11p15.4 | 11 | 4885548 | C | G | cg19250571 | 11 | 4935471 | | -0.24 | 0.04 | 2.23E-08 | McRae2018SciRep;Wu2018NC |
| | | | | | | cg23295623 | | 4994112 | *OR51A7* | 0.41 | 0.06 | 1.51E-12 | Hatton2024NC_East Asian ancestry |
| rs2956095 | 11p13 | 11 | 34887382 | C | T | cg02333667 | 11 | 35072615 | | -0.51 | 0.04 | 1.42E-37 | McRae2018SciRep;Wu2018NC |
| | | | | | | cg25948180 | | 35074715 | | -0.25 | 0.04 | 5.09E-10 | McRae2018SciRep;Wu2018NC |
| | | | | | | cg24088639 | | 35075504 | | 0.32 | 0.04 | 2.26E-16 | McRae2018SciRep;Wu2018NC |
| | | | | | | cg08824847 | | 35190405 | | 0.23 | 0.04 | 2.27E-08 | McRae2018SciRep;Wu2018NC |
| | | | | | | cg16968851 | | 35259441 | | 0.23 | 0.04 | 9.43E-09 | McRae2018SciRep;Wu2018NC |
| | | | | | | cg02333667 | | 35072615 | *PDHX* | -0.67 | 0.04 | 2.73E-67 | Hatton2024NC_East Asian ancestry |
| | | | | | | cg25948180 | | 35074715 | | -0.49 | 0.04 | 8.40E-36 | Hatton2024NC_East Asian ancestry |
| | | | | | | cg24088639 | | 35075504 | | 0.23 | 0.04 | 1.77E-08 | Hatton2024NC_East Asian ancestry |
| | | | | | | cg18508148 | | 35075513 | | -0.42 | 0.04 | 1.49E-26 | Hatton2024NC_East Asian ancestry |
| | | | | | | cg11058730 | | 35075718 | | -0.57 | 0.04 | 8.73E-48 | Hatton2024NC_East Asian ancestry |
| | | | | | | cg06937548 | | 35076083 | *APIP* | -0.61 | 0.04 | 3.84E-55 | Hatton2024NC_East Asian ancestry |
| | | | | | | cg15745106 | | 35076447 | | 0.31 | 0.04 | 3.11E-14 | Hatton2024NC_East Asian ancestry |
| rs28550680 | 22q13.31 | 22 | 43950046 | T | C | cg10799608 | 22 | 44826208 | | -0.57 | 0.04 | 8.49E-51 | McRae2018SciRep;Wu2018NC |
| | | | | | | cg10799608 | | 44826208 | | -0.66 | 0.03 | 7.79E-131 | Hatton2024NC_European ancestry |
| | | | | | | cg10799608 | | 44826208 | *SAMM50* | -0.77 | 0.03 | 4.97E-148 | Hatton2024NC_East Asian ancestry |

**Table S6.** mQTLs for novel loci identified in the Chinese population using Chinese-derived methylation data.

| SNP | Cytogenetic band | Chr | BP | Probe | Probe_bp | BETA | SE | P | R2 | Study |
|---|---|---|---|---|---|---|---|---|---|---|
| rs3219489 | 1p34.1 | 1 | 45331833 | cg08664529 | 45148685 | 0.06 | 0.01 | 8.89E-15 | 0.02 | PengQ2024NG |
| | | | | cg24643057 | 45152721 | -0.16 | 0.01 | 1.94E-39 | 0.05 | PengQ2024NG |
| | | | | cg13431108 | 45340689 | 0.07 | 0.01 | 4.18E-15 | 0.02 | PengQ2024NG |
| | | | | cg00940659 | 45344257 | 0.06 | 0.01 | 6.64E-26 | 0.03 | PengQ2024NG |
| | | | | cg01182332 | 45349263 | -0.07 | 0.01 | 1.97E-14 | 0.02 | PengQ2024NG |
| | | | | cg10742135 | 45408355 | 0.15 | 0.02 | 1.89E-16 | 0.02 | PengQ2024NG |
| | | | | cg02387368 | 45644527 | 0.03 | 0.00 | 2.15E-15 | 0.02 | PengQ2024NG |
| | | | | cg19235065 | 45672790 | -0.12 | 0.01 | 5.47E-16 | 0.02 | PengQ2024NG |
| | | | | cg10384264 | 45681928 | -0.10 | 0.01 | 6.24E-48 | 0.06 | PengQ2024NG |
| | | | | cg15436426 | 45718460 | 0.08 | 0.01 | 5.75E-31 | 0.04 | PengQ2024NG |
| | | | | cg17953869 | 45737387 | -0.22 | 0.01 | 1.17E-174 | 0.20 | PengQ2024NG |
| | | | | cg08574774 | 45772522 | -0.16 | 0.01 | 3.22E-52 | 0.06 | PengQ2024NG |
| | | | | cg10186336 | 45787987 | -0.79 | 0.02 | 0.00E+00 | 0.43 | PengQ2024NG |
| | | | | cg02879171 | 45832396 | 0.05 | 0.00 | 6.02E-29 | 0.03 | PengQ2024NG |
| | | | | cg15605315 | 45834250 | -0.10 | 0.01 | 1.13E-71 | 0.09 | PengQ2024NG |
| | | | | cg00008971 | 45844122 | -0.15 | 0.01 | 5.12E-81 | 0.10 | PengQ2024NG |
| | | | | cg06530443 | 45852993 | -0.05 | 0.01 | 4.10E-15 | 0.02 | PengQ2024NG |
| | | | | cg01836612 | 45853677 | -0.25 | 0.01 | 6.78E-147 | 0.17 | PengQ2024NG |
| | | | | cg08483560 | 45858623 | 0.11 | 0.01 | 2.34E-34 | 0.04 | PengQ2024NG |
| | | | | cg06070002 | 45863319 | 0.12 | 0.01 | 1.89E-38 | 0.05 | PengQ2024NG |
| | | | | cg24642151 | 45885731 | -0.04 | 0.00 | 5.43E-21 | 0.03 | PengQ2024NG |
| | | | | cg24332710 | 45893374 | 0.09 | 0.00 | 2.14E-63 | 0.08 | PengQ2024NG |
| | | | | cg23317297 | 45893390 | 0.04 | 0.00 | 4.56E-15 | 0.02 | PengQ2024NG |

| | | | | | | | | | | |
|---|---|---|---|---|---|---|---|---|---|---|
| | | | | cg09791107 | 45900461 | -0.10 | 0.01 | 8.21E-47 | 0.06 | PengQ2024NG |
| | | | | cg22837372 | 45909689 | 0.03 | 0.00 | 4.87E-16 | 0.02 | PengQ2024NG |
| | | | | cg04486013 | 45912476 | 0.05 | 0.00 | 2.23E-33 | 0.04 | PengQ2024NG |
| | | | | cg22963343 | 45916406 | -0.28 | 0.01 | 3.68E-262 | 0.29 | PengQ2024NG |
| | | | | cg27165784 | 45963306 | 0.17 | 0.01 | 1.96E-79 | 0.10 | PengQ2024NG |
| | | | | cg04296837 | 45965037 | -0.11 | 0.01 | 2.20E-88 | 0.11 | PengQ2024NG |
| | | | | cg11750580 | 45965580 | 0.06 | 0.01 | 4.53E-11 | 0.01 | PengQ2024NG |
| | | | | cg06687305 | 45965730 | 0.07 | 0.01 | 1.28E-19 | 0.02 | PengQ2024NG |
| | | | | cg03335125 | 45965785 | 0.28 | 0.02 | 3.04E-38 | 0.05 | PengQ2024NG |
| | | | | cg11240129 | 45965861 | 0.06 | 0.01 | 1.36E-17 | 0.02 | PengQ2024NG |
| | | | | cg14220678 | 45966268 | 0.08 | 0.01 | 1.14E-10 | 0.01 | PengQ2024NG |
| | | | | cg02987388 | 45966289 | 0.08 | 0.01 | 1.03E-10 | 0.01 | PengQ2024NG |
| | | | | cg15705203 | 45966937 | 0.06 | 0.01 | 2.88E-22 | 0.03 | PengQ2024NG |
| | | | | cg06784218 | 45967048 | 0.26 | 0.01 | 2.76E-235 | 0.26 | PengQ2024NG |
| | | | | cg17721068 | 45967094 | 0.19 | 0.01 | 4.59E-222 | 0.25 | PengQ2024NG |
| | | | | cg18025133 | 45988770 | -0.05 | 0.01 | 1.22E-10 | 0.01 | PengQ2024NG |
| | | | | cg02853497 | 45990241 | 0.15 | 0.01 | 4.06E-32 | 0.04 | PengQ2024NG |
| | | | | cg09827048 | 45990308 | 0.09 | 0.01 | 9.25E-31 | 0.04 | PengQ2024NG |
| | | | | cg18210414 | 45990376 | 0.22 | 0.01 | 9.11E-108 | 0.13 | PengQ2024NG |
| | | | | cg24447116 | 46003469 | 0.06 | 0.01 | 9.50E-17 | 0.02 | PengQ2024NG |
| | | | | cg13917023 | 46012226 | 0.07 | 0.01 | 1.07E-12 | 0.01 | PengQ2024NG |
| | | | | cg27171835 | 46021998 | 0.09 | 0.01 | 3.15E-35 | 0.04 | PengQ2024NG |
| | | | | cg26899948 | 46030419 | 0.01 | 0.00 | 1.26E-04 | 0.00 | PengQ2024NG |
| | | | | cg14122995 | 46066100 | -0.11 | 0.01 | 7.01E-16 | 0.02 | PengQ2024NG |
| | | | | cg04038943 | 46073518 | 0.08 | 0.01 | 5.53E-19 | 0.02 | PengQ2024NG |

| | | | | | | | | | | |
|---|---|---|---|---|---|---|---|---|---|---|
| | | | | cg02261135 | 46096430 | -0.20 | 0.01 | 1.96E-69 | 0.08 | PengQ2024NG |
| | | | | cg04964007 | 46110328 | 0.07 | 0.01 | 2.10E-12 | 0.01 | PengQ2024NG |
| | | | | cg24200825 | 46112309 | 0.05 | 0.01 | 1.13E-12 | 0.01 | PengQ2024NG |
| | | | | cg10685628 | 46128735 | 0.05 | 0.01 | 1.26E-14 | 0.02 | PengQ2024NG |
| | | | | cg27447795 | 46186310 | 0.07 | 0.01 | 1.13E-14 | 0.02 | PengQ2024NG |
| | | | | cg06158320 | 46256392 | 0.04 | 0.00 | 8.31E-21 | 0.02 | PengQ2024NG |
| | | | | cg05450708 | 46266437 | 0.32 | 0.01 | 9.20E-111 | 0.13 | PengQ2024NG |
| | | | | cg16575491 | 46272239 | -0.06 | 0.01 | 1.37E-13 | 0.02 | PengQ2024NG |
| | | | | cg04471037 | 46272368 | -0.33 | 0.01 | 3.89E-112 | 0.14 | PengQ2024NG |
| | | | | cg15449408 | 46320744 | 0.11 | 0.01 | 1.22E-25 | 0.03 | PengQ2024NG |
| | | | | cg08644498 | 46379823 | 0.04 | 0.00 | 5.07E-32 | 0.04 | PengQ2024NG |
| | | | | cg24329497 | 46396793 | -0.04 | 0.01 | 1.62E-12 | 0.01 | PengQ2024NG |
| | | | | cg20650505 | 46405920 | 0.09 | 0.01 | 2.99E-49 | 0.06 | PengQ2024NG |
| | | | | cg13261392 | 46440505 | -0.06 | 0.01 | 6.28E-15 | 0.02 | PengQ2024NG |
| | | | | cg04181682 | 46451303 | -0.09 | 0.01 | 8.35E-58 | 0.07 | PengQ2024NG |
| | | | | cg08638929 | 46517482 | 0.13 | 0.01 | 2.32E-31 | 0.04 | PengQ2024NG |
| | | | | cg04975920 | 46523463 | -0.03 | 0.00 | 5.00E-11 | 0.01 | PengQ2024NG |
| | | | | cg22112435 | 46541689 | 0.07 | 0.01 | 1.02E-12 | 0.01 | PengQ2024NG |
| | | | | cg04036898 | 46541838 | 0.03 | 0.00 | 2.57E-12 | 0.01 | PengQ2024NG |
| | | | | cg27048324 | 46566719 | 0.05 | 0.01 | 3.35E-12 | 0.01 | PengQ2024NG |
| rs2165738 | 2p23.3 | 2 | 24469940 | cg03879180 | 24748067 | -0.08 | 0.01 | 4.71E-27 | 0.03 | PengQ2024NG |
| | | | | cg23006163 | 24749878 | 0.07 | 0.01 | 2.96E-11 | 0.01 | PengQ2024NG |
| | | | | cg11380327 | 24749997 | 0.13 | 0.01 | 7.51E-118 | 0.14 | PengQ2024NG |
| | | | | cg14300132 | 24775143 | -0.08 | 0.01 | 8.86E-31 | 0.04 | PengQ2024NG |
| | | | | cg09625143 | 24844968 | 0.08 | 0.01 | 1.16E-20 | 0.02 | PengQ2024NG |
| | | | | cg20055284 | 25007022 | -0.17 | 0.01 | 5.50E-74 | 0.09 | PengQ2024NG |

| | | | | | | | | | | |
|---|---|---|---|---|---|---|---|---|---|---|
| | | | | cg10398589 | 25013233 | -0.05 | 0.01 | 2.70E-10 | 0.01 | PengQ2024NG |
| | | | | cg26165826 | 25020517 | -0.02 | 0.01 | 9.91E-03 | 0.00 | PengQ2024NG |
| | | | | cg00260798 | 25088716 | 0.05 | 0.01 | 1.39E-13 | 0.02 | PengQ2024NG |
| | | | | cg10662790 | 25093411 | -0.08 | 0.01 | 9.10E-15 | 0.02 | PengQ2024NG |
| | | | | cg16411340 | 25098957 | -0.05 | 0.01 | 2.44E-16 | 0.02 | PengQ2024NG |
| | | | | cg11023668 | 25129486 | -0.13 | 0.01 | 1.67E-18 | 0.02 | PengQ2024NG |
| | | | | cg23528203 | 25130022 | -0.06 | 0.01 | 1.03E-12 | 0.01 | PengQ2024NG |
| | | | | cg15020926 | 25131537 | -0.11 | 0.01 | 1.09E-20 | 0.02 | PengQ2024NG |
| | | | | cg15405964 | 25132342 | -0.17 | 0.02 | 1.75E-20 | 0.02 | PengQ2024NG |
| | | | | cg09505516 | 25144744 | -0.13 | 0.02 | 5.99E-18 | 0.02 | PengQ2024NG |
| | | | | cg25286768 | 25152304 | -0.05 | 0.01 | 8.94E-11 | 0.01 | PengQ2024NG |
| | | | | cg04586622 | 25170061 | 0.16 | 0.01 | 1.65E-37 | 0.05 | PengQ2024NG |
| | | | | cg20518994 | 25175985 | 0.07 | 0.01 | 7.71E-18 | 0.02 | PengQ2024NG |
| | | | | cg15423357 | 25184430 | 0.15 | 0.01 | 1.65E-26 | 0.03 | PengQ2024NG |
| | | | | cg01884057 | 25184504 | 0.17 | 0.01 | 4.84E-40 | 0.05 | PengQ2024NG |
| | | | | cg03554320 | 25299659 | 0.10 | 0.01 | 7.60E-25 | 0.03 | PengQ2024NG |
| rs10024137 | 4q13.2 | 4 | 68667043 | cg20840634 | 72769309 | -0.30 | 0.02 | 1.65E-61 | 0.08 | PengQ2024NG |
| | | | | cg22264257 | 72871485 | -0.34 | 0.02 | 8.57E-78 | 0.10 | PengQ2024NG |
| | | | | cg06071003 | 72894115 | 0.19 | 0.02 | 9.78E-28 | 0.03 | PengQ2024NG |
| | | | | cg26596456 | 72933136 | 0.43 | 0.02 | 5.17E-91 | 0.11 | PengQ2024NG |
| | | | | cg04962286 | 72933653 | 0.12 | 0.01 | 3.01E-22 | 0.03 | PengQ2024NG |
| | | | | cg03363834 | 72949395 | -0.06 | 0.01 | 1.09E-11 | 0.01 | PengQ2024NG |
| | | | | cg19508985 | 73106571 | -0.74 | 0.03 | 5.36E-103 | 0.12 | PengQ2024NG |
| | | | | cg22519480 | 73148900 | 0.11 | 0.02 | 1.46E-11 | 0.01 | PengQ2024NG |
| | | | | cg21287356 | 73154649 | 0.05 | 0.01 | 2.13E-12 | 0.01 | PengQ2024NG |
| | | | | cg21844184 | 73194533 | -0.12 | 0.01 | 1.14E-29 | 0.04 | PengQ2024NG |
| | | | | cg25583503 | 73297470 | 0.05 | 0.01 | 4.35E-09 | 0.01 | PengQ2024NG |
| rs3733631 | 4q24 | 4 | 1.04E+08 | cg12866229 | 107951171 | 0.07 | 0.01 | 3.83E-22 | 0.03 | PengQ2024NG |

| | | | | | | | | | | |
|---|---|---|---|---|---|---|---|---|---|---|
| | | | | cg04263186 | 107951463 | 0.10 | 0.01 | 6.13E-20 | 0.02 | PengQ2024NG |
| | | | | cg18538958 | 107951636 | 0.10 | 0.01 | 8.51E-16 | 0.02 | PengQ2024NG |
| | | | | cg16461251 | 107951807 | 0.09 | 0.01 | 3.34E-17 | 0.02 | PengQ2024NG |
| | | | | cg04535008 | 107952007 | 0.10 | 0.01 | 4.79E-18 | 0.02 | PengQ2024NG |
| | | | | cg06180821 | 107952130 | 0.04 | 0.01 | 2.38E-12 | 0.01 | PengQ2024NG |
| | | | | cg17160751 | 107952227 | 0.10 | 0.01 | 8.95E-23 | 0.03 | PengQ2024NG |
| | | | | cg05389335 | 107952296 | 0.06 | 0.01 | 6.98E-10 | 0.01 | PengQ2024NG |
| rs2962370 | 5p15.1 | 5 | 17215444 | cg17857235 | 17153173 | -0.10 | 0.01 | 4.21E-70 | 0.09 | PengQ2024NG |
| rs272868 | 5q31.1 | 5 | 1.32E+08 | cg18276568 | 131965364 | 0.09 | 0.01 | 5.13E-23 | 0.03 | PengQ2024NG |
| | | | | cg07929728 | 131993836 | 0.05 | 0.01 | 2.74E-15 | 0.02 | PengQ2024NG |
| | | | | cg09011406 | 132034844 | 0.05 | 0.01 | 1.14E-12 | 0.01 | PengQ2024NG |
| | | | | cg05026773 | 132037479 | -0.06 | 0.01 | 3.29E-11 | 0.01 | PengQ2024NG |
| | | | | cg14527110 | 132081230 | -0.03 | 0.00 | 2.38E-16 | 0.02 | PengQ2024NG |
| | | | | cg17861653 | 132081301 | -0.29 | 0.01 | 4.65E-109 | 0.13 | PengQ2024NG |
| | | | | cg14151197 | 132081420 | -0.07 | 0.01 | 3.10E-19 | 0.02 | PengQ2024NG |
| | | | | cg07487948 | 132086620 | -0.07 | 0.01 | 8.19E-26 | 0.03 | PengQ2024NG |
| | | | | cg14336706 | 132112003 | 0.06 | 0.01 | 6.77E-21 | 0.02 | PengQ2024NG |
| | | | | cg12564285 | 132113048 | -0.16 | 0.01 | 4.12E-66 | 0.08 | PengQ2024NG |
| | | | | cg04518342 | 132113050 | -0.19 | 0.01 | 3.46E-69 | 0.08 | PengQ2024NG |
| | | | | cg18301423 | 132113162 | -0.18 | 0.01 | 3.11E-56 | 0.07 | PengQ2024NG |
| | | | | cg22638593 | 132113202 | -0.15 | 0.01 | 5.96E-41 | 0.05 | PengQ2024NG |
| | | | | cg02033258 | 132113204 | -0.13 | 0.01 | 6.98E-24 | 0.03 | PengQ2024NG |
| | | | | cg07262247 | 132113673 | -0.14 | 0.01 | 3.85E-36 | 0.04 | PengQ2024NG |
| | | | | cg01305625 | 132113755 | -0.08 | 0.01 | 1.11E-19 | 0.02 | PengQ2024NG |
| | | | | cg11233650 | 132139333 | 0.11 | 0.01 | 5.70E-31 | 0.04 | PengQ2024NG |
| | | | | cg13373085 | 132149024 | 0.16 | 0.01 | 2.90E-74 | 0.09 | PengQ2024NG |
| | | | | cg20595710 | 132149191 | 0.07 | 0.01 | 1.27E-22 | 0.03 | PengQ2024NG |
| | | | | cg03853968 | 132151116 | 0.06 | 0.01 | 7.72E-21 | 0.02 | PengQ2024NG |

| | | | | | | | | | | |
|---|---|---|---|---|---|---|---|---|---|---|
| | | | | cg26482198 | 132156797 | -0.03 | 0.00 | 6.80E-23 | 0.03 | PengQ2024NG |
| | | | | cg05883541 | 132157366 | -0.20 | 0.01 | 7.56E-165 | 0.19 | PengQ2024NG |
| | | | | cg27094470 | 132157519 | -0.08 | 0.00 | 4.67E-75 | 0.09 | PengQ2024NG |
| | | | | cg16087826 | 132174540 | -0.07 | 0.01 | 4.81E-25 | 0.03 | PengQ2024NG |
| | | | | cg14622915 | 132177678 | -0.12 | 0.01 | 1.07E-36 | 0.04 | PengQ2024NG |
| | | | | cg02948635 | 132190740 | -0.05 | 0.00 | 5.13E-24 | 0.03 | PengQ2024NG |
| | | | | cg12468553 | 132199564 | -0.12 | 0.01 | 5.89E-28 | 0.03 | PengQ2024NG |
| | | | | cg00318034 | 132200982 | -0.10 | 0.01 | 1.82E-51 | 0.06 | PengQ2024NG |
| | | | | cg10979594 | 132217253 | -0.07 | 0.01 | 5.43E-11 | 0.01 | PengQ2024NG |
| | | | | cg02983324 | 132219235 | -0.22 | 0.01 | 2.40E-178 | 0.21 | PengQ2024NG |
| | | | | cg01023783 | 132221221 | -0.21 | 0.01 | 7.43E-88 | 0.11 | PengQ2024NG |
| | | | | cg14196790 | 132225012 | 0.13 | 0.01 | 3.36E-40 | 0.05 | PengQ2024NG |
| | | | | cg07538946 | 132225165 | 0.06 | 0.01 | 8.10E-14 | 0.02 | PengQ2024NG |
| | | | | cg21911579 | 132225202 | -0.30 | 0.01 | 1.51E-156 | 0.18 | PengQ2024NG |
| | | | | cg16647868 | 132226043 | -0.11 | 0.01 | 8.68E-83 | 0.10 | PengQ2024NG |
| | | | | cg04774966 | 132226308 | -0.13 | 0.01 | 1.03E-70 | 0.09 | PengQ2024NG |
| | | | | cg06796115 | 132226922 | -0.06 | 0.01 | 5.28E-19 | 0.02 | PengQ2024NG |
| | | | | cg16022017 | 132233619 | -0.04 | 0.01 | 5.19E-13 | 0.01 | PengQ2024NG |
| | | | | cg26647941 | 132234242 | -0.14 | 0.01 | 1.03E-71 | 0.09 | PengQ2024NG |
| | | | | cg03979695 | 132241125 | -0.04 | 0.01 | 1.29E-11 | 0.01 | PengQ2024NG |
| | | | | cg05640674 | 132242534 | -0.05 | 0.01 | 2.69E-17 | 0.02 | PengQ2024NG |
| | | | | cg12610030 | 132242977 | 0.05 | 0.01 | 3.73E-16 | 0.02 | PengQ2024NG |
| | | | | cg06516533 | 132252444 | 0.15 | 0.01 | 1.09E-24 | 0.03 | PengQ2024NG |
| | | | | cg03800775 | 132263769 | 0.10 | 0.01 | 6.74E-17 | 0.02 | PengQ2024NG |
| | | | | cg07395648 | 132263789 | 0.11 | 0.01 | 1.69E-13 | 0.02 | PengQ2024NG |
| | | | | cg06032302 | 132282636 | -0.06 | 0.01 | 1.39E-13 | 0.02 | PengQ2024NG |

| | | | | | | | | | | |
|---|---|---|---|---|---|---|---|---|---|---|
| | | | | cg21881287 | 132286555 | -0.05 | 0.01 | 3.23E-21 | 0.03 | PengQ2024NG |
| | | | | cg00594167 | 132292012 | 0.31 | 0.02 | 2.26E-79 | 0.10 | PengQ2024NG |
| | | | | cg15380772 | 132295458 | -0.08 | 0.01 | 9.72E-35 | 0.04 | PengQ2024NG |
| | | | | cg27641240 | 132306248 | -0.24 | 0.03 | 9.73E-22 | 0.03 | PengQ2024NG |
| | | | | cg11586513 | 132307162 | 0.08 | 0.01 | 5.07E-27 | 0.03 | PengQ2024NG |
| | | | | cg10248037 | 132312682 | 0.06 | 0.01 | 2.02E-15 | 0.02 | PengQ2024NG |
| | | | | cg21171858 | 132312687 | 0.07 | 0.01 | 5.41E-26 | 0.03 | PengQ2024NG |
| | | | | cg23103826 | 132315794 | 0.08 | 0.00 | 4.64E-64 | 0.08 | PengQ2024NG |
| | | | | cg19734164 | 132316561 | 0.65 | 0.02 | 1.25E-214 | 0.24 | PengQ2024NG |
| | | | | cg05635287 | 132325645 | -0.15 | 0.01 | 2.04E-53 | 0.07 | PengQ2024NG |
| | | | | cg09248869 | 132328662 | -0.16 | 0.01 | 1.65E-32 | 0.04 | PengQ2024NG |
| | | | | cg02964085 | 132328834 | -0.10 | 0.01 | 8.88E-45 | 0.05 | PengQ2024NG |
| | | | | cg13907973 | 132345714 | -0.31 | 0.03 | 6.75E-32 | 0.04 | PengQ2024NG |
| | | | | cg21138405 | 132347677 | -0.17 | 0.02 | 1.59E-17 | 0.02 | PengQ2024NG |
| | | | | cg00255919 | 132347788 | -0.18 | 0.02 | 1.26E-17 | 0.02 | PengQ2024NG |
| rs2534815 | 6p21.32 | 6 | 30499127 | cg08027810 | 29468164 | -0.11 | 0.02 | 2.13E-13 | 0.02 | PengQ2024NG |
| | | | | cg23732781 | 29469698 | 0.09 | 0.01 | 7.00E-22 | 0.03 | PengQ2024NG |
| | | | | cg12292060 | 29472703 | 0.06 | 0.01 | 1.51E-13 | 0.02 | PengQ2024NG |
| | | | | cg09740560 | 29477101 | 0.04 | 0.01 | 1.55E-11 | 0.01 | PengQ2024NG |
| | | | | cg16994534 | 29494837 | 0.04 | 0.01 | 2.25E-12 | 0.01 | PengQ2024NG |
| | | | | cg00433866 | 29498345 | 0.11 | 0.01 | 1.91E-18 | 0.02 | PengQ2024NG |
| | | | | cg17977304 | 29498691 | 0.10 | 0.01 | 3.13E-18 | 0.02 | PengQ2024NG |
| | | | | cg16118803 | 29504420 | 0.12 | 0.01 | 6.02E-17 | 0.02 | PengQ2024NG |
| | | | | cg05279622 | 29504462 | 0.13 | 0.02 | 1.10E-15 | 0.02 | PengQ2024NG |
| | | | | cg08725892 | 29504690 | 0.12 | 0.02 | 2.36E-13 | 0.02 | PengQ2024NG |
| | | | | cg05813221 | 29504692 | 0.12 | 0.01 | 1.41E-17 | 0.02 | PengQ2024NG |
| | | | | cg24938286 | 29505999 | 0.12 | 0.02 | 3.34E-11 | 0.01 | PengQ2024NG |
| | | | | cg15841167 | 29508326 | 0.15 | 0.02 | 3.06E-14 | 0.02 | PengQ2024NG |

| | | | | | | | | | | |
|---|---|---|---|---|---|---|---|---|---|---|
| | | | | cg03147503 | 29633969 | 0.09 | 0.01 | 1.42E-17 | 0.02 | PengQ2024NG |
| | | | | cg23019585 | 29509808 | 0.07 | 0.01 | 7.56E-15 | 0.02 | PengQ2024NG |
| | | | | cg20704602 | 29510071 | 0.09 | 0.01 | 5.05E-16 | 0.02 | PengQ2024NG |
| | | | | cg20198768 | 29510279 | 0.08 | 0.01 | 2.19E-14 | 0.02 | PengQ2024NG |
| | | | | cg24444631 | 29511066 | 0.07 | 0.01 | 7.40E-15 | 0.02 | PengQ2024NG |
| | | | | cg06061002 | 29513618 | 0.07 | 0.01 | 1.46E-15 | 0.02 | PengQ2024NG |
| | | | | cg12833048 | 29514283 | 0.05 | 0.01 | 2.51E-13 | 0.02 | PengQ2024NG |
| | | | | cg00539542 | 29519243 | -0.12 | 0.01 | 2.77E-19 | 0.02 | PengQ2024NG |
| | | | | cg12463578 | 29519454 | -0.14 | 0.01 | 2.53E-22 | 0.03 | PengQ2024NG |
| | | | | cg25978138 | 29522858 | -0.27 | 0.04 | 1.52E-13 | 0.02 | PengQ2024NG |
| | | | | cg11747594 | 29522922 | -0.26 | 0.04 | 2.07E-13 | 0.02 | PengQ2024NG |
| | | | | cg15708526 | 29522968 | -0.25 | 0.03 | 1.62E-15 | 0.02 | PengQ2024NG |
| | | | | cg04071440 | 29522972 | -0.25 | 0.03 | 1.43E-15 | 0.02 | PengQ2024NG |
| | | | | cg08022281 | 29523042 | -0.16 | 0.02 | 1.38E-15 | 0.02 | PengQ2024NG |
| | | | | cg10648573 | 29523045 | -0.16 | 0.02 | 8.29E-13 | 0.01 | PengQ2024NG |
| | | | | cg22494932 | 29523076 | -0.34 | 0.04 | 4.74E-15 | 0.02 | PengQ2024NG |
| | | | | cg25699073 | 29523078 | -0.30 | 0.04 | 3.56E-15 | 0.02 | PengQ2024NG |
| | | | | cg07134666 | 29523097 | -0.30 | 0.05 | 1.65E-11 | 0.01 | PengQ2024NG |
| | | | | cg16885113 | 29523204 | -0.35 | 0.05 | 2.50E-13 | 0.02 | PengQ2024NG |
| | | | | cg20228636 | 29523222 | -0.26 | 0.04 | 1.83E-11 | 0.01 | PengQ2024NG |
| | | | | cg11383134 | 29523287 | -0.31 | 0.04 | 7.51E-13 | 0.01 | PengQ2024NG |
| | | | | cg03449857 | 29523320 | -0.28 | 0.04 | 1.45E-12 | 0.01 | PengQ2024NG |
| | | | | cg19636627 | 29523782 | -0.35 | 0.04 | 6.90E-16 | 0.02 | PengQ2024NG |
| | | | | cg10876145 | 29545327 | 0.34 | 0.03 | 4.39E-24 | 0.03 | PengQ2024NG |
| | | | | cg15933418 | 29547999 | -0.07 | 0.01 | 2.36E-10 | 0.01 | PengQ2024NG |
| | | | | cg15018934 | 29564470 | -0.21 | 0.02 | 7.52E-26 | 0.03 | PengQ2024NG |
| | | | | cg11201654 | 29565503 | 0.17 | 0.02 | 1.85E-18 | 0.02 | PengQ2024NG |
| | | | | cg05358170 | 29567135 | 0.06 | 0.01 | 1.35E-11 | 0.01 | PengQ2024NG |
| | | | | cg25252977 | 29571384 | 0.18 | 0.03 | 7.42E-11 | 0.01 | PengQ2024NG |

| | | | | | | | | | | |
|---|---|---|---|---|---|---|---|---|---|---|
| | | | | cg17264941 | 29573143 | -0.13 | 0.01 | 2.70E-20 | 0.02 | PengQ2024NG |
| | | | | cg16101636 | 29586173 | -0.06 | 0.01 | 4.47E-11 | 0.01 | PengQ2024NG |
| | | | | cg24778368 | 29600168 | -0.04 | 0.00 | 4.00E-16 | 0.02 | PengQ2024NG |
| | | | | cg16051154 | 29630549 | 0.15 | 0.02 | 5.04E-13 | 0.01 | PengQ2024NG |
| | | | | cg24621922 | 29631823 | 0.09 | 0.01 | 7.54E-18 | 0.02 | PengQ2024NG |
| | | | | cg08560874 | 29631966 | -0.08 | 0.01 | 2.98E-15 | 0.02 | PengQ2024NG |
| | | | | cg16787652 | 29631974 | -0.07 | 0.01 | 1.24E-14 | 0.02 | PengQ2024NG |
| | | | | cg17670496 | 29631991 | -0.08 | 0.01 | 1.88E-13 | 0.02 | PengQ2024NG |
| | | | | cg17512365 | 29632109 | -0.06 | 0.01 | 5.72E-15 | 0.02 | PengQ2024NG |
| | | | | cg24699876 | 29635962 | -0.09 | 0.01 | 4.15E-17 | 0.02 | PengQ2024NG |
| | | | | cg17213462 | 29667409 | 0.05 | 0.00 | 7.79E-30 | 0.04 | PengQ2024NG |
| | | | | cg10556772 | 29667432 | 0.13 | 0.01 | 7.87E-26 | 0.03 | PengQ2024NG |
| | | | | cg03096746 | 29667434 | 0.09 | 0.01 | 6.47E-21 | 0.02 | PengQ2024NG |
| | | | | cg03432955 | 29667499 | 0.12 | 0.01 | 2.01E-28 | 0.03 | PengQ2024NG |
| | | | | cg09865095 | 29795557 | 0.08 | 0.01 | 8.02E-26 | 0.03 | PengQ2024NG |
| | | | | cg00126638 | 29795583 | 0.08 | 0.01 | 6.31E-26 | 0.03 | PengQ2024NG |
| | | | | cg03521696 | 29795595 | 0.13 | 0.01 | 2.87E-28 | 0.03 | PengQ2024NG |
| | | | | cg03997176 | 29667857 | -0.06 | 0.01 | 3.14E-12 | 0.01 | PengQ2024NG |
| | | | | cg19141489 | 29667859 | -0.06 | 0.01 | 1.56E-11 | 0.01 | PengQ2024NG |
| | | | | cg11862551 | 29689771 | 0.34 | 0.03 | 5.15E-33 | 0.04 | PengQ2024NG |
| | | | | cg03118604 | 29689887 | 0.17 | 0.02 | 7.52E-18 | 0.02 | PengQ2024NG |
| | | | | cg23376846 | 29689962 | 0.06 | 0.01 | 2.58E-16 | 0.02 | PengQ2024NG |
| | | | | cg10181330 | 29690051 | 0.22 | 0.02 | 2.22E-31 | 0.04 | PengQ2024NG |
| | | | | cg10821226 | 29690085 | 0.13 | 0.01 | 3.30E-23 | 0.03 | PengQ2024NG |
| | | | | cg03863342 | 29703491 | 0.19 | 0.02 | 4.33E-32 | 0.04 | PengQ2024NG |
| | | | | cg23273834 | 29726833 | -0.03 | 0.00 | 6.31E-18 | 0.02 | PengQ2024NG |
| | | | | cg03861427 | 29726906 | 0.13 | 0.01 | 5.06E-20 | 0.02 | PengQ2024NG |
| | | | | cg01362455 | 29727071 | -0.11 | 0.01 | 2.40E-20 | 0.02 | PengQ2024NG |
| | | | | cg18255335 | 29727693 | 0.09 | 0.01 | 4.70E-14 | 0.02 | PengQ2024NG |

| | | | | | | | | | | |
|---|---|---|---|---|---|---|---|---|---|---|
| | | | | cg11507793 | 29727944 | -0.08 | 0.01 | 2.81E-11 | 0.01 | PengQ2024NG |
| | | | | cg20114890 | 29728011 | -0.16 | 0.02 | 1.40E-14 | 0.02 | PengQ2024NG |
| | | | | cg06354398 | 29728058 | -0.18 | 0.02 | 1.23E-23 | 0.03 | PengQ2024NG |
| | | | | cg11406274 | 29728145 | -0.08 | 0.01 | 1.73E-10 | 0.01 | PengQ2024NG |
| | | | | cg04276750 | 29728507 | -0.30 | 0.03 | 1.47E-23 | 0.03 | PengQ2024NG |
| | | | | cg16716514 | 29758856 | -0.12 | 0.02 | 1.96E-15 | 0.02 | PengQ2024NG |
| | | | | cg16302021 | 29759835 | -0.09 | 0.01 | 2.85E-19 | 0.02 | PengQ2024NG |
| | | | | cg10595298 | 29759850 | -0.10 | 0.01 | 5.21E-44 | 0.05 | PengQ2024NG |
| | | | | cg24751894 | 29760036 | -0.65 | 0.05 | 1.56E-39 | 0.05 | PengQ2024NG |
| | | | | cg15070894 | 29760057 | -0.61 | 0.04 | 2.83E-41 | 0.05 | PengQ2024NG |
| | | | | cg04520169 | 29760090 | -0.55 | 0.04 | 1.06E-36 | 0.04 | PengQ2024NG |
| | | | | cg23237314 | 29760092 | -0.61 | 0.05 | 3.14E-35 | 0.04 | PengQ2024NG |
| | | | | cg07657192 | 29760881 | 0.11 | 0.01 | 4.77E-15 | 0.02 | PengQ2024NG |
| | | | | cg01499815 | 29760969 | -0.17 | 0.02 | 2.01E-15 | 0.02 | PengQ2024NG |
| | | | | cg15671450 | 29761011 | -0.34 | 0.03 | 6.37E-23 | 0.03 | PengQ2024NG |
| | | | | cg26121931 | 29761099 | -0.20 | 0.02 | 4.65E-21 | 0.02 | PengQ2024NG |
| | | | | cg24838316 | 29761155 | -0.23 | 0.03 | 1.59E-16 | 0.02 | PengQ2024NG |
| | | | | cg16845367 | 29767830 | -0.19 | 0.03 | 4.08E-10 | 0.01 | PengQ2024NG |
| | | | | cg07163603 | 29775939 | -0.09 | 0.01 | 7.29E-39 | 0.05 | PengQ2024NG |
| | | | | cg05523662 | 29775989 | -0.08 | 0.01 | 9.11E-14 | 0.02 | PengQ2024NG |
| | | | | cg11808100 | 29776643 | 0.38 | 0.03 | 7.17E-30 | 0.04 | PengQ2024NG |
| | | | | cg25548869 | 29776664 | -0.08 | 0.01 | 4.31E-15 | 0.02 | PengQ2024NG |
| | | | | cg09803951 | 29776684 | -0.06 | 0.01 | 7.41E-12 | 0.01 | PengQ2024NG |
| | | | | cg14018363 | 29777153 | 0.22 | 0.03 | 1.20E-12 | 0.01 | PengQ2024NG |
| | | | | cg25637655 | 29777430 | -0.29 | 0.02 | 1.71E-34 | 0.04 | PengQ2024NG |
| | | | | cg17608381 | 29777438 | -0.24 | 0.02 | 4.04E-29 | 0.04 | PengQ2024NG |
| | | | | cg11946459 | 29777446 | -0.23 | 0.02 | 1.16E-30 | 0.04 | PengQ2024NG |
| | | | | cg14357455 | 29789850 | 0.08 | 0.01 | 1.30E-12 | 0.01 | PengQ2024NG |
| | | | | cg03977339 | 29790152 | -0.09 | 0.01 | 4.83E-11 | 0.01 | PengQ2024NG |

| | | | | | | | | | | |
|---|---|---|---|---|---|---|---|---|---|---|
| | | | | cg25423002 | 29790195 | 0.10 | 0.01 | 1.41E-15 | 0.02 | PengQ2024NG |
| | | | | cg14375543 | 29933411 | -0.07 | 0.01 | 1.51E-12 | 0.01 | PengQ2024NG |
| | | | | cg04007531 | 29806432 | -0.15 | 0.02 | 1.83E-15 | 0.02 | PengQ2024NG |
| | | | | cg26071631 | 29806633 | -0.10 | 0.02 | 6.74E-10 | 0.01 | PengQ2024NG |
| | | | | cg07792871 | 29806878 | -0.27 | 0.04 | 9.97E-11 | 0.01 | PengQ2024NG |
| | | | | cg07370894 | 29807115 | 0.09 | 0.01 | 1.23E-16 | 0.02 | PengQ2024NG |
| | | | | cg19593490 | 29807136 | 0.08 | 0.01 | 3.27E-18 | 0.02 | PengQ2024NG |
| | | | | cg00599564 | 29807328 | 0.06 | 0.01 | 1.25E-17 | 0.02 | PengQ2024NG |
| | | | | cg23244913 | 29807335 | 0.10 | 0.01 | 5.44E-16 | 0.02 | PengQ2024NG |
| | | | | cg06710082 | 29807341 | 0.10 | 0.01 | 1.11E-14 | 0.02 | PengQ2024NG |
| | | | | cg04623837 | 29807352 | 0.11 | 0.01 | 5.37E-15 | 0.02 | PengQ2024NG |
| | | | | cg16368146 | 29809654 | 0.11 | 0.01 | 5.81E-27 | 0.03 | PengQ2024NG |
| | | | | cg10221391 | 29824696 | 0.10 | 0.01 | 5.99E-16 | 0.02 | PengQ2024NG |
| | | | | cg24400518 | 29836779 | 0.04 | 0.01 | 3.00E-12 | 0.01 | PengQ2024NG |
| | | | | cg22012577 | 29837116 | -0.04 | 0.01 | 2.56E-11 | 0.01 | PengQ2024NG |
| | | | | cg04628742 | 29837550 | 0.11 | 0.01 | 9.26E-14 | 0.02 | PengQ2024NG |
| | | | | cg17590657 | 29837903 | -0.13 | 0.01 | 1.70E-25 | 0.03 | PengQ2024NG |
| | | | | cg09368485 | 29837964 | -0.05 | 0.01 | 5.23E-12 | 0.01 | PengQ2024NG |
| | | | | cg23318893 | 29837975 | -0.09 | 0.01 | 3.24E-18 | 0.02 | PengQ2024NG |
| | | | | cg23694113 | 29838725 | -0.07 | 0.01 | 1.36E-22 | 0.03 | PengQ2024NG |
| | | | | cg23483840 | 29841838 | -0.07 | 0.01 | 2.34E-11 | 0.01 | PengQ2024NG |
| | | | | cg24694606 | 29845126 | -0.07 | 0.01 | 8.12E-12 | 0.01 | PengQ2024NG |
| | | | | cg08397993 | 29845150 | -0.19 | 0.02 | 5.75E-27 | 0.03 | PengQ2024NG |
| | | | | cg18066181 | 29845280 | -0.09 | 0.01 | 6.80E-16 | 0.02 | PengQ2024NG |
| | | | | cg12854611 | 29864080 | -0.06 | 0.01 | 2.47E-15 | 0.02 | PengQ2024NG |
| | | | | cg01915885 | 29867804 | 0.07 | 0.01 | 1.22E-11 | 0.01 | PengQ2024NG |
| | | | | cg02881170 | 29873070 | 0.12 | 0.02 | 4.56E-11 | 0.01 | PengQ2024NG |
| | | | | cg20380424 | 29874797 | -0.15 | 0.02 | 1.90E-17 | 0.02 | PengQ2024NG |
| | | | | cg26453130 | 29903039 | -0.04 | 0.01 | 5.99E-12 | 0.01 | PengQ2024NG |

| | | | | | | | | | | |
|---|---|---|---|---|---|---|---|---|---|---|
| | | | | cg17322683 | 29903130 | -0.03 | 0.01 | 2.55E-08 | 0.01 | PengQ2024NG |
| | | | | cg09093485 | 29903203 | -0.09 | 0.01 | 4.74E-12 | 0.01 | PengQ2024NG |
| | | | | cg24330456 | 29903223 | -0.10 | 0.01 | 5.81E-15 | 0.02 | PengQ2024NG |
| | | | | cg03293507 | 29903246 | -0.09 | 0.01 | 2.04E-16 | 0.02 | PengQ2024NG |
| | | | | cg16582419 | 29903258 | -0.09 | 0.01 | 1.22E-16 | 0.02 | PengQ2024NG |
| | | | | cg12736877 | 29903273 | -0.13 | 0.02 | 1.85E-15 | 0.02 | PengQ2024NG |
| | | | | cg01286685 | 29903380 | -0.12 | 0.02 | 6.93E-14 | 0.02 | PengQ2024NG |
| | | | | cg23712018 | 29903390 | -0.20 | 0.02 | 1.74E-16 | 0.02 | PengQ2024NG |
| | | | | cg00947782 | 29903423 | -0.19 | 0.02 | 8.78E-16 | 0.02 | PengQ2024NG |
| | | | | cg03343571 | 29903450 | -0.15 | 0.02 | 2.37E-13 | 0.02 | PengQ2024NG |
| | | | | cg13185413 | 29903454 | -0.16 | 0.02 | 3.53E-12 | 0.01 | PengQ2024NG |
| | | | | cg06249604 | 29903622 | -0.16 | 0.02 | 4.89E-12 | 0.01 | PengQ2024NG |
| | | | | cg20249327 | 29903624 | -0.15 | 0.02 | 1.71E-11 | 0.01 | PengQ2024NG |
| | | | | cg15877520 | 29903628 | -0.16 | 0.02 | 1.78E-11 | 0.01 | PengQ2024NG |
| | | | | cg13918754 | 29903651 | -0.21 | 0.03 | 4.61E-12 | 0.01 | PengQ2024NG |
| | | | | cg09279736 | 29903656 | -0.14 | 0.02 | 1.97E-12 | 0.01 | PengQ2024NG |
| | | | | cg07382347 | 29903680 | -0.20 | 0.03 | 1.86E-13 | 0.02 | PengQ2024NG |
| | | | | cg13401893 | 29903683 | -0.18 | 0.02 | 1.00E-13 | 0.02 | PengQ2024NG |
| | | | | cg12633154 | 29903714 | -0.19 | 0.03 | 2.12E-14 | 0.02 | PengQ2024NG |
| | | | | cg16078649 | 29903724 | -0.21 | 0.03 | 1.45E-14 | 0.02 | PengQ2024NG |
| | | | | cg10930308 | 29903772 | -0.26 | 0.03 | 4.74E-17 | 0.02 | PengQ2024NG |
| | | | | cg02188185 | 29903848 | -0.10 | 0.01 | 6.59E-15 | 0.02 | PengQ2024NG |
| | | | | cg26730543 | 29904049 | -0.08 | 0.01 | 3.01E-16 | 0.02 | PengQ2024NG |
| | | | | cg24103044 | 29908456 | -0.12 | 0.01 | 2.69E-15 | 0.02 | PengQ2024NG |
| | | | | cg03782043 | 29908509 | 0.05 | 0.01 | 2.13E-17 | 0.02 | PengQ2024NG |
| | | | | cg01692379 | 29935701 | 0.05 | 0.01 | 9.93E-18 | 0.02 | PengQ2024NG |
| | | | | cg13196879 | 29935875 | -0.12 | 0.01 | 6.50E-20 | 0.02 | PengQ2024NG |
| | | | | cg13436843 | 29943329 | -0.10 | 0.01 | 7.11E-12 | 0.01 | PengQ2024NG |
| | | | | cg13446859 | 29958587 | -0.08 | 0.01 | 3.82E-13 | 0.01 | PengQ2024NG |

| | | | | | | | | | | |
|---|---|---|---|---|---|---|---|---|---|---|
| | | | | cg01641092 | 29967973 | -0.11 | 0.02 | 4.80E-09 | 0.01 | PengQ2024NG |
| | | | | cg13044052 | 29969159 | 0.06 | 0.01 | 9.60E-11 | 0.01 | PengQ2024NG |
| | | | | cg06195293 | 29978050 | -0.07 | 0.01 | 9.02E-16 | 0.02 | PengQ2024NG |
| | | | | cg08996678 | 29978092 | 0.05 | 0.01 | 1.14E-12 | 0.01 | PengQ2024NG |
| | | | | cg04006457 | 29981440 | 0.04 | 0.01 | 3.85E-13 | 0.01 | PengQ2024NG |
| | | | | cg08464235 | 29984302 | 0.04 | 0.01 | 2.11E-11 | 0.01 | PengQ2024NG |
| | | | | cg19495308 | 29985186 | 0.09 | 0.01 | 1.42E-19 | 0.02 | PengQ2024NG |
| | | | | cg08060195 | 30003961 | 0.15 | 0.02 | 2.27E-17 | 0.02 | PengQ2024NG |
| | | | | cg25540824 | 30051755 | -0.07 | 0.01 | 4.96E-11 | 0.01 | PengQ2024NG |
| | | | | cg00606578 | 30091549 | -0.08 | 0.01 | 9.39E-13 | 0.01 | PengQ2024NG |
| | | | | cg04483720 | 30276146 | 0.08 | 0.01 | 2.03E-13 | 0.02 | PengQ2024NG |
| | | | | cg01824410 | 30276563 | 0.11 | 0.01 | 8.98E-14 | 0.02 | PengQ2024NG |
| | | | | cg04679849 | 30285193 | 0.16 | 0.02 | 5.02E-23 | 0.03 | PengQ2024NG |
| | | | | cg26922678 | 30285301 | 0.27 | 0.02 | 4.02E-45 | 0.05 | PengQ2024NG |
| | | | | cg15946590 | 30285303 | 0.14 | 0.01 | 5.26E-21 | 0.02 | PengQ2024NG |
| | | | | cg05685892 | 30320135 | 0.16 | 0.02 | 2.15E-21 | 0.03 | PengQ2024NG |
| | | | | cg08416870 | 30321469 | 0.08 | 0.01 | 1.29E-11 | 0.01 | PengQ2024NG |
| | | | | cg17475918 | 30321997 | -0.08 | 0.01 | 7.44E-12 | 0.01 | PengQ2024NG |
| | | | | cg15118824 | 30322393 | -0.07 | 0.01 | 8.22E-16 | 0.02 | PengQ2024NG |
| | | | | cg09326440 | 30324478 | 0.05 | 0.01 | 1.08E-12 | 0.01 | PengQ2024NG |
| | | | | cg20105257 | 30325032 | 0.07 | 0.01 | 5.70E-38 | 0.05 | PengQ2024NG |
| | | | | cg11019014 | 30333017 | 0.12 | 0.01 | 4.82E-36 | 0.04 | PengQ2024NG |
| | | | | cg06274766 | 30360155 | 0.11 | 0.01 | 5.27E-94 | 0.11 | PengQ2024NG |
| | | | | cg21941262 | 30360271 | 0.09 | 0.01 | 1.33E-23 | 0.03 | PengQ2024NG |
| | | | | cg10692140 | 30362057 | 0.07 | 0.01 | 5.65E-27 | 0.03 | PengQ2024NG |
| | | | | cg25805999 | 30385166 | -0.06 | 0.01 | 1.20E-29 | 0.04 | PengQ2024NG |
| | | | | cg03686959 | 30400056 | 0.06 | 0.00 | 3.37E-59 | 0.07 | PengQ2024NG |
| | | | | cg09664433 | 30402058 | 0.12 | 0.01 | 2.49E-20 | 0.02 | PengQ2024NG |
| | | | | cg26484464 | 30403626 | -0.08 | 0.00 | 7.36E-52 | 0.06 | PengQ2024NG |

| | | | | | | | | | | |
|---|---|---|---|---|---|---|---|---|---|---|
| | | | | cg26884376 | 30403632 | -0.11 | 0.01 | 2.51E-21 | 0.03 | PengQ2024NG |
| | | | | cg06251929 | 30422495 | -0.09 | 0.01 | 8.41E-11 | 0.01 | PengQ2024NG |
| | | | | cg22409504 | 30423263 | 0.11 | 0.01 | 1.14E-34 | 0.04 | PengQ2024NG |
| | | | | cg12716083 | 30505359 | 0.07 | 0.00 | 9.87E-55 | 0.07 | PengQ2024NG |
| | | | | cg20378070 | 30505623 | -0.08 | 0.01 | 1.36E-19 | 0.02 | PengQ2024NG |
| | | | | cg17223971 | 30511026 | -0.04 | 0.01 | 1.66E-13 | 0.02 | PengQ2024NG |
| | | | | cg00663832 | 30517862 | 0.05 | 0.01 | 1.94E-13 | 0.02 | PengQ2024NG |
| | | | | cg18335326 | 30549943 | 0.05 | 0.01 | 6.40E-12 | 0.01 | PengQ2024NG |
| | | | | cg18611205 | 30549950 | 0.13 | 0.02 | 6.02E-15 | 0.02 | PengQ2024NG |
| | | | | cg16580700 | 30617344 | 0.09 | 0.02 | 5.32E-10 | 0.01 | PengQ2024NG |
| | | | | cg07680374 | 30620300 | 0.06 | 0.01 | 6.82E-14 | 0.02 | PengQ2024NG |
| | | | | cg24433124 | 30646413 | 0.44 | 0.04 | 2.29E-28 | 0.03 | PengQ2024NG |
| | | | | cg20802826 | 30650816 | 0.05 | 0.01 | 9.48E-15 | 0.02 | PengQ2024NG |
| | | | | cg24945895 | 30660886 | -0.06 | 0.01 | 1.06E-21 | 0.03 | PengQ2024NG |
| | | | | cg22028727 | 30662196 | 0.04 | 0.01 | 7.34E-11 | 0.01 | PengQ2024NG |
| | | | | cg25246144 | 30668906 | -0.09 | 0.01 | 4.03E-20 | 0.02 | PengQ2024NG |
| | | | | cg15910469 | 30670019 | -0.19 | 0.02 | 5.99E-32 | 0.04 | PengQ2024NG |
| | | | | cg05647770 | 30708598 | -0.10 | 0.01 | 6.24E-26 | 0.03 | PengQ2024NG |
| | | | | cg13643948 | 30708631 | -0.13 | 0.02 | 5.51E-16 | 0.02 | PengQ2024NG |
| | | | | cg08904491 | 30713612 | -0.13 | 0.01 | 7.54E-26 | 0.03 | PengQ2024NG |
| | | | | cg15516187 | 30722699 | 0.18 | 0.03 | 1.51E-07 | 0.01 | PengQ2024NG |
| | | | | cg25613385 | 30765232 | 0.02 | 0.00 | 1.66E-10 | 0.01 | PengQ2024NG |
| | | | | cg15063443 | 30765820 | -0.08 | 0.01 | 2.94E-23 | 0.03 | PengQ2024NG |
| | | | | cg27369431 | 30785588 | 0.15 | 0.01 | 9.60E-49 | 0.06 | PengQ2024NG |
| | | | | cg18796319 | 30886218 | 0.06 | 0.01 | 1.16E-12 | 0.01 | PengQ2024NG |
| | | | | cg21655448 | 30890940 | -0.10 | 0.01 | 1.81E-20 | 0.02 | PengQ2024NG |
| | | | | cg14343414 | 30918026 | 0.06 | 0.01 | 2.04E-14 | 0.02 | PengQ2024NG |
| | | | | cg08256922 | 30947631 | -0.16 | 0.02 | 6.35E-21 | 0.02 | PengQ2024NG |
| | | | | cg17663033 | 30978201 | 0.14 | 0.02 | 2.56E-19 | 0.02 | PengQ2024NG |

| | | | | | | | | | | |
|---|---|---|---|---|---|---|---|---|---|---|
| | | | | cg07613268 | 30978375 | 0.04 | 0.01 | 4.52E-11 | 0.01 | PengQ2024NG |
| | | | | cg01140214 | 31006114 | 0.07 | 0.01 | 1.71E-12 | 0.01 | PengQ2024NG |
| | | | | cg16626479 | 31006305 | -0.03 | 0.00 | 1.71E-11 | 0.01 | PengQ2024NG |
| | | | | cg00357273 | 31032058 | -0.09 | 0.01 | 1.28E-13 | 0.02 | PengQ2024NG |
| | | | | cg18923388 | 31034195 | -0.14 | 0.02 | 2.22E-14 | 0.02 | PengQ2024NG |
| | | | | cg03081173 | 31034197 | 0.11 | 0.02 | 1.83E-12 | 0.01 | PengQ2024NG |
| | | | | cg13212186 | 31034451 | 0.12 | 0.02 | 8.38E-13 | 0.01 | PengQ2024NG |
| | | | | cg27536700 | 31034491 | 0.11 | 0.01 | 1.57E-17 | 0.02 | PengQ2024NG |
| | | | | cg03921216 | 31106546 | 0.08 | 0.01 | 6.13E-14 | 0.02 | PengQ2024NG |
| | | | | cg15634083 | 31141914 | 0.28 | 0.04 | 1.39E-13 | 0.02 | PengQ2024NG |
| | | | | cg12076350 | 31141930 | 0.24 | 0.03 | 1.48E-11 | 0.01 | PengQ2024NG |
| | | | | cg02446475 | 31141998 | 0.25 | 0.03 | 2.13E-12 | 0.01 | PengQ2024NG |
| | | | | cg17250082 | 31142004 | 0.36 | 0.05 | 3.44E-15 | 0.02 | PengQ2024NG |
| | | | | cg08762424 | 31226273 | 0.31 | 0.04 | 4.41E-15 | 0.02 | PengQ2024NG |
| | | | | cg15340334 | 31322298 | 0.03 | 0.00 | 9.30E-13 | 0.01 | PengQ2024NG |
| | | | | cg13031097 | 31322577 | 0.08 | 0.01 | 2.39E-22 | 0.03 | PengQ2024NG |
| | | | | cg27529346 | 31322926 | 0.04 | 0.01 | 1.04E-10 | 0.01 | PengQ2024NG |
| | | | | cg01517384 | 31323856 | 0.09 | 0.01 | 1.76E-11 | 0.01 | PengQ2024NG |
| | | | | cg05129479 | 31323998 | -0.23 | 0.03 | 3.15E-16 | 0.02 | PengQ2024NG |
| | | | | cg02844892 | 31370412 | -0.12 | 0.02 | 7.16E-11 | 0.01 | PengQ2024NG |
| | | | | cg05054864 | 31311617 | -0.09 | 0.01 | 9.29E-13 | 0.01 | PengQ2024NG |
| | | | | cg23471770 | 31317824 | 0.09 | 0.01 | 3.11E-11 | 0.01 | PengQ2024NG |
| | | | | cg00429767 | 31321599 | -0.41 | 0.06 | 2.24E-13 | 0.02 | PengQ2024NG |
| | | | | cg06937348 | 31327310 | -0.20 | 0.03 | 6.31E-12 | 0.01 | PengQ2024NG |
| rs2160860 | 8p11.21 | 8 | 39965626 | cg08579408 | 40027353 | -0.06 | 0.01 | 1.46E-21 | 0.03 | PengQ2024NG |
| | | | | cg22073530 | 40048665 | -0.10 | 0.01 | 1.27E-37 | 0.05 | PengQ2024NG |
| | | | | cg16603385 | 40069204 | -0.07 | 0.01 | 7.61E-17 | 0.02 | PengQ2024NG |
| | | | | cg11251498 | 40069984 | 0.09 | 0.00 | 2.20E-65 | 0.08 | PengQ2024NG |
| | | | | cg12565636 | 40074712 | 0.20 | 0.01 | 4.39E-63 | 0.08 | PengQ2024NG |

| | | | | | | | | | | |
|---|---|---|---|---|---|---|---|---|---|---|
| | | | | cg10801473 | 40077695 | 0.22 | 0.01 | 2.12E-140 | 0.17 | PengQ2024NG |
| | | | | cg22831658 | 40078782 | -0.07 | 0.01 | 3.88E-24 | 0.03 | PengQ2024NG |
| | | | | cg17709637 | 40083279 | -0.22 | 0.01 | 3.81E-180 | 0.21 | PengQ2024NG |
| | | | | cg05163930 | 40083420 | -0.04 | 0.01 | 1.76E-17 | 0.02 | PengQ2024NG |
| | | | | cg14599160 | 40095865 | 0.21 | 0.01 | 2.56E-115 | 0.14 | PengQ2024NG |
| | | | | cg13975437 | 40098790 | -0.06 | 0.01 | 2.84E-15 | 0.02 | PengQ2024NG |
| | | | | cg15702662 | 40101184 | 0.08 | 0.01 | 2.55E-19 | 0.02 | PengQ2024NG |
| | | | | cg18716679 | 40113164 | 0.05 | 0.01 | 1.38E-18 | 0.02 | PengQ2024NG |
| | | | | cg05621259 | 40113227 | 0.05 | 0.01 | 2.89E-21 | 0.03 | PengQ2024NG |
| | | | | cg05893709 | 40113284 | 0.08 | 0.01 | 2.38E-29 | 0.04 | PengQ2024NG |
| rs2196122 | 11p15.4 | 11 | 4885548 | cg05046377 | 4993449 | 0.12 | 0.01 | 2.41E-69 | 0.08 | PengQ2024NG |
| | | | | cg23520574 | 4994105 | 0.12 | 0.01 | 3.42E-32 | 0.04 | PengQ2024NG |
| | | | | cg23295623 | 4994112 | 0.15 | 0.01 | 7.14E-69 | 0.08 | PengQ2024NG |
| | | | | cg00467296 | 4994293 | 0.04 | 0.00 | 7.90E-20 | 0.02 | PengQ2024NG |
| | | | | cg25641451 | 5033200 | -0.14 | 0.02 | 6.57E-08 | 0.01 | PengQ2024NG |
| | | | | cg07589078 | 5034861 | 0.26 | 0.02 | 1.25E-33 | 0.04 | PengQ2024NG |
| rs2956095 | 11p13 | 11 | 34887382 | cg00507010 | 35049708 | 0.19 | 0.01 | 6.58E-62 | 0.08 | PengQ2024NG |
| | | | | cg07578891 | 35051206 | 0.45 | 0.02 | 1.69E-91 | 0.11 | PengQ2024NG |
| | | | | cg03569764 | 35067923 | -0.21 | 0.01 | 1.79E-155 | 0.18 | PengQ2024NG |
| | | | | cg02333667 | 35072615 | -0.14 | 0.01 | 6.24E-73 | 0.09 | PengQ2024NG |
| | | | | cg25948180 | 35074715 | -0.09 | 0.01 | 1.30E-31 | 0.04 | PengQ2024NG |
| | | | | cg18508148 | 35075513 | 0.26 | 0.02 | 2.10E-55 | 0.07 | PengQ2024NG |
| | | | | cg11058730 | 35075718 | -0.14 | 0.01 | 7.95E-33 | 0.04 | PengQ2024NG |
| | | | | cg15745106 | 35076447 | 0.14 | 0.01 | 2.98E-46 | 0.06 | PengQ2024NG |
| | | | | cg24460113 | 35076814 | 0.16 | 0.01 | 1.64E-51 | 0.06 | PengQ2024NG |

| | | | | | | | | | | |
|---|---|---|---|---|---|---|---|---|---|---|
| | | | | cg21916100 | 35076822 | 0.22 | 0.01 | 2.57E-87 | 0.11 | PengQ2024NG |
| | | | | cg15818696 | 35099743 | -0.49 | 0.02 | 4.83E-184 | 0.21 | PengQ2024NG |
| | | | | cg12517288 | 35099963 | -0.11 | 0.02 | 7.93E-12 | 0.01 | PengQ2024NG |
| | | | | cg01186460 | 35146769 | -0.13 | 0.01 | 2.51E-32 | 0.04 | PengQ2024NG |
| | | | | cg21815029 | 35147417 | -0.17 | 0.02 | 2.15E-29 | 0.04 | PengQ2024NG |
| rs28550680 | 11p13 | 22 | 43950046 | cg10799608 | 44826208 | -0.20 | 0.01 | 2.71E-231 | 0.26 | PengQ2024NG |
| | | | | cg00539955 | 44879271 | -0.08 | 0.01 | 1.87E-31 | 0.04 | PengQ2024NG |
| | | | | cg01313284 | 44883532 | -0.06 | 0.01 | 1.61E-12 | 0.01 | PengQ2024NG |

**Table S7.** Allele frequency comparison of novel risk-associated SNPs across Chinese and East Asian populations.

| SNP | A1 | A2 | CHR | BP | Freq_case | Freq_control | Freq_gnomAD (East Asian) | Freq_genomeasia100k (Northeast Asia) |
|---|---|---|---|---|---|---|---|---|
| rs3219489 | G | C | 1 | 45331833 | 0.4633 | 0.3761 | 0.415 | 0.3632 |
| rs2165738 | C | G | 2 | 24469940 | 0.4924 | 0.3346 | 0.316 | 0.359 |
| rs10024137 | A | G | 4 | 68667043 | 0.07474 | 0.2148 | 0.285 | 0.233618 |
| rs3733631 | C | G | 4 | 103719946 | 0.4451 | 0.3386 | 0.359 | 0.352601 |
| rs2962370 | G | C | 5 | 17215335 | 0.377 | 0.3466 | 0.34 | 0.3006 |
| rs272868 | C | G | 5 | 132345058 | 0.4068 | 0.389 | 0.402 | 0.4046 |
| rs9358491 | A | G | 6 | 22139885 | 0.2183 | 0.362 | 0.333 | NA |
| rs2534815 | C | G | 6 | 30531350 | 0.4195 | 0.3635 | 0.345 | 0.3191 |
| rs2160860 | A | T | 8 | 39965626 | 0.3724 | 0.426 | 0.421 | 0.453 |
| rs2196122 | C | G | 11 | 4864318 | 0.4936 | 0.326 | 0.35 | 0.314814 |
| rs2956095 | C | T | 11 | 34887382 | 0.35 | 0.2042 | 0.158 | 0.1852 |
| rs28550680 | T | C | 22 | 43950046 | 0.1734 | 0.3348 | 0.32 | 0.307692 |

**Table S8.** Associations of novel risk loci in the replication cohort.

| SNP | CHR | POS | A1 | A2 | OR | L95 | U95 | Z_STAT | P | Gene | Locus | Annotation |
|---|---|---|---|---|---|---|---|---|---|---|---|---|
| rs3219489 | 1 | 45331833 | G | C | 1.05 | 0.69 | 1.58 | 0.22 | 0.83 | *MUTYH* | 1p34.1 | new |
| rs2165738 | 2 | 24469940 | C | C | 1.03 | 0.67 | 1.57 | 0.12 | 0.90 | *NCOA1* | 2p23.3 | new |
| rs10024137 | 4 | 68667043 | A | G | 0.99 | 0.58 | 1.67 | -0.05 | 0.96 | *UGT2B15* | 4q13.2 | new |
| rs3733631 | 4 | 103719946 | C | G | 0.94 | 0.64 | 1.40 | -0.29 | 0.77 | *TACR3* | 4q24 | new |
| rs2962370 | 5 | 17215335 | G | G | 0.96 | 0.62 | 1.49 | -0.19 | 0.85 | *BASP1* | 5p15.1 | new |
| rs272868 | 5 | 132345058 | C | C | 1.48 | 1.00 | 2.20 | 1.94 | 0.05 | *SLC22A4/ SLC22A5* | 5q31.1 | new |
| rs2631372 | 5 | 132367886 | G | C | 1.50 | 1.01 | 2.23 | 2.00 | 0.05 | *SLC22A4/ SLC22A5* | 5q31.1 | new (in high LD with rs272868, $r^2 = 0.99$) |
| rs2160860 | 8 | 39965626 | A | A | 1.22 | 0.84 | 1.77 | 1.06 | 0.29 | *IDO1/IDO2* | 8p11.21 | new |
| rs2196122 | 11 | 4864318 | C | G | 0.80 | 0.51 | 1.26 | -0.96 | 0.34 | *MMP26* | 11p15.4 | new |
| rs2956095 | 11 | 34887382 | C | C | 0.51 | 0.29 | 0.93 | -2.21 | 0.03 | *APIP/PDHX* | 11p13 | new |
| rs28550680 | 22 | 43950046 | T | C | 1.31 | 0.87 | 1.99 | 1.29 | 0.20 | *PNPLA3/SAMM50* | 22q13.31 | new |

**Table S9.** gRNA Primers for rs2631372 and rs272868.

| Primer name | | Primer sequence | Antibiotic screening |
|---|---|---|---|
| rs2631372-gRNA | gRNA-1 | F-caccGAACTGGACGGCACCATCTT | puromycin |
| | | R-aaacAAGATGGTGCCGTCCAGTTC | |
| | gRNA-2 | F-caccGTAGCTATTGGTTATCTTAT | neomycin |
| | | R-aaacATAAGATAACCAATAGCTAC | |
| rs272868-gRNA | gRNA-1 | F-caccGCAGATTGGCCAGATGAGCC | puromycin |
| | | R-aaacGGCTCATCTGGCCAATCTGC | |
| | gRNA-2 | F-caccGCTAATGAGGTTACTTAATG | neomycin |
| | | R-aaacCATTAAGTAACCTCATTAGC | |

**TableS10.** Primer design for qPCR gene determination.

| Gene | Full Gene Name | Transcript Variant | RNA Type | Primer sequence | NCBI Reference Sequence |
|---|---|---|---|---|---|
| *SLC22A4* | Homo sapiens solute carrier family 22 member 4 | N/A | mRNA | SLC22A4-1F: GCGAATTCGCCACCatgcgggactacgacgaggt | NM_003059.3 |
| | | | | SLC22A4-1704R: GTCGGATCCgaatgcagttattagaaccttggg | |
| *SLC22A5* | Homo sapiens solute carrier family 22 member 5 | Transcript Variant 1 | mRNA | SLC22A5-1F: ggtGAATTCGCCACCatgcgggactacgacgaggt | NM_001308122.2 |
| | | | | SLC22A5-1794R: CGAGGATCCgaaggctgtgcttttaaggat | |
| *LOC553103* | Homo sapiens MIR3936HG host gene | N/A | Long Non-Coding RNA | MIR3936HG-1F: AGCGAATTCGCGCGTGCGCGCGGGGCACC | NR_110997.1 |
| | | | | MIR3936HG-1793R: TCGCGGCCGCGTCATTGGCATCTGTTTGTTTATTTATT | |

| | | | | | |
|---|---|---|---|---|---|
| *PDLIM4* | Homo sapiens PDZ and LIM domain 4 | Transcript Variant 1 | mRNA | PDLIM4-1F: AGCGAATTCgccaccATGCCCCATTCCGTGAC<br>PDLIM4-1000R: TCGCGGCCGCGctaCTTGTCATCGTCGTCCTTGTAATCGACGAGTTCCACCTTGGC | NM_003687.4 |
| *P4HA2* | Homo sapiens prolyl 4-hydroxylase subunit alpha 2 | Transcript Variant 1 | mRNA | P4HA2-1F: GATTCTAGAGCCACCatgaaactctgggtgtctgc<br>P4HA2-1700R: CGCGGATCCtcagtcaacttctgttgatccaca | NM_004199.3 |
